# Geographic Distributions of Top Beef Salmonella Serovars in the U.S.

**DOI:** 10.64898/2026.09.17.26363322

**Authors:** Tatum S. Katz, Tatum D. Mortimer, Amy T. Siceloff, Nikki W. Shariat, Dayna M. Harhay, Joshua M. Blackstock, Tommy L. Wheeler

## Abstract

We live in an era where advances in microbial detection, DNA sequencing technology, and the availability of big data have allowed inroads to the field of predictive microbiology. The tools presently in use, as well as those on the horizon, provide an opportunity to examine emerging infectious diseases in ways previously not possible. We analysed publicly available data to investigate where and why *Salmonella* is detected on raw beef products. In addition to analysing beef-related serovars across space and time, we utilised a machine-learning approach (Species Distribution Modelling) to identify areas and predictors of environmental suitability for the most frequent *Salmonella* serovars found in beef. Our results demonstrate that some serovars, such as Dublin, Montevideo, and Muenchen, exhibit strong regional prevalence while others, notably Anatum, are found broadly throughout the country. The Species Distribution Models show concurrence with these regional trends. Predictors that contributed 10% or more to multiple serovar models included ecoregions (7/15 serovars), avian flyways (8/15), the number of local cattle operations (3/15), and other environmental variables. Our work demonstrates that certain *Salmonella* serovars commonly associated with beef show regional trends that can be predicted.

## Introduction

Non-typhoidal *Salmonella enterica* (hereafter *Salmonella*) is the leading bacterial foodborne pathogen in the United States and worldwide. It is estimated that approximately 1.3 million people in the United States suffer from illnesses caused by non-typhoidal *Salmonella* annually [1]. Consumption of contaminated beef products are implicated in 6.9% of domestic salmonellosis cases [2]. Despite considerable efforts and a decrease in *Salmonella* on raw meat products [3], the control of *Salmonella* remains a recalcitrant problem in the United States. In fact, beef-attributed *Salmonella* outbreaks have occurred at an unchanged rate over the past two decades [4].

A major challenge in the control of *Salmonella* is its high biological diversity. There are over 2,600 different serovars, and these can differ significantly in their pathogenicity and host specificity [5–7]. Despite this extremely high diversity, only 13 serovars represent 99.5% of recent beef-attributed *Salmonella* outbreak illnesses [7,8], and of the 42 beef outbreaks that have occurred from 2009 to 2023, just 16 different serovars have been implicated [4]. A growing body of work highlights the underlying genetics responsible for why certain serovars cause illness [9–11]. This indicates that targeting specific serovars, rather than *Salmonella* as a whole, may be more effective in reducing beef-attributed salmonellosis.

*Salmonella* serovars that are prevalent in a certain product (e.g., beef) and that are also linked to significant human illness are considered serovars of concern (SoCs) for that product. While molecular techniques continue to advance, allowing the field to by-pass conventional serotyping as the major subtyping method, focusing on SoCs can help to target the most clinically relevant serovars in the meantime. SoCs have been defined using epidemiological data [7,8,12], and genomic data [9,13]. In beef, the top three SoCs based on burden and trajectory of outbreak illnesses [12] and number of outbreaks, outbreak size, number of illnesses, and hospitalization rate [7] are Newport, Typhimurium, and Dublin.

The diversity in *Salmonella* pathogenicity may be reflected in other aspects of the pathogen, such as drivers of occurrence. Over the last decade, predictive modelling methods have gained attention for their ability to identify trends and predictors of infectious disease [14–18]. These tools exploit the growing wealth of machine learning and artificial intelligence approaches to generate predictions about how, where, and why pathogens occur. Popular techniques that have been applied to foodborne pathogens include random forest [19,20], neural networks [15,21], time series [22,23], and ensemble approaches [24], though techniques vary widely. Predictive models can incorporate climatic data, other environmental variables such as land use or topography, and disease surveillance data to identify system levers that could be pulled for greater disease control. Focusing on *Salmonella* specifically, environmental predictors such as climate and meteorological factors have been used to estimate salmonellosis outbreaks, incidence, and risk. Many studies report higher *Salmonella* loads in the warmer seasons and/or hotter regions [19,25–31], but the underlying mechanism has not been determined and may be related to climate and meteorological changes. These findings span from preharvest to postharvest: Smith et al [32] identified precipitation and temperature metrics as correlative with *Salmonella* prevalence on meat products. There also exists a great deal of observational evidence and support in the literature [19,25,26,28,30,31,33] of regionality in *Salmonella* risk. These regional trends may additionally be explained by regional soil composition, as *Salmonella* is a soil-dwelling pathogen [34,35]. Alternatively, avian migration pathways continue to be linked to an increasing number of zoonotic diseases including *Salmonella* [36–40], representing another possible mechanism driving regional *Salmonella* patterns.

Based on these previous findings we sought to develop a broader analysis of the relationship between *Salmonella* prevalence and environmental variables. Due to the rising importance of SoC control and evidence of regional serovar trends, we wanted to investigate whether environmental preferences differed between serovars found in beef. Here we interrogate the spatial occurrence of beef-related *Salmonella* serovars and introduce Species Distribution Modelling (SDM) as a novel tool for modelling *Salmonella* to understand serovar distribution throughout the US and to test hypotheses on why certain serovars show up in certain areas. Specifically, we tested the hypothesis that the spatial distribution of *Salmonella* serovars on raw beef products at processing facilities across the United States could be predicted when utilising climate, ecoregions, agriculture facilities, avian flyways, and soil geochemistry features.

## Materials and Methods

### *Salmonella* occurrence on raw beef products

To understand *Salmonella* populations on raw beef products, we downloaded data from the Raw Beef Sampling program from USDA’s Food Safety Inspection Service (FSIS) on 12 February 2025 [3]. These data describe regulatory sampling of pathogens from various raw beef products across establishments in the U.S. from 2014 to 2024. From 197 913 total observations, we selected only those samples which were analysed for *Salmonella*, providing 184 370 samples for analysis. These data were cleaned differently for the two analyses, the spatial patterns and species distribution modelling, based on the sensitivity of those analyses to spatial and temporal biases in the data. Details on data cleaning are in the respective sections.

### Geospatial predictors

We hypothesized that the spatial distribution of *Salmonella* on raw beef products at processing facilities across the United States could be predicted by climate, ecoregions, agriculture facilities, avian flyways, and soil geochemistry. All spatial data were rasterised, cropped to the 48 contiguous United States of America (CONUS), and resampled to a 2.5 arc minute (∼4.6km near the equator) resolution as needed.

Climate. 19 variables describing temperature and rainfall were collected from WorldClim.org [41]. These variables are commonly used in SDM for their biological relevance [41,42]: annual mean temperature, mean diurnal range, isothermality, temperature seasonality, maximum temperature of the warmest month, minimum temperature of the coldest month, temperature annual range, mean temperature of the wettest quarter, mean temperature of the driest quarter, mean temperature of the warmest quarter, mean temperature of the coldest quarter, annual precipitation, precipitation of the wettest month, precipitation of the driest month, precipitation seasonality, precipitation of the wettest quarter, precipitation of the driest quarter, precipitation of the warmest quarter, precipitation of the coldest quarter. These variables were downloaded as GeoTiffs at the 2.5 arc minute scale.

Ecoregions. Defined areas where ecosystems and their associated environmental resources are generally similar [43]. We downloaded the province-level Bailey ecoregions [44] from (https://www.sciencebase.gov/catalog/item/620da87bd34e6c7e83ba9ba7, [45]) as netCDF files on a 25km grid. The file was subsequently rasterized, cropped to CONUS, and resampled to a 2.5 arc minute resolution, resulting in 42 ecoregions (Supplementary File 1).

Agriculture Facilities. Data describing county-level agricultural variables was downloaded from the USDA National Agricultural Statistic Service’s 2022 census through their Quick Stats tool [46] as a text file. We developed the following seven variables from the census data at the county level for the year 2022: ‘n cattle operations’, the total number of cattle operations; ‘total value of cattle’, the total value of cattle in dollars; ‘n other livestock operations’, the total number of livestock operations excluding cattle; ‘total value of other livestock’, the total value of other livestock excluding cattle in dollars; ‘n poultry operations’, the number of poultry operations; ‘total value of poultry’, the total value of poultry in dollars; ‘n head other livestock’, the total number of head of livestock excluding cattle; and ‘n produce operations’, the number of produce operations. Poultry is defined as chicken, turkey, other game meats such as quail and pheasants, and large birds including ostriches and emus [46]. These data are organised to the county level. To develop a spatial raster for analysis we mapped these values to county shapefiles from the U.S. Census Bureau at a 5m resolution (https://www.census.gov/geographies/mapping-files/2017/geo/carto-boundary-file.html) and rasterized them. For some variables in some counties, there are not enough operations to meet the NASS data disclosure criteria, and these are marked with a (D) rather than a numeric value. Following a sensitivity analysis to evaluate how imputation of these values affected the model, the redacted values were imputed with a 1 for analysis.

Avian Flyways. A shapefile describing the four U.S. Fish and Wildlife Service Administrative Waterfowl Flyway Boundaries was downloaded from https://iris.fws.gov/APPS/ServCat/Reference/Profile/42276. These flyways are major pathways used by migratory birds and are formed latitudinally across North America. From east to west, they are the Atlantic Flyway, the Mississippi Flyway, the Central Flyway, and the Pacific Flyway.

Soil Geochemistry. Data describing soil composition were downloaded from two sources, the University of California at Davis Soil Resource Lab (https://casoilresource.lawr.ucdavis.edu/soil-properties/download.php, [47]) and the USGS Data Series 801 (https://pubs.usgs.gov/ds/801/downloads/, [48]). From the U.C. Davis soil property data, we downloaded GeoTiffs describing pH, calcium carbonate, cation exchange capacity, electrical conductivity, soil organic matter, soil texture, and drainage class for the top 0-5cm of soil at an 800m resolution. From the USGS Data Series 801, we additionally downloaded text files describing calcium, copper, iron, potassium, manganese, sodium, phosphorus, sulfur, and zinc from the top 0-5cm of soil at a resolution of 1 site per 1600 square kilometers, resulting in 4857 sites across the CONUS [48]. We followed the inverse distance weighted interpolation protocol described in [48] for interpolation of these variables to develop rasters for spatial analysis.

### Spatial patterns of top serovars

To gain a better understanding of any regional serovar trends, we divided the FSIS raw beef sampling results into eight production regions, as set by The Beef Industry Food Safety Council (BIFSCo). Only samples collected as part of standard regulatory operations were included (project codes “MT43”, “MT60”, “MT60_C”, “MT64”, “MT65”, and “MT65_C”), spanning from 2014 to 2024. The regions were defined as follows: 1, Washington, Oregon, Idaho; 2, California, Nevada; 3, Arizona, New Mexico, Texas; 4, Wyoming, Montana, Utah, Colorado; 5, South Dakota, Nebraska, North Dakota, Minnesota, Wisconsin; 6, Missouri, Iowa, Kansas; 7, Arkansas, Louisiana, Alabama, Georgia, Mississippi, Florida, South Carolina, North Carolina, Oklahoma, Tennessee; 8, Michigan, Illinois, Indiana, Ohio, Kentucky, West Virginia, Maryland, Virginia, Pennsylvania, New York, New Jersey, Vermont, Maine, New Hampshire, Connecticut, Massachusetts, Rhode Island, Washington D.C., Delaware. Samples collected from outside of the continental United States were not included in the overall analysis. Only serovars that were ranked in the top 10 most abundant annually are shown (n = 17).

### Species Distribution Modelling of top serovars

To predict suitable habitat for top beef-associated *Salmonella* serovars, we developed a suite of SDMs; these use geospatial data, occurrence locations of a species, and machine learning algorithms to make species-specific predictions about the most important influencing variables and where it could possibly occur, i.e. habitat suitability.

We used the FSIS raw beef sampling data as our occurrence points for the SDMs [3]. These samples are collected at processing establishments rather than the point of origin of the animal, therefore, we limited our samples to those associated with project codes “MT60”, “MT64”, “MT60_C”, and “MT64_C”, which are trim (60) and ground beef components (64) and are usually sourced locally [49]. We eliminated the “MT43” and “MT65” project code samples that correspond to ground beef and bench trim samples, as these usually come from locations far from the processing establishment. We additionally truncated the data to 2016 when sampling became more standardized [49]. This resulted in 1239 positive *Salmonella* isolates for analysis.

We limited our analysis to serovars with at least 21 occurrence points to ensure enough data to fit reasonably complex models [50,51]. The 15 serovars that met this threshold were Agona, Anatum, Cerro, Dublin, Give, I 4,[5],12:i:-, Infantis, Kentucky, Mbandaka, Meleagridis, Montevideo, Muenchen, Muenster, Newport, and Typhimurium. For each serovar, we used the algorithm MaxEnt [52] to generate an SDM from the provided predictors.

Model development proceeded as follows. ENMeval [53] was used for model selection and validation. Locality points from the FSIS Raw Beef Sampling dataset were used as occurrence points and the 52 geospatial variables described above were used as predictors. Allowed features were linear, quadratic, product, and threshold, and the regularization multiplier was varied from 0.5 to 4 in increments of 0.5. All other settings were left at the default. For validation the data were partitioned in a jackknife (leave-one-out) fashion and the maxent.jar algorithm was used. ENMeval returns a suite of models with Area Under the receiver operating characteristic Curve (AUC) and Akaike’s Information Criterion (AICc, with a correction for small sample sizes) values; the model with the lowest AICc was chosen as the best model, unless a model with comparable AICc (delta AICc<2) returned a higher Training AUC, in which case the next best model by AICc which had a higher AUC was selected [51,54]. The best model for each serovar was interrogated for the variables included and habitat suitability predictions were plotted.

## Results

### Spatial patterns of top serovars

The overall serotyping results from the FSIS regulatory beef dataset are relatively consistent between the years, with serovars Anatum, Dublin, Montevideo, and Muenchen prevalent throughout (Figure 1A). The serovars shown include those present in the top 10 of at least one region between 2014 – 2024. From the final list (n = 17), all serovars were found annually in at least one region, with the exception of serovars Brandenburg and Give in 2014, 2016, 2017 and 2014, respectively. Serovar Montevideo was the most abundant overall (24%, n = 609/2549), followed by serovars Anatum (11%, n = 270), Muenchen (9%, n = 232), and Dublin (8%, n = 214).

**Figure 1.**
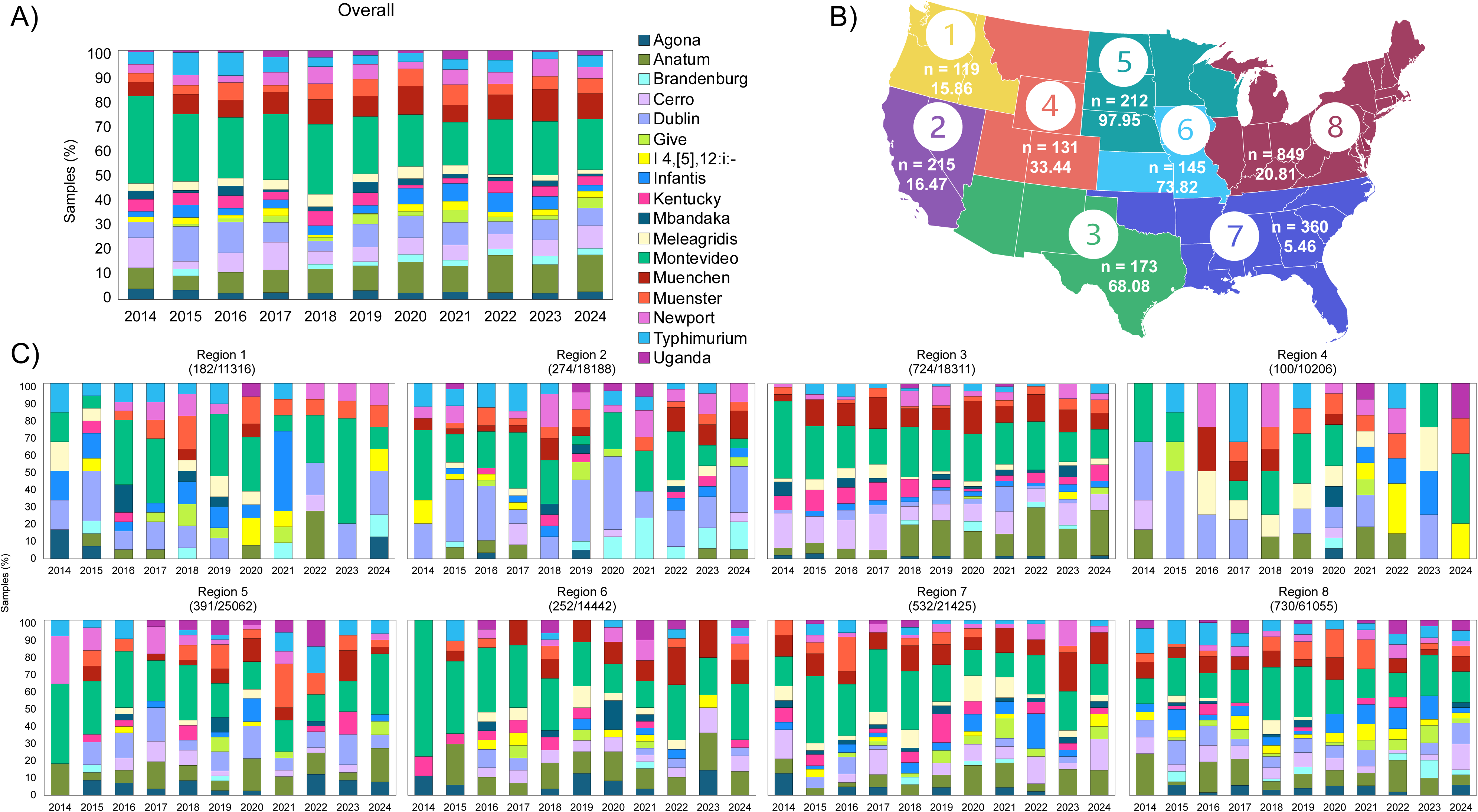
Top serovars reported in beef products sampled by FSIS from 2014 to 2024 and regional differences. A) Overall results for the top 17 serovars found in beef products in the continental United States. B) Map of production regions. Large numbers inside circles give the regions, n gives the total number of establishments sampled, and the number beneath is the total head of cattle slaughtered in that region from 2014 to 2024 in millions. C) Regional serotyping results across the eight production regions. Number of *Salmonella*-positive and total samples collected listed in parenthesis. Alt text: Graphs showing the relative abundances of top beef serovars across years and BIFSCo regions, with a map showing the BIFSCo regions.

To consider the impact of each beef production region (Figure 1B), the FSIS surveillance results were divided respectively. Region 8 has both the greatest number of states (n = 16) and, subsequently, number of processing establishments sampled (n = 849), despite relatively low slaughter numbers in total (20.8 million head of cattle). Conversely, Region 1 had the lowest number of establishments (n = 119) with three states included. The serotyping results had greater variance year-to-year when split across the regions. Serovars Montevideo and Anatum were present across all regions and were relatively consistent (Figure 1C). Regions 1 and 4 were the smallest in number of facilities and also in terms of positives, but not in terms of head slaughtered. This appears to have manifested in more instability in the relative frequency of different serovars over the 10-year period.

The relative frequency of serovars found in Region 3 was consistent over time, with the exception of serovar Anatum displacing serovar Cerro, perhaps reflecting a larger production volume (68 million head slaughtered). Some regional trends were identified. For example, there was a higher incidence of serovar Dublin in regions associated with higher dairy production (Regions 1, 2, 5, 8) than those without. For other serovars, regional differences appeared to reflect a disruption from the norm, such as the reduced presence of serovar I 4,[5],12:i:- from Regions 3 and 5 and of serovar Kentucky from Regions 1, 2, and 4. We also observed serovars that were present in adjacent regions. For example, 73% (37/51) of Uganda isolates were collected in regions 5, 6, and 8, which suggests a possible northeast signal. Additionally, 59% (35/59) of Give isolates originated from regions 7 and 8, representing the east coast. Of the I 4,[5],12:i:- isolates, 44% (24/54) were found in Region 8.

### Geospatial predictors of top serovars

To examine the possibility of spatially explicit predictors of *Salmonella* serovar, we utilised SDMs. These models use occurrence data of a species (here, a *Salmonella* serovar, determined using USDA-FSIS data) and gridded, geospatial predictors to calculate the probability of the given species’ habitat suitability across the landscape (22). Areas with similar climate to the occurrence data are typically scored with high probability of suitability, and dissimilar areas are scored with low probability of suitability. Using the developed SDMs, we generated predictions for the distributions of serovars Agona, Anatum, Cerro, Dublin, Give, I 4,[5],12:i:-, Infantis, Kentucky, Mbandaka, Meleagridis, Montevideo, Muenchen, Muenster, Newport, and Typhimurium for the contiguous United States (Figure 2). Details for each model are available in Supplementary Files 2 through 16.

**Figure 2.**
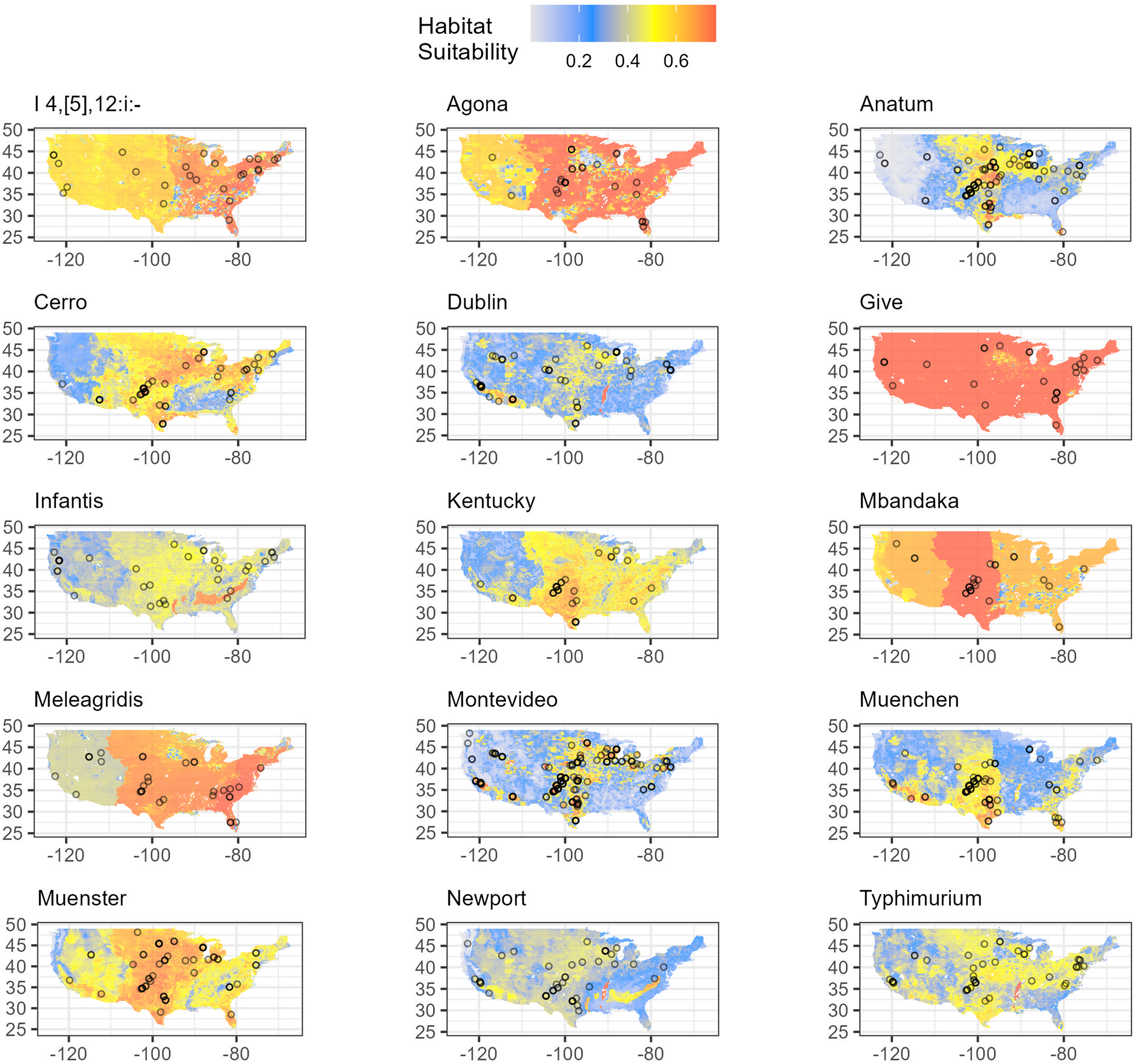
Predictions of serovar distributions based on geospatial predictors. Individual circles represent a single sample of the given serovar. Alt text: Heat maps showing habitat suitability as predicted by the SDMs for 15 beef serovars in the United States.

Generally, model fitting and validation was successful with most AUC scores over 0.70 indicating good prediction (Table 1), except for serovar Give whose top model returned an AUC of just 0.43, indicating model prediction is worse than random chance. The models for serovars Mbandaka and Meleagridis returned weak AUC scores indicating only moderate prediction (0.67 and 0.69 respectively). Additionally, serovar Kentucky’s lowest AICc model produced an AUC of 0.75 yet a second model with a delta AICc of only 0.14 returned an AUC of 0.83 and therefore this second-best model for Kentucky was retained.

**Table 1.** Model tuning results for all serovars. All fitted models with delta AICc < 2 were considered for selection. The model with the lowest AICc was chosen unless a model with comparable AICc (delta AICc<2) returned a higher Training AUC, in which case the next best model by AICc which had a higher AUC was selected. When two models are present for a serovar, selected best models are denoted with an *. P indicates the number of parameters in the model.

| Feature Classes | Regularization Multiplier | Training AUC | AICc | delta AICc | p | Serovar |
| --- | --- | --- | --- | --- | --- | --- |
| LQPT | 4.0 | 0.72 | 378.40 | 0.00 | 7 | Agona |
| LQPT | 3.5 | 0.87 | 779.29 | 0.00 | 15 | Anatum |
| LQPT | 4.0 | 0.86 | 396.30 | 0.00 | 8 | Cerro |
| LQPT | 4.0 | 0.89 | 461.99 | 0.00 | 8 | Dublin |
| LQPT | 4.0 | 0.42 | 202.78 | 0.00 | 1 | Give |
| LQPT | 4.0 | 0.79 | 355.64 | 0.00 | 7 | I 4,[5],12:i:- |
| LQPT | 3.0 | 0.83 | 353.77 | 0.00 | 5 | Infantis |
| LQPT | 3.5 | 0.83 | 354.82 | 0.14 | 4 | Kentucky* |
| LQPT | 4.0 | 0.75 | 354.69 | 0.00 | 3 | Kentucky |
| LQPT | 4.0 | 0.67 | 306.36 | 0.00 | 3 | Mbandaka |
| LQPT | 4.0 | 0.69 | 411.66 | 0.00 | 5 | Meleagridis |
| LQPT | 4.0 | 0.87 | 1078.06 | 0.00 | 18 | Montevideo |
| LQPT | 4.0 | 0.85 | 699.99 | 0.00 | 16 | Muenchen |
| LQPT | 4.0 | 0.74 | 530.21 | 0.00 | 5 | Muenster |
| LQPT | 3.5 | 0.86 | 458.26 | 0.00 | 9 | Newport |
| LQPT | 4.0 | 0.84 | 571.94 | 0.00 | 8 | Typhimurium |

Visual inspection of the predictions reveals important spatial differences among predicted serovar distributions (Figure 2). Notably, serovars Dublin, Montevideo, Muenchen, and Muenster have a strong and distinct West Coast signal, as indicated by the respective yellow-red shading with higher habitat suitability (>60%) in this area. In Florida, serovars I 4,[5],12:i:-, Agona, Anatum, Cerro, Meleagridis, Muenchen, and Muenster are highly suited. Serovars Anatum, Montevideo, and Muenchen have strong plains area signal. Finally, serovars I 4,[5],12:i:-, Agona, Cerro, Meleagridis, and Montevideo have a strong northeast grouping. Most serovars have broad habitat suitability across the U.S., such as Anatum, while serovars Dublin and Muenchen are more limited (Figure 2).

Overall, the most important predictors (contributing to at least 10% of the model variance) for the presence of any of the 15 serovars were, in descending order of frequency, ecoregions, avian flyways, the number of cattle operations in the county, the total value of other livestock in the county, and mean temperature of the warmest quarter (Figure 3A, 4). Serovar distribution models were separated into two categories concerning potential public health impact: i) SoC (I 4,[5],12:i:-, Dublin, Infantis, Montevideo, Muenchen, Newport, and Typhimurium) and ii) non-SoC (Agona, Anatum, Cerro, Give, Kentucky, Mbandaka, Meleagridis, and Muenster) (Katz et al 2024) to look for differences in important predictors for these two groups. Ecoregions were a top predictor in 5/7 SoC but only 2/8 non-SoC SDMs. Avian flyways were a top predictor in 6/8 non-SoC models but were only a top predictor in 2/7 SoC models. Finally, the number of cattle operations in the county was a top predictor in 3/7 SoC models but none of the non-SoC models (Figure 3B, 4).

**Figure 3.**
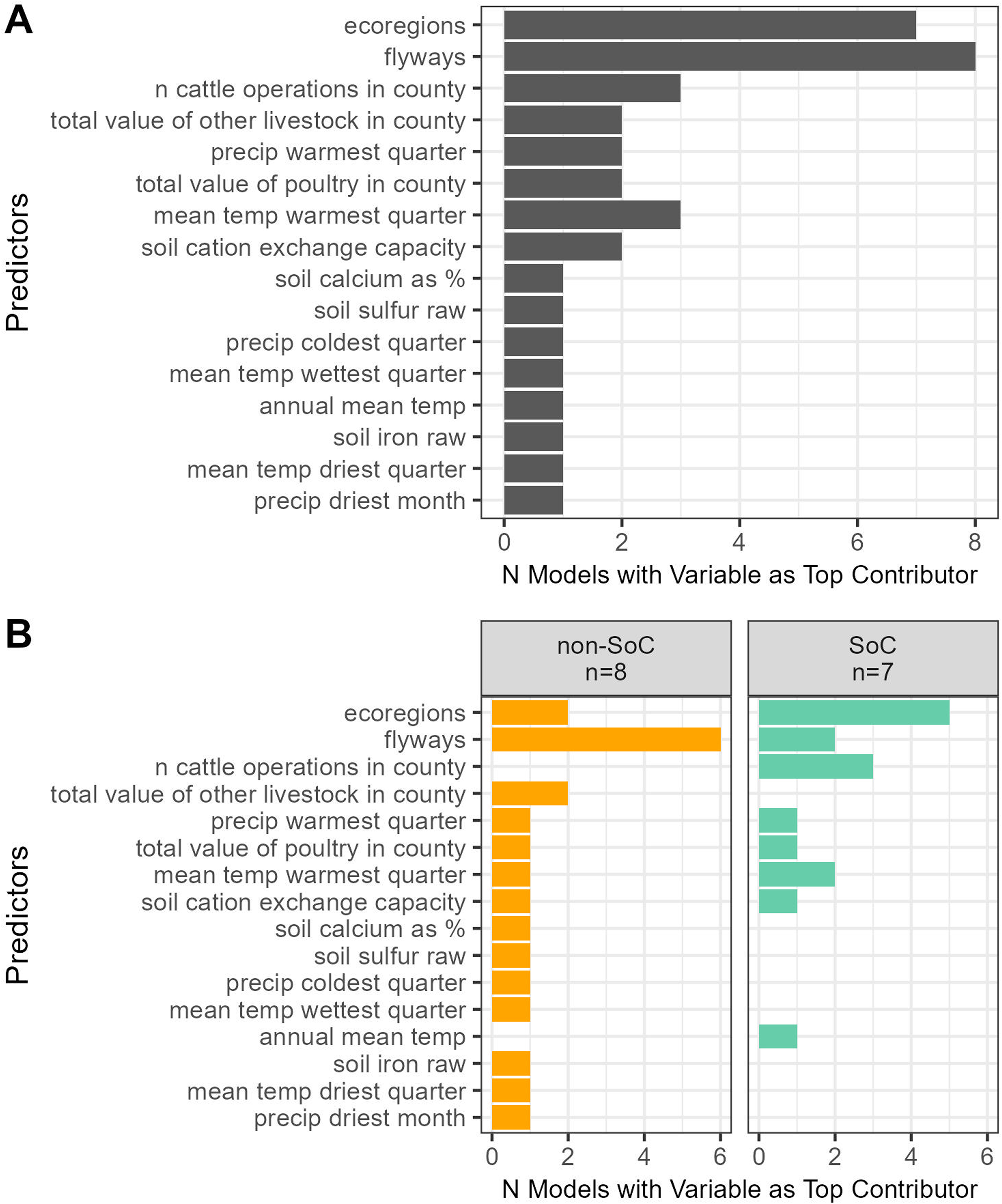
(A). Predictors which contributed at least 10% to model fitting were identified for each serovar. (B) Most important predictors for the distributions of SoC and non-SoC serovars. Only predictors which contributed 10% or more to the distribution predictions were included. Alt text: Bar plot showing how many times variables contributed 10% or more of the predictions for the 15 serovar models, broken down by serotypes of concern classification.

**Figure 4.**
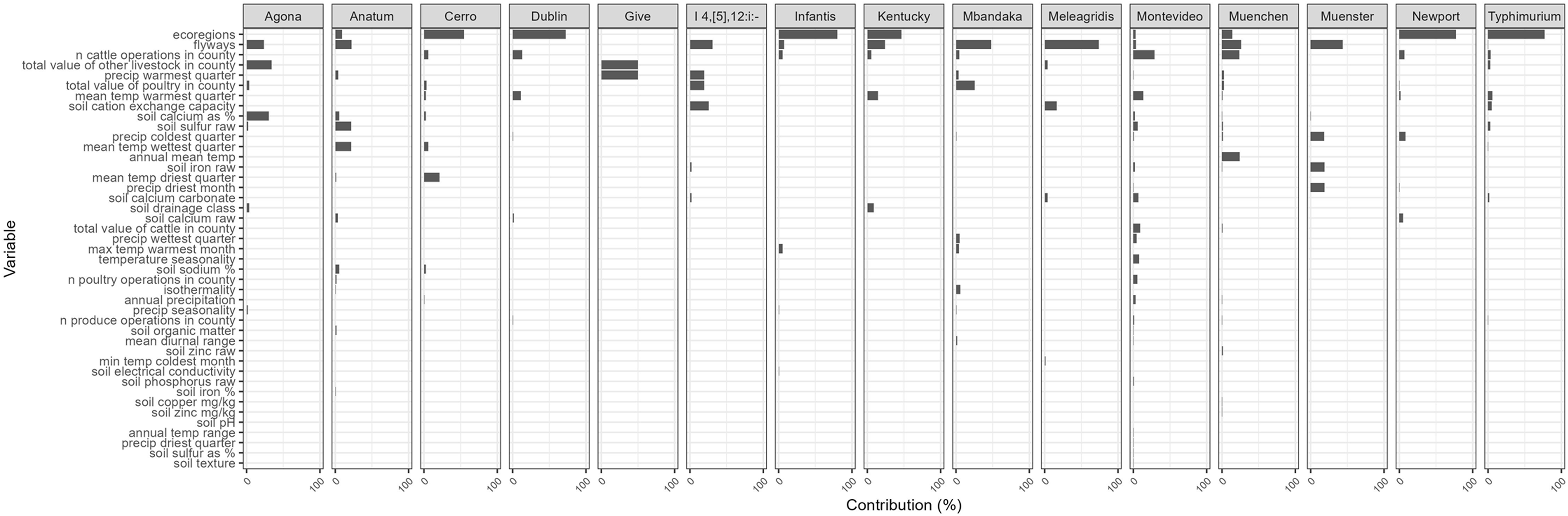
Variable contributions for all 15 serovar models. Contributions describe how important that variable is for model predictions, but do not describe direction or shape of the relationship. Alt text: Bar plots showing contributions of all variables to the 15 serovar models.

## Discussion and Conclusions

Managing beef-related *Salmonella* continues to be a significant challenge. The species is highly diverse, with most serovars causing reduced concern for human health while a select few (SoCs) cause outbreak events. We sought to understand spatial patterns and predictors of beef-related *Salmonella* to evaluate them as possible management tools.

In the exploratory spatial analysis, we found that serovar prevalence was relatively consistent through time, with most of the top serovars being found annually in some regions across the CONUS. This indicates that emergence or disappearance of important serovars is a rare occurrence. However, we also demonstrate that year-to-year variance increases at the regional level, with certain serovars occurring primarily in a set of adjacent regions: Uganda in regions 5, 6, and 8 representing the Midwest and Northeast; Give in regions 7 and 8 representing the East Coast; and I 4,[5],12:i:- also primarily from region 8 in the Northeast. These results are partially mirrored in our SDMs. While Uganda did not have enough data for modelling and the Give model failed, we observed a strong Northeast signal for I 4,[5],12:i:-. These findings together support our hypothesis that the risk of each serovar’s occurrence is not distributed homogenously across the U.S., and that instead serovars appear to have some regional linkages in space.

Our SDM results further corroborate that *Salmonella* serovars in beef appear to be associated with particular geographic regions above random chance. These regional trends may be explained in part by variables other than cattle operation distribution, such as the ecoregion, avian flyway, and mean temperature in the warmest quarter. Additionally, we identified combinatorial effects among some variables, such that two together predict *Salmonella* prevalence. While we found there is overlap in the distribution of the examined serovars, distinct differences in regionality can inform the meat industry on which serovars could be targeted for their locality. As *Salmonella* management shifts from eliminating all *Salmonella* to targeting just the most relevant variants, this information can be useful to guide resource allocation for surveillance, biosecurity, and related decision making. Understanding regional trends and potential drivers can empower beef industry producers to develop management strategies that prioritize resources for the most likely serovars of concern in their production systems.

While model fitting and validation was generally successful, with AUC values over 0.70 for 12 out of the 15 SDMs, three models failed to achieve sufficiently high AUC scores (AUC >= 0.70 indicating acceptable prediction). These models were for serovars Give (0.41), Mbandaka (0.67), and Meleagridis (0.69). AUC scores can range from 0 to 1, where 1 is perfect prediction and 0.5 is equivalent to random prediction [55]. These low values may be due to the major limitation of this approach: it is necessary to assume that serovar occurrence is related to the location of the production facility rather than where the cattle were finished. No standardized data exist regarding serovar occurrence on finishing cattle, therefore, the FSIS Raw Beef Sampling data which occurs at processing facilities is currently the best proxy. To mitigate the bias introduced by this assumption, we selected relatively local product sample types. Specifically, the production of ground beef at a single processing establishment usually supplements in-house trimmings with purchased trimmings to adjust lean:fat ratios of final ground product and often these originate in different regions or may even be imported from other countries. Further, previous findings demonstrate that the final finishing location of cattle has a significant impact on the *Salmonella* load and serovar prevalence [56], yet this may not be sufficient to accurately model distributions for all serovars. *Salmonella* serovar occurrence can be different on finished product compared to pre-harvest samples, therefore this is the major limiting assumption of our analysis.

While top predictors for each serovar remained relatively consistent (ecoregions, flyways, and the number of cattle operations), there is variety in the remaining identified predictors. Non-cattle meat animal operations appear to be associated with beef-related serovar distributions, although these relationships are not always positive or linear and often occur as interactions with other terms (Supplementary Files 2 through 16). Specifically, non-cattle livestock (Agona, Meleagridis, and Typhimurium), poultry (Agona, Anatum, Cerro, I 4,[5],12:i:-, Mbandaka, Montevideo, Muenchen, and Newport), and produce (Dublin, Montevideo, Muenchen, and Typhimurium). These relationships may arise due to shared/adjacent land with other animal operations.

Temperature variables are predictive for all serovars except I 4,[5],12:i:- and Muenster, while precipitation variables were found to be predictive for every serovar except Kentucky, Meleagridis, and Typhimurium. Interestingly, day length (mean diurnal range) was only found to be a predictor for Cerro, Mbandaka, and Montevideo. Temperature and precipitation are known parameters influencing *Salmonella* growth and survival in the environment [34,35]. Their generally low contribution estimates in our models may be due to the inclusion of avian flyways and ecoregions which are both proxies for temperature and precipitation information.

Soil variables of importance were cation exchange capacity (I 4,[5],12:i:-, Meleagridis, and Typhimurium), drainage class (Agona, I 4,[5],12:i:-, and Kentucky), organic matter (Agona, Anatum and Montevideo), electrical conductivity (I 4,[5],12:i:- and Infantis), pH (Agona, Anatum, and Dublin,), and texture (Montevideo). Soil geochemical variables of importance were calcium (Agona, Anatum, Cerro, Dublin, Montevideo, Muenchen, Muenster, and Newport), sulfur (Agona, Anatum, Cerro, Montevideo, Muenchen, and Typhimurium), iron (Anatum, I 4,[5],12:i:-, Montevideo, Muenchen, and Muenster), calcium carbonate (I 4,[5],12:i:-, Meleagridis, Montevideo, and Typhimurium), sodium (Anatum and Cerro), zinc (Cerro and Muenchen), phosphorus (Anatum and Montevideo), and copper (Muenchen). These varied responses to soil geochemistry may be related to strain- and isolate-level metabolic capabilities [57] and should be further investigated *in vitro*.

While variance in serovar communities year-to-year is the norm across most BIFSCo regions, Region 3 does not follow this trend. Year-to-year serovar abundances are relatively stable and dominated by Montevideo, Cerro, and Anatum, all non-SoCs. Despite consisting of just three states, Region 3 is the third-highest producer of beef in terms of head slaughtered (68.08 million head from 2014 to 2024), following Regions 6 (73.82) and 5 (97.95). This trend of stability may be due to large number of cattle moving in and out of the region rather than the overall production size. Increased transit of cattle, along with large production sizes, may help to homogenize *Salmonella* serovar communities. More studies on the transportation of cattle and its effect on their *Salmonella* communities could elucidate this trend.

Avian flyways were a dominant variable in eight of our 15 serovar models. Interestingly, those serovars skew non-SoC (6/8 serovars). We hypothesize that while migrating birds are likely contributing to the *Salmonella* detected on beef, they may be contributing serovars and strains which are less pathogenic towards humans.

There appears to be a “trade-off” in variable contributions between avian flyways and ecoregions. Only the models for Muenchen (avian flyways = 25%, ecoregions = 18% variable contribution) and Kentucky (44% and 50% respectively7) incorporate both with similar importance. We hypothesize that this is due to both variables being categorical proxies for each other, in addition to many of the other variables such as climate and soil geochemistry. The mechanistic importance of these variables on the distribution of *Salmonella* serovars is probably much less than that of other variables, and therefore management based directly on flyway or ecoregion membership should be extremely cautious and conservative.

This analysis could be extended to other agriculture systems. While our results demonstrate the incredible diversity of beef-related *Salmonella*, the frameworks of our analysis are easily transferable to other meat commodities and possibly to produce. We found that local animal production facilities can help describe *Salmonella* habitat suitability and we hypothesize this relationship may be bidirectional. Future work could utilize our framework to evaluate the possibility of *Salmonella* “spillover” events between agriculture production systems.

In conclusion, our results demonstrate that certain beef-related serovars occupy different regions through the U.S. and may have differing environmental preferences. Future work should further evaluate and confirm these environmental preferences in laboratory, mesocosm, and observational studies. If spatial trends exhibit persistence across studies, these predictors could become important management factors, for example, increasing feedlot shade to reduce temperatures or incorporating soil amendments to deter *Salmonella* growth and colonization. Our data demonstrate that serovar trends occur on smaller, more local scales, and we suggest that risk prediction should therefore occur on a local scale rather than nationally. However, this incorporates a significant limitation: data are already extremely limited in both spatial and temporal resolutions. Modeling on a local scale, there are likely too few occurrences so as not to produce any meaningful relationships. Therefore, an obvious and important takeaway from this study is that more relevant surveillance sampling is needed if we are to understand and control the occurrence of beef-associated *Salmonella*.

## Acknowledgements

We wish to thank Joanna VanDenBoom for administrative support. The use of product and company names is necessary to accurately report the methods and results; however, the United States Department of Agriculture (USDA) neither guarantees nor warrants the standard of the products, and the use of names by the USDA implies no approval of the product to the exclusion of others that may also be suitable. The USDA is an equal opportunity provider and employer.

## Data Availability Statement

The datasets generated during and/or analysed during the current study are available in the Github repository, https://github.com/tatumskatz/spatialSalmonella.

