## Supplementary File 1 for "Geographic Distributions of Top Beef Salmonella Serovars in the U.S."

### Supplementary File 1. Ecoregions.

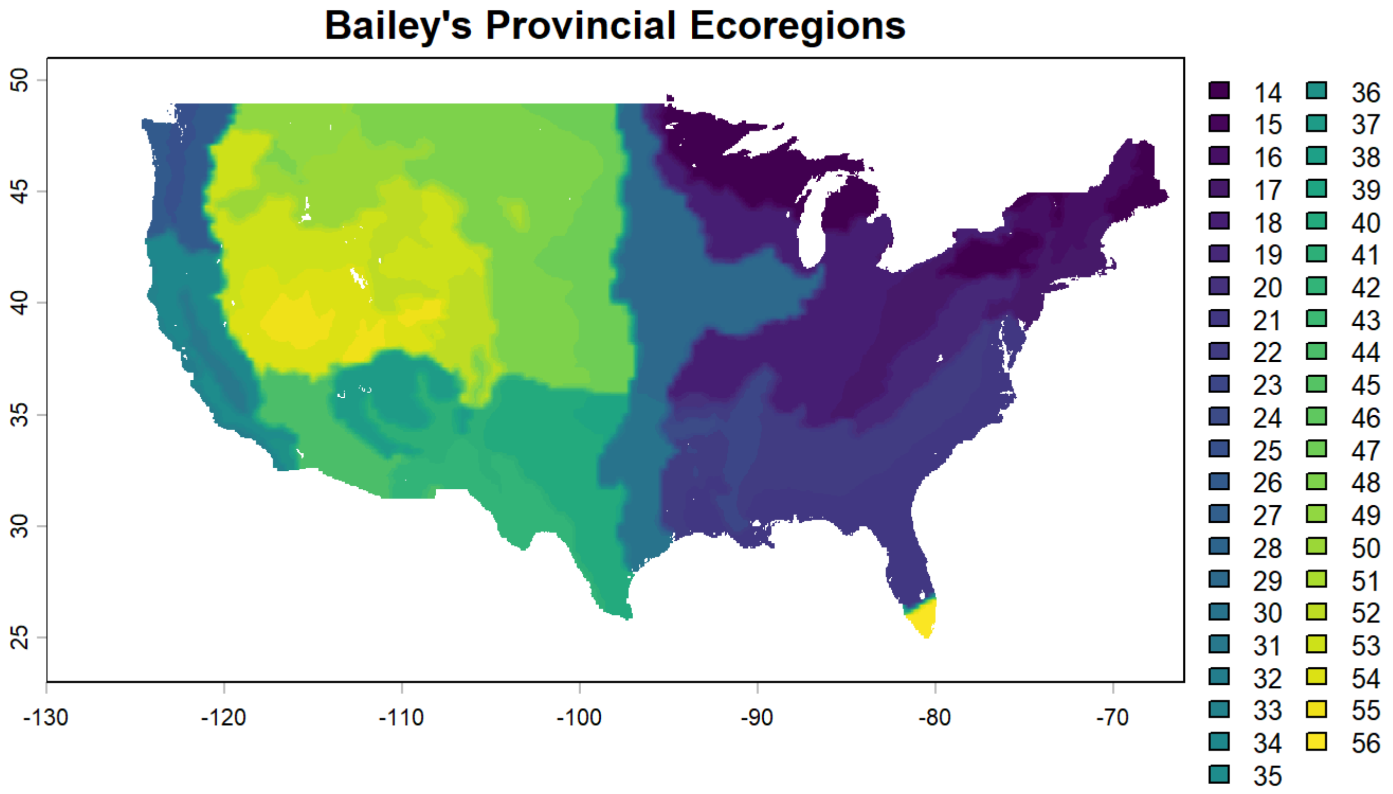


| Index | Ecoregion |
|---|---|
| 14 | Mixed deciduous-coniferous forests (HUMID TEMPERATE DOMAIN, Warm Continental Division) |
| 15 | Mixed forest - coniferous forest - tundra, medium (HUMID TEMPERATE DOMAIN, Warm Continental Mountains) |
| 16 | Mixed forest - coniferous forest - tundra, high (HUMID TEMPERATE DOMAIN, Warm Continental Mountains) |
| 17 | Broadleaved forests, oceanic (HUMID TEMPERATE DOMAIN, Hot Continental Division) |
| 18 | Broadleaved forests, continental (HUMID TEMPERATE DOMAIN, Hot Continental Division) |
| 19 | Deciduous or mixed forest - coniferous forest - meadow (HUMID TEMPERATE DOMAIN, Hot Continental Mountains) |
| 20 | Broadleaf forest - meadow (HUMID TEMPERATE DOMAIN, Hot Continental Mountains) |
| 21 | Broadleaved-coniferous evergreen forests (HUMID TEMPERATE DOMAIN, Subtropical Division) |
| 22 | Coniferous-broadleaved semi-evergreen forests (HUMID TEMPERATE DOMAIN, Subtropical Division) |
| 23 | Riverine forest (HUMID TEMPERATE DOMAIN, Subtropical Division) |
| 24 | Mixed forest - meadow (HUMID TEMPERATE DOMAIN, Subtropical Mountains) |
| 25 | Mixed forests (HUMID TEMPERATE DOMAIN, Marine Division) |
| 26 | Deciduous or mixed forest - coniferous forest - meadow (HUMID TEMPERATE DOMAIN, Marine Mountains) |
| 27 | Forest - meadow, medium (HUMID TEMPERATE DOMAIN, Marine Mountains) |
| 28 | Forest - meadow, high (HUMID TEMPERATE DOMAIN, Marine Mountains) |
| 29 | Forest-steppes and prairies (HUMID TEMPERATE DOMAIN, Prairie Division) |
| 30 | Prairies and savannas (HUMID TEMPERATE DOMAIN, Prairie Division) |

| Index | Ecoregion |
|---|---|
| 31 | Dry steppe (HUMID TEMPERATE DOMAIN, Mediterranean Division) |
| 32 | Mediterranean hardleaved evergreen forests, open woodlands and shrub (HUMID TEMPERATE DOMAIN, Mediterranean Division) |
| 33 | Redwood forests (HUMID TEMPERATE DOMAIN, Mediterranean Division) |
| 34 | Mixed forest - coniferous forest - alpine meadow (HUMID TEMPERATE DOMAIN, Mediterranean Mountains) |
| 35 | Mediterranean woodland or shrub - mixed or coniferous forest - steppe or meadow (HUMID TEMPERATE DOMAIN, Mediterranean Mountains) |
| 36 | Shrub or woodland - steppe - meadow (HUMID TEMPERATE DOMAIN, Mediterranean Mountains) |
| 37 | Coniferous open woodland and semideserts (DRY DOMAIN, Tropical/Subtropical Steppe Division) |
| 38 | Steppes (DRY DOMAIN, Tropical/Subtropical Steppe Division) |
| 39 | Steppes and shrubs (DRY DOMAIN, Tropical/Subtropical Steppe Division) |
| 40 | Shortgrass steppes (DRY DOMAIN, Tropical/Subtropical Steppe Division) |
| 41 | Steppe or semidesert - mixed forest - alpine meadow or steppe (DRY DOMAIN, Tropical/Subtropical Steppe Mountains) |
| 42 | Semideserts (DRY DOMAIN, Tropical/Subtropical Desert Division) |
| 43 | Oceanic Semideserts (DRY DOMAIN, Tropica/Subtropical Desert Division) |
| 44 | Deserts on sand (DRY DOMAIN, Tropical/Subtropical Desert Division) |
| 45 | Semidesert - shrub - open woodland - steppe or alphine meadow (DRY DOMAIN - Tropical/Subtropical Desert Mountains) |
| 46 | Desert or semidesert - open woodland or shrub - desert or steppe (DRY DOMAIN - Tropical/Subtropical Desert Mountains) |
| 47 | Steppes (DRY DOMAIN, Temperate Steppe Division) |
| 48 | Dry steppes (DRY DOMAIN, Temperate Steppe Division) |
| 49 | Forest-steppe - coniferous forest - meadow - tundra (DRY DOMAIN, Temperate Steppe Mountains) |
| 50 | Steppe - coniferous forest - tundra (DRY DOMAIN, Temperate Steppe Mountains) |
| 51 | Steppe - coniferous forest (DRY DOMAIN, Temperate Steppe Mountains) |
| 52 | Steppe - open woodland - coniferous forest - alpine meadow (DRY DOMAIN, Temperate Steppe Mountains) |
| 53 | Semideserts (DRY DOMAIN, Temperate Desert Division) |
| 54 | Semideserts and deserts (DRY DOMAIN, Temperate Desert Division) |
| 55 | Semidesert - open woodland - coniferous forest - alpine meadow (DRY DOMAIN, Temperate Desert Mountains) |
| 56 | Open woodlands, shrubs, and savannas (HUMID TROPICAL DOMAIN, Savanna Division) |
