## Supplementary File 2 for "Geographic Distributions of Top Beef Salmonella Serovars in the U.S."

### Species Distribution Model Results for Serovar I 4,[5],12:i:-

Model tuning results for I 4,[5],12:i:-.

| N Points | N Training Points | N Background Points | Regularization Multiplier | Features | Training AUC |
|---|---|---|---|---|---|
| 26 | 13 | 10012 | 4 | LQPT | 0.79 |

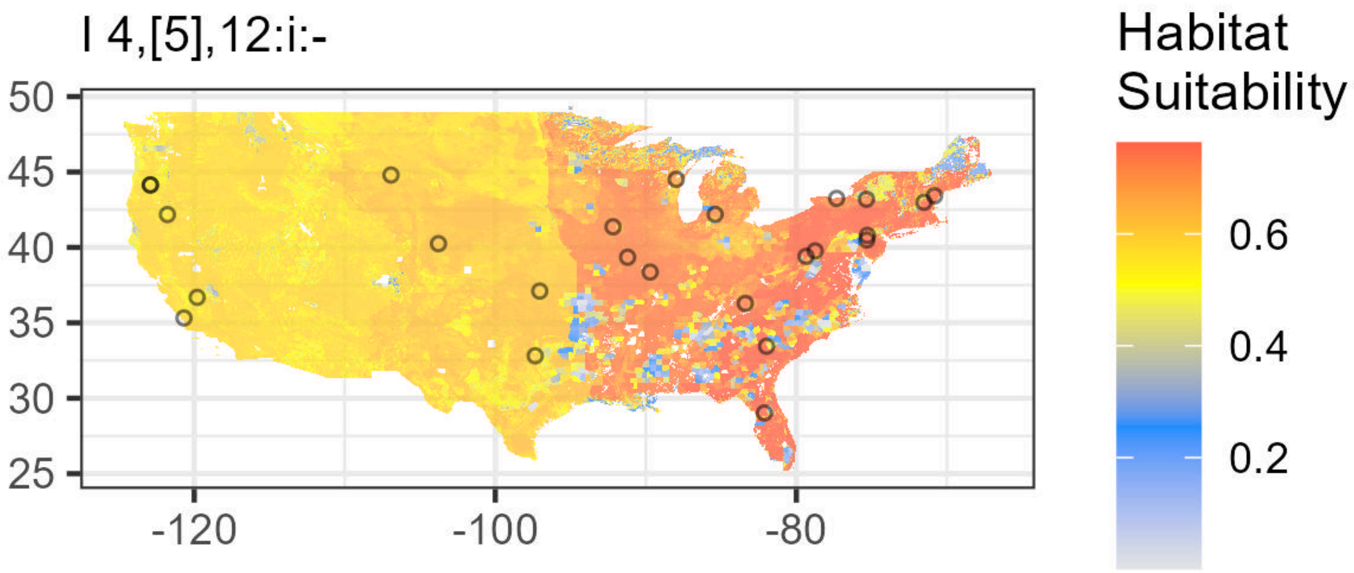

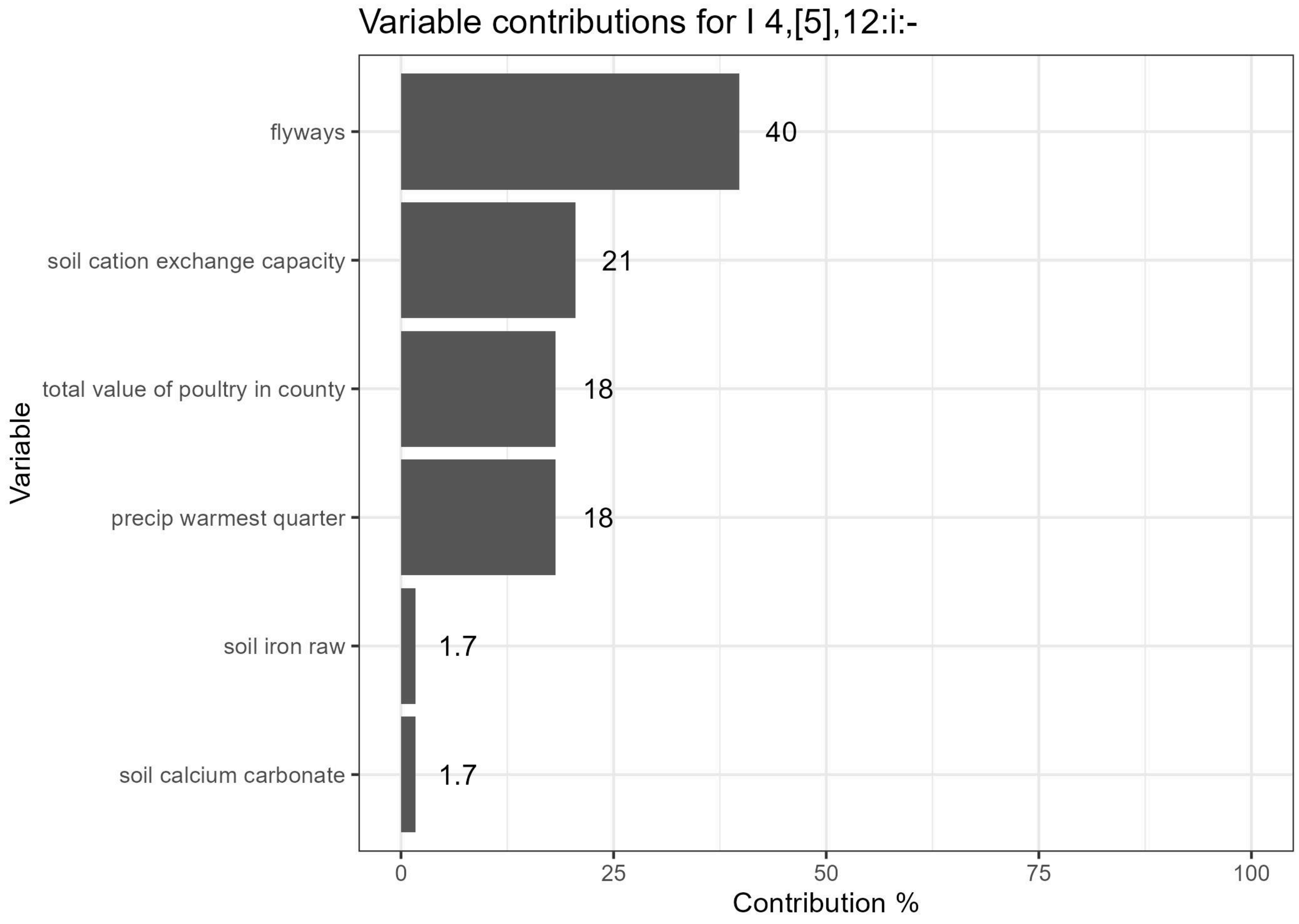


Lambdas for I 4,[5],12:i:-. Flyways are indicated as follows: (1) Atlantic, (2) Central, (3) Mississippi, (4) Pacific. See Supplementary File 1 for Ecoregion values.

| Feature | Variable | Type | Lambda | Minimum Encountered Value | Maximum Encountered Value |
|---|---|---|---|---|---|
| ecoregions==6.0 | ecoregions | categorical | 0.0000000 | 0.0000000 | 1.000000e+00 |
| flyways==1.0 | flyways | categorical | 0.0858532 | 0.0000000 | 1.000000e+00 |
| flyways==2.0 | flyways | categorical | -0.2923380 | 0.0000000 | 1.000000e+00 |
| flyways==4.0 | flyways | categorical | -0.3913029 | 0.0000000 | 1.000000e+00 |
| Top5_Ca | soil calcium raw | linear | 0.0000000 | 0.0000000 | 2.550000e+02 |
| Top5_Ca_percWt | soil calcium as % | linear | 0.0000000 | 0.0602393 | 1.992601e+01 |
| Top5_Cu | soil copper raw | linear | 0.0000000 | 0.0000000 | 2.550000e+02 |
| Top5_Cu_mgPerKg | soil copper mg/kg | linear | 0.0000000 | 2.8719833 | 1.058925e+02 |
| Top5_Fe | soil iron raw | linear | 0.0000000 | 0.0000000 | 2.550000e+02 |
| Top5_Fe_percWt | soil iron % | linear | 0.0000000 | 0.1996400 | 7.030277e+00 |
| Top5_K | soil potassium raw | linear | 0.0000000 | 0.0000000 | 2.550000e+02 |
| Top5_K_percWt | soil potassium as % | linear | 0.0000000 | 0.0594150 | 3.245399e+00 |
| Top5_Mn | soil manganese raw | linear | 0.0000000 | 0.0000000 | 2.550000e+02 |

| Feature | Variable | Type | Lambda | Minimum Encountered Value | Maximum Encountered Value |
|---|---|---|---|---|---|
| Top5_Na | soil sodium raw | linear | 0.0000000 | 0.0000000 | 2.550000e+02 |
| Top5_Na_percWt | soil sodium % | linear | 0.0000000 | 0.0193413 | 2.787977e+00 |
| Top5_P | soil phosphorus raw | linear | 0.0000000 | 0.0000000 | 2.550000e+02 |
| Top5_P_mgPerKg | soil phosphorus mg/kg | linear | 0.0000000 | 129.0937042 | 2.175466e+03 |
| Top5_S | soil sulfur raw | linear | 0.0000000 | 0.0000000 | 2.550000e+02 |
| Top5_S_percWt | soil sulfur as % | linear | 0.0000000 | 0.0065229 | 2.838830e-01 |
| Top5_Zn | soil zinc raw | linear | 0.0000000 | 0.0000000 | 2.550000e+02 |
| Top5_Zn_mgPerKg | soil zinc mg/kg | linear | 0.0000000 | 7.6673822 | 3.405013e+02 |
| caco3_kg_sq_m | soil calcium carbonate | linear | 0.0000000 | 0.0000000 | 1.111125e+03 |
| cec_05 | soil cation exchange capacity | linear | 0.0000000 | 0.1000000 | 2.267935e+02 |
| drainage_class_int | soil drainage class | linear | 0.0000000 | 1.0000000 | 8.000000e+00 |
| ec_05 | soil electrical conductivity | linear | 0.0000000 | 0.0000000 | 1.285708e+02 |
| headCattle_nOperations | n cattle operations in county | linear | 0.0000000 | 4.0000000 | 4.000000e+01 |
| headCattle_totalValue | total value of cattle in county | linear | 0.0000000 | 4.0000000 | 4.838209e+06 |
| headOtherLivestock_nOperations | n other livestock operations in county | linear | 0.0000000 | 2.0000000 | 1.900000e+01 |
| headOtherLivestock_totalValue | total value of other livestock in county | linear | 0.0000000 | 2.0000000 | 3.769605e+06 |
| headPoultry_nOperations | n poultry operations in county | linear | 0.0000000 | 1.0000000 | 1.700000e+01 |
| headPoultry_totalValue | total value of poultry in county | linear | 0.0000000 | 1.0000000 | 3.690649e+07 |
| om_kg_sq_m | soil organic matter | linear | 0.0000000 | 0.1549386 | 4.057721e+02 |
| ph_05 | soil pH | linear | 0.0000000 | 3.5535202 | 9.520809e+00 |
| produce_nOperations | n produce operations in county | linear | 0.0000000 | 0.0000000 | 4.100000e+01 |
| texture_05 | soil texture | linear | 0.0000000 | 1.0000000 | 1.200000e+01 |
| wc2.1_2.5m_bio_10 | mean temp warmest quarter | linear | 0.0000000 | 7.2746668 | 3.568067e+01 |

| Feature | Variable | Type | Lambda | Minimum Encountered Value | Maximum Encountered Value |
|---|---|---|---|---|---|
| wc2.1_2.5m_bio_11 | mean temp coldest quarter | linear | 0.0000000 | -15.1360006 | 2.027333e+01 |
| wc2.1_2.5m_bio_12 | annual precipitation | linear | 0.0000000 | 56.0000000 | 3.265000e+03 |
| wc2.1_2.5m_bio_13 | precip wettest month | linear | 0.0000000 | 10.0000000 | 5.200000e+02 |
| wc2.1_2.5m_bio_14 | precip driest month | linear | 0.0000000 | 0.0000000 | 1.480000e+02 |
| wc2.1_2.5m_bio_15 | precip seasonality | linear | 0.0000000 | 5.6854510 | 9.270832e+01 |
| wc2.1_2.5m_bio_16 | precip wettest quarter | linear | 0.0000000 | 28.0000000 | 1.440000e+03 |
| wc2.1_2.5m_bio_17 | precip driest quarter | linear | 0.0000000 | 2.0000000 | 4.620000e+02 |
| wc2.1_2.5m_bio_18 | precip warmest quarter | linear | 0.0000000 | 2.0000000 | 6.400000e+02 |
| wc2.1_2.5m_bio_19 | precip coldest quarter | linear | 0.0000000 | 15.0000000 | 1.362000e+03 |
| wc2.1_2.5m_bio_1 | annual mean temp | linear | 0.0000000 | -1.7851665 | 2.438933e+01 |
| wc2.1_2.5m_bio_2 | mean diurnal range | linear | 0.0000000 | 6.4996662 | 2.152800e+01 |
| wc2.1_2.5m_bio_3 | isothermality | linear | 0.0000000 | 22.8106346 | 5.848952e+01 |
| wc2.1_2.5m_bio_4 | temperature seasonality | linear | 0.0000000 | 314.8252258 | 1.358003e+03 |
| wc2.1_2.5m_bio_5 | max temp warmest month | linear | 0.0000000 | 15.5039997 | 4.526000e+01 |
| wc2.1_2.5m_bio_6 | min temp coldest month | linear | 0.0000000 | -23.7679996 | 1.528800e+01 |
| wc2.1_2.5m_bio_7 | annual temp range | linear | 0.0000000 | 16.2666664 | 4.986800e+01 |
| wc2.1_2.5m_bio_8 | mean temp wettest quarter | linear | 0.0000000 | -8.7200003 | 3.338200e+01 |
| wc2.1_2.5m_bio_9 | mean temp driest quarter | linear | 0.0000000 | -14.7966671 | 2.854333e+01 |
| cec_05^2 | soil cation exchange capacity | quadratic | -5.3579930 | 0.0100000 | 5.143531e+04 |
| Top5_Fe*caco3_kg_sq_m | soil iron raw,soil calcium carbonate | product | -0.7723855 | 0.0000000 | 2.811267e+05 |
| drainage_class_int*ec_05 | soil drainage class,soil electrical conductivity | product | -0.3850983 | 0.0000000 | 7.220865e+02 |

| Feature | Variable | Type | Lambda | Minimum Encountered Value | Maximum Encountered Value |
|---|---|---|---|---|---|
| headPoultry_totalValue*wc2.1_2.5m_bio_18 | total value of poultry in county,precip warmest quarter | product | -7.1147382 | 24.0000000 | 1.125870e+10 |

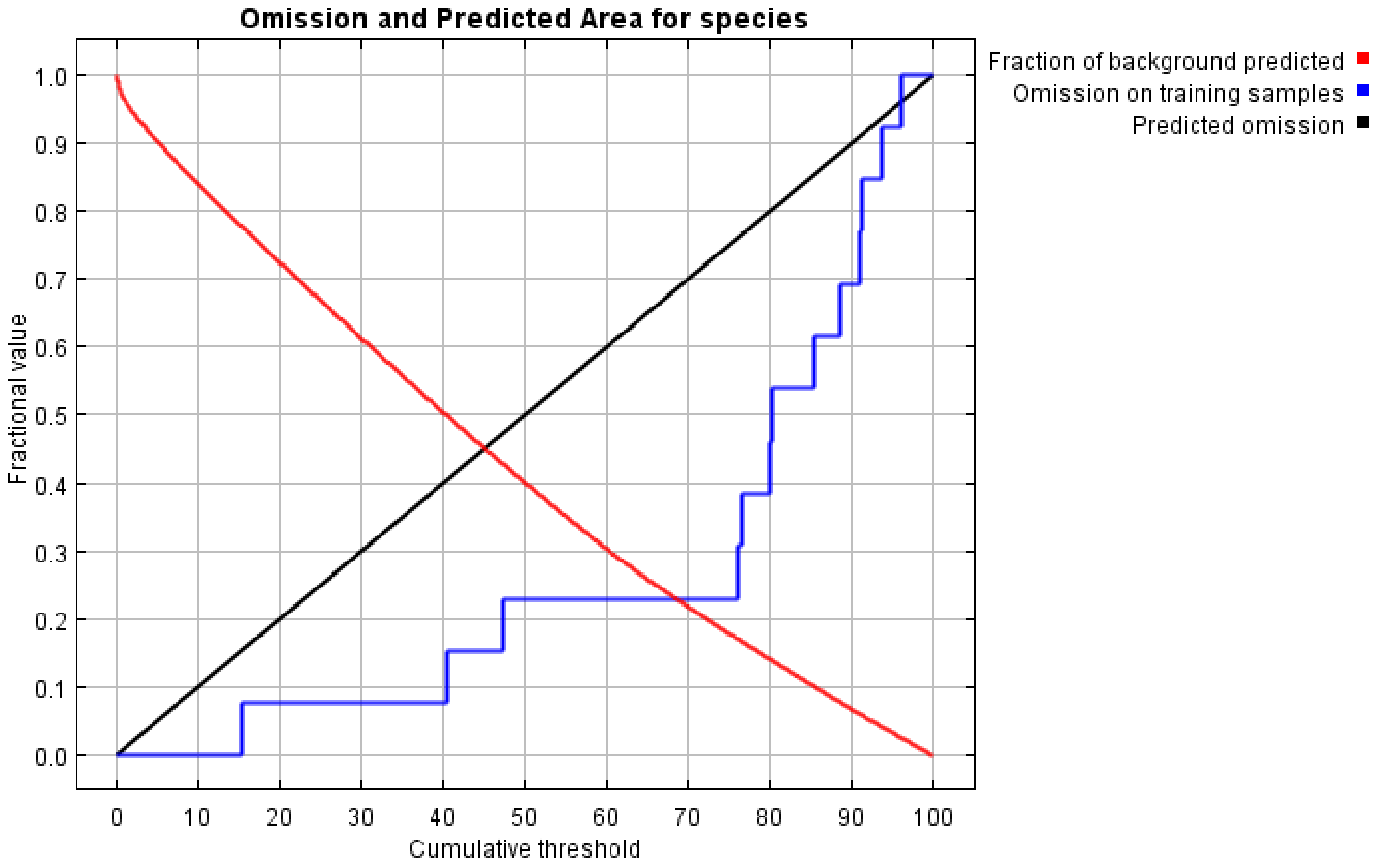

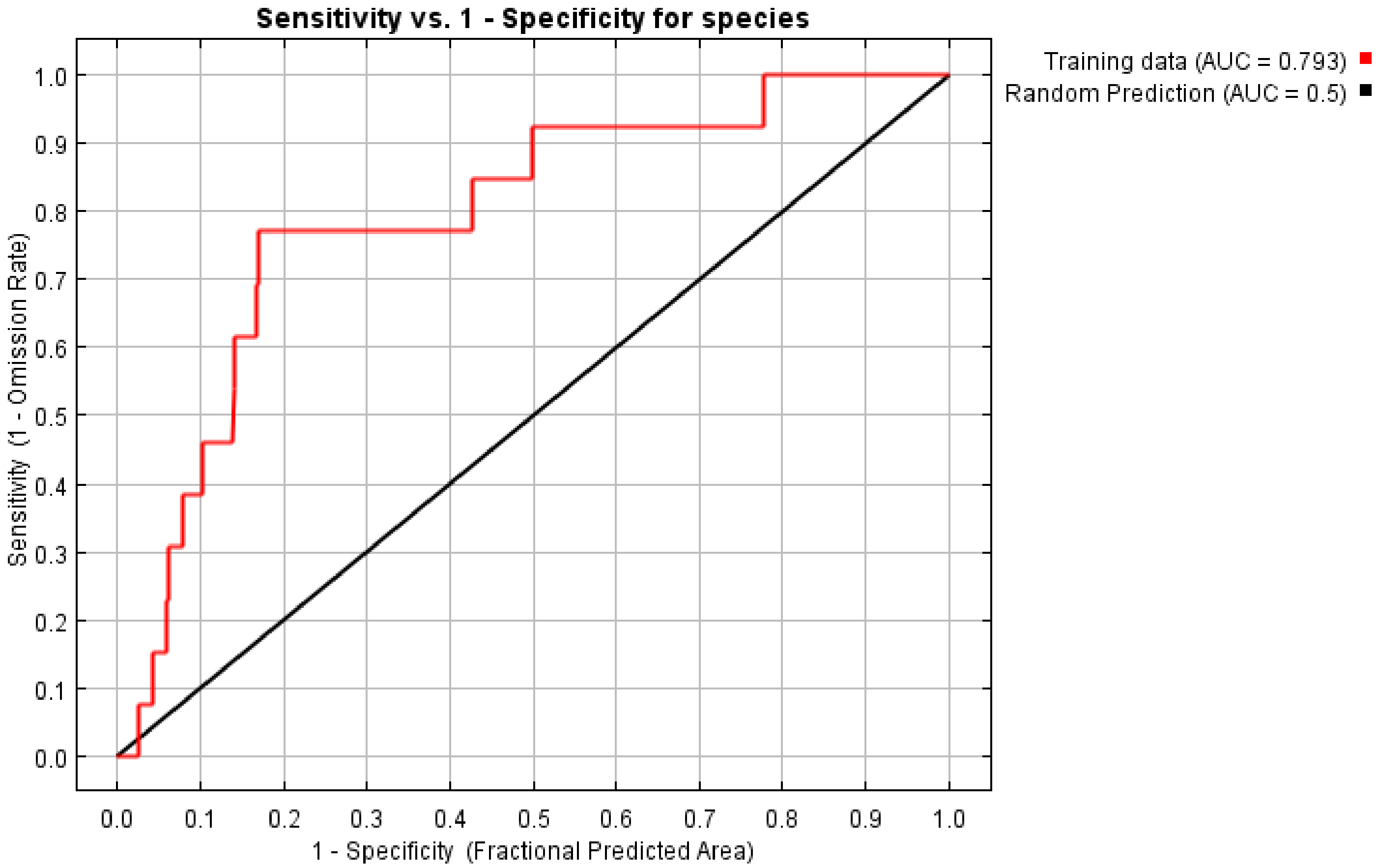
Sensitivity vs. 1 - Specificity for species
Training data (AUC = 0.793)
Random Prediction (AUC = 0.5)
Sensitivity (1 - Omission Rate)
1 - Specificity (Fractional Predicted Area)
0.0
0.1
0.2
0.3
0.4
0.5
0.6
0.7
0.8
0.9
1.0
