## Supplementary File 4 for "Geographic Distributions of Top Beef Salmonella Serovars in the U.S."

### Species Distribution Model Results for Serovar Anatum

Model tuning results for Anatum.

| N Points | N Training Points | N Background Points | Regularization Multiplier | Features | Training AUC |
|---|---|---|---|---|---|
| 143 | 31 | 10028 | 3.5 | LQPT | 0.87 |

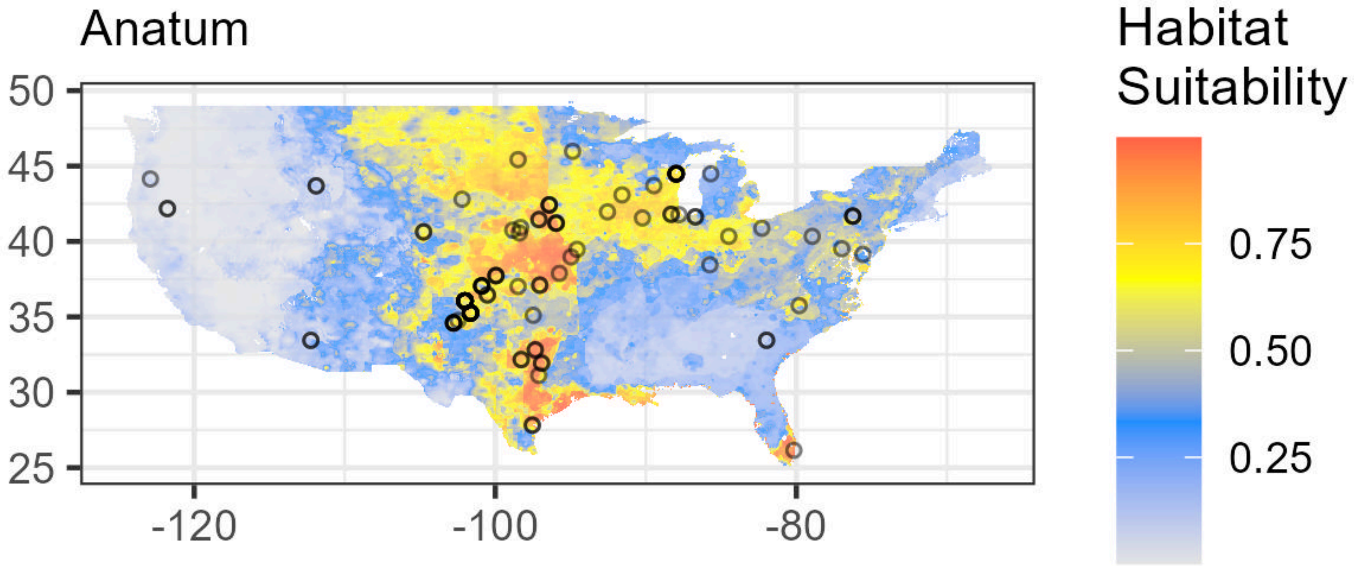

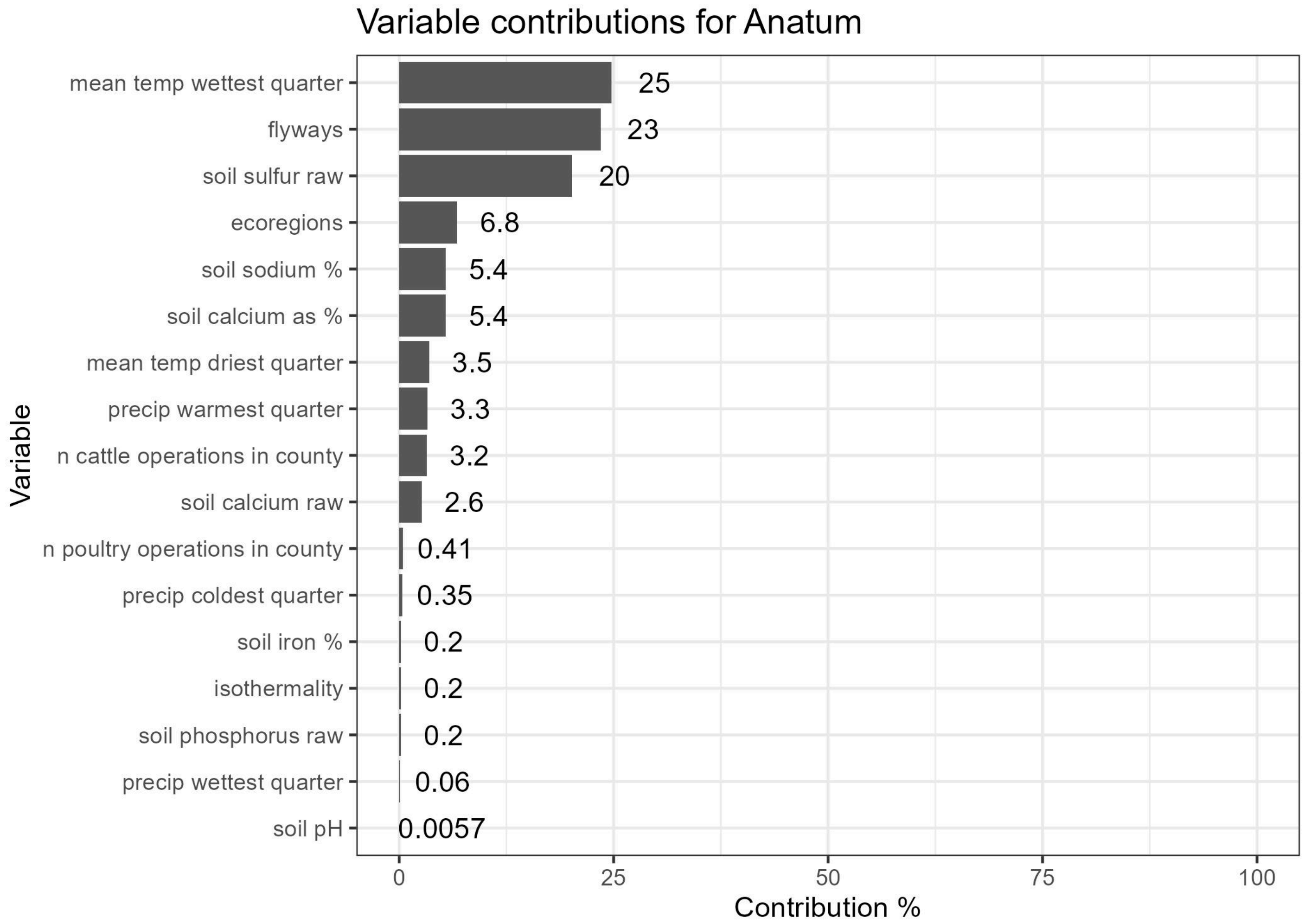


Lambdas for Anatum. Flyways are indicated as follows: (1) Atlantic, (2) Central, (3) Mississippi, (4) Pacific. See Supplementary File 1 for Ecoregion values.

| Feature | Variable | Type | Lambda | Minimum Encountered Value | Maximum Encountered Value |
|---|---|---|---|---|---|
| ecoregions==6.0 | ecoregions | categorical | 0.0000000 | 0.0000000 | 1.000000e+00 |
| ecoregions==10.0 | ecoregions | categorical | 0.1538778 | 0.0000000 | 1.000000e+00 |
| ecoregions==40.0 | ecoregions | categorical | -0.5496806 | 0.0000000 | 1.000000e+00 |
| flyways==1.0 | flyways | categorical | -0.0452216 | 0.0000000 | 1.000000e+00 |
| flyways==2.0 | flyways | categorical | 0.6928406 | 0.0000000 | 1.000000e+00 |
| Top5_Ca | soil calcium raw | linear | 0.0000000 | 0.0000000 | 2.550000e+02 |
| Top5_Ca_percWt | soil calcium as % | linear | 0.0000000 | 0.0697423 | 2.044179e+01 |
| Top5_Cu | soil copper raw | linear | 0.0000000 | 0.0000000 | 2.550000e+02 |
| Top5_Cu_mgPerKg | soil copper mg/kg | linear | 0.0000000 | 3.3159254 | 1.143508e+02 |
| Top5_Fe | soil iron raw | linear | 0.0000000 | 0.0000000 | 2.550000e+02 |
| Top5_Fe_percWt | soil iron % | linear | 0.0000000 | 0.1342737 | 6.475775e+00 |
| Top5_K | soil potassium raw | linear | 0.0000000 | 0.0000000 | 2.550000e+02 |
| Top5_K_percWt | soil potassium as % | linear | 0.0000000 | 0.0550254 | 3.334370e+00 |
| Top5_Mn | soil manganese raw | linear | 0.0000000 | 0.0000000 | 2.550000e+02 |

| Feature | Variable | Type | Lambda | Minimum Encountered Value | Maximum Encountered Value |
|---|---|---|---|---|---|
| Top5_Na | soil sodium raw | linear | 0.0000000 | 0.0000000 | 2.550000e+02 |
| Top5_Na_percWt | soil sodium % | linear | 0.0000000 | 0.0228538 | 2.957286e+00 |
| Top5_P | soil phosphorus raw | linear | 0.0000000 | 0.0000000 | 2.550000e+02 |
| Top5_P_mgPerKg | soil phosphorus mg/kg | linear | 0.0000000 | 96.5034027 | 2.095814e+03 |
| Top5_S | soil sulfur raw | linear | 0.0000000 | 0.0000000 | 2.550000e+02 |
| Top5_S_percWt | soil sulfur as % | linear | 0.0000000 | 0.0064985 | 4.163159e-01 |
| Top5_Zn | soil zinc raw | linear | 0.0000000 | 0.0000000 | 2.550000e+02 |
| Top5_Zn_mgPerKg | soil zinc mg/kg | linear | 0.0000000 | 6.1639123 | 3.657585e+02 |
| caco3_kg_sq_m | soil calcium carbonate | linear | 0.0000000 | 0.0000000 | 1.102458e+03 |
| cec_05 | soil cation exchange capacity | linear | 0.0000000 | 0.3875331 | 1.965865e+02 |
| drainage_class_int | soil drainage class | linear | 0.0000000 | 1.0000000 | 8.000000e+00 |
| ec_05 | soil electrical conductivity | linear | 0.0000000 | 0.0000000 | 1.400000e+02 |
| headCattle_nOperations | n cattle operations in county | linear | 0.0000000 | 4.0000000 | 4.000000e+01 |
| headCattle_totalValue | total value of cattle in county | linear | 0.0000000 | 4.0000000 | 4.838209e+06 |
| headOtherLivestock_nOperations | n other livestock operations in county | linear | 0.0000000 | 2.0000000 | 1.900000e+01 |
| headOtherLivestock_totalValue | total value of other livestock in county | linear | 0.0000000 | 2.0000000 | 3.769605e+06 |
| headPoultry_nOperations | n poultry operations in county | linear | 0.0000000 | 1.0000000 | 1.700000e+01 |
| headPoultry_totalValue | total value of poultry in county | linear | 0.0000000 | 1.0000000 | 3.690649e+07 |
| om_kg_sq_m | soil organic matter | linear | 0.0000000 | 0.0112372 | 3.741486e+02 |
| ph_05 | soil pH | linear | 0.0000000 | 3.2691472 | 9.588049e+00 |
| produce_nOperations | n produce operations in county | linear | 0.0000000 | 0.0000000 | 4.100000e+01 |
| texture_05 | soil texture | linear | 0.0000000 | 1.0000000 | 1.200000e+01 |
| wc2.1_2.5m_bio_10 | mean temp warmest quarter | linear | 0.0000000 | 6.6120000 | 3.623200e+01 |

| Feature | Variable | Type | Lambda | Minimum Encountered Value | Maximum Encountered Value |
|---|---|---|---|---|---|
| wc2.1_2.5m_bio_11 | mean temp coldest quarter | linear | 0.0000000 | -15.1859999 | 2.051600e+01 |
| wc2.1_2.5m_bio_12 | annual precipitation | linear | 0.0000000 | 56.0000000 | 3.329000e+03 |
| wc2.1_2.5m_bio_13 | precip wettest month | linear | 0.0000000 | 10.0000000 | 5.420000e+02 |
| wc2.1_2.5m_bio_14 | precip driest month | linear | 0.0000000 | 0.0000000 | 1.360000e+02 |
| wc2.1_2.5m_bio_15 | precip seasonality | linear | 0.0000000 | 5.9053688 | 9.344160e+01 |
| wc2.1_2.5m_bio_16 | precip wettest quarter | linear | 0.0000000 | 27.0000000 | 1.488000e+03 |
| wc2.1_2.5m_bio_17 | precip driest quarter | linear | 0.0000000 | 2.0000000 | 4.310000e+02 |
| wc2.1_2.5m_bio_18 | precip warmest quarter | linear | 0.0000000 | 2.0000000 | 6.370000e+02 |
| wc2.1_2.5m_bio_19 | precip coldest quarter | linear | 0.0000000 | 16.0000000 | 1.416000e+03 |
| wc2.1_2.5m_bio_1 | annual mean temp | linear | 0.0000000 | -2.3285000 | 2.463317e+01 |
| wc2.1_2.5m_bio_2 | mean diurnal range | linear | 0.0000000 | 6.0920005 | 2.155233e+01 |
| wc2.1_2.5m_bio_3 | isothermality | linear | 0.0000000 | 23.5659561 | 6.171973e+01 |
| wc2.1_2.5m_bio_4 | temperature seasonality | linear | 0.0000000 | 246.9219360 | 1.357718e+03 |
| wc2.1_2.5m_bio_5 | max temp warmest month | linear | 0.0000000 | 14.0360003 | 4.595600e+01 |
| wc2.1_2.5m_bio_6 | min temp coldest month | linear | 0.0000000 | -24.0760002 | 1.531200e+01 |
| wc2.1_2.5m_bio_7 | annual temp range | linear | 0.0000000 | 16.5388889 | 4.973600e+01 |
| wc2.1_2.5m_bio_8 | mean temp wettest quarter | linear | 0.0000000 | -8.6806669 | 3.331667e+01 |
| wc2.1_2.5m_bio_9 | mean temp driest quarter | linear | 0.0000000 | -14.4813328 | 2.880467e+01 |
| wc2.1_2.5m_bio_19^2 | precip coldest quarter | quadratic | -4.1922472 | 256.0000000 | 2.005056e+06 |
| wc2.1_2.5m_bio_9^2 | mean temp driest quarter | quadratic | -0.6403255 | 0.0000071 | 8.297088e+02 |
| Top5_Ca*headPoultry_nOperations | soil calcium raw,n poultry operations in county | product | 0.5510366 | 0.0000000 | 4.335000e+03 |
| Top5_Ca*wc2.1_2.5m_bio_16 | soil calcium raw,precip wettest quarter | product | 2.2033700 | 0.0000000 | 3.493500e+05 |

| Feature | Variable | Type | Lambda | Minimum Encountered Value | Maximum Encountered Value |
|---|---|---|---|---|---|
| Top5_Ca_percWt*Top5_Na_percWt | soil calcium as %,soil sodium % | product | -5.7278595 | 0.0017054 | 1.173110e+01 |
| Top5_Fe_percWt*wc2.1_2.5m_bio_3 | soil iron %,isothermality | product | -0.5430348 | 6.6346901 | 2.916144e+02 |
| Top5_Na_percWt*wc2.1_2.5m_bio_19 | soil sodium %,precip coldest quarter | product | -1.0340212 | 4.0908321 | 1.993141e+03 |
| Top5_P*wc2.1_2.5m_bio_8 | soil phosphorus raw,mean temp wettest quarter | product | 0.6519095 | -2213.5700655 | 8.449680e+03 |
| Top5_S*wc2.1_2.5m_bio_8 | soil sulfur raw,mean temp wettest quarter | product | 1.8550731 | -2213.5700655 | 8.354650e+03 |
| headCattle_totalValue*wc2.1_2.5m_bio_14 | total value of cattle in county,precip driest month | product | 0.2075549 | 0.0000000 | 5.114264e+07 |
| ph_05*wc2.1_2.5m_bio_8 | soil pH,mean temp wettest quarter | product | 1.4428110 | -61.5996741 | 2.774830e+02 |

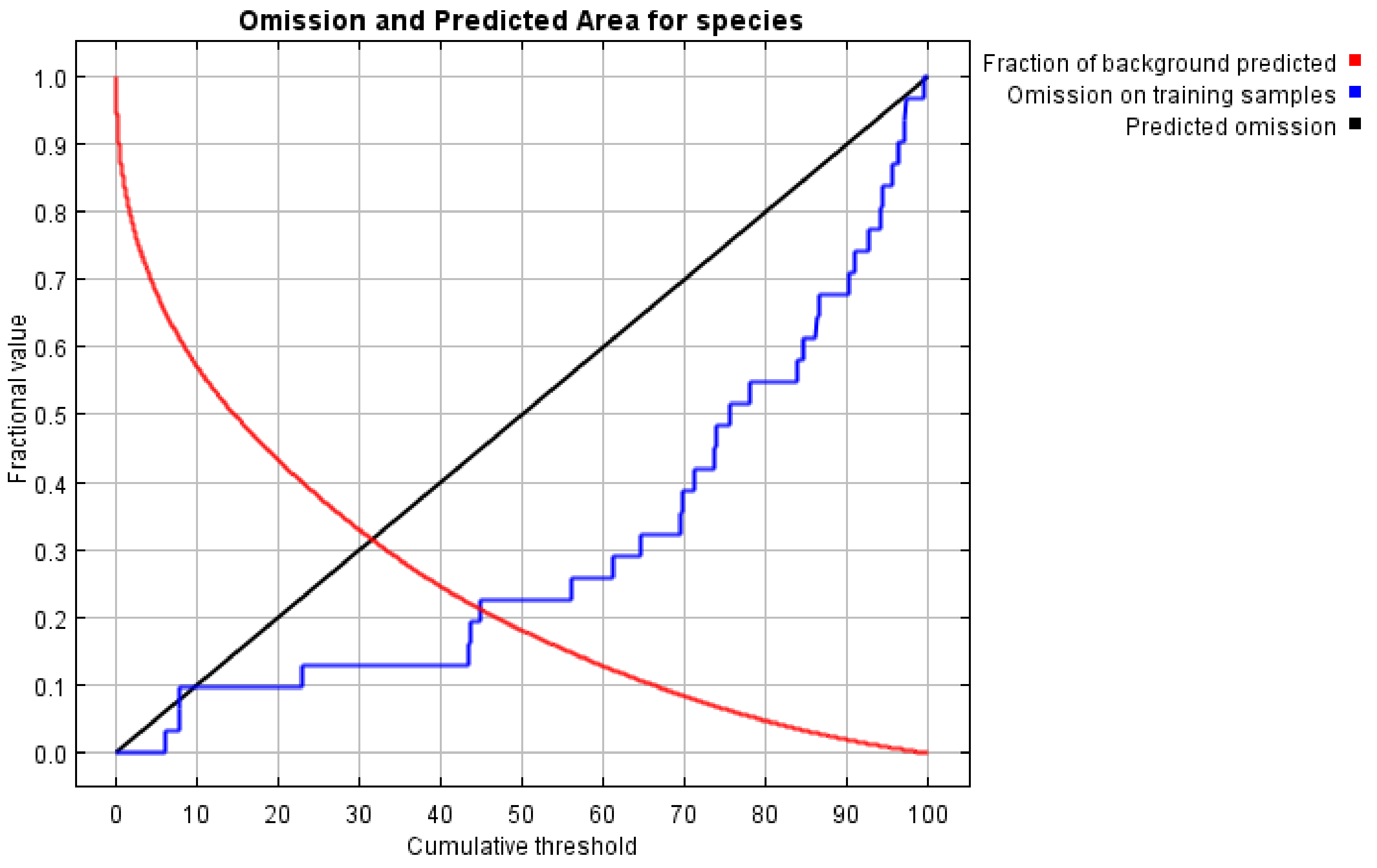

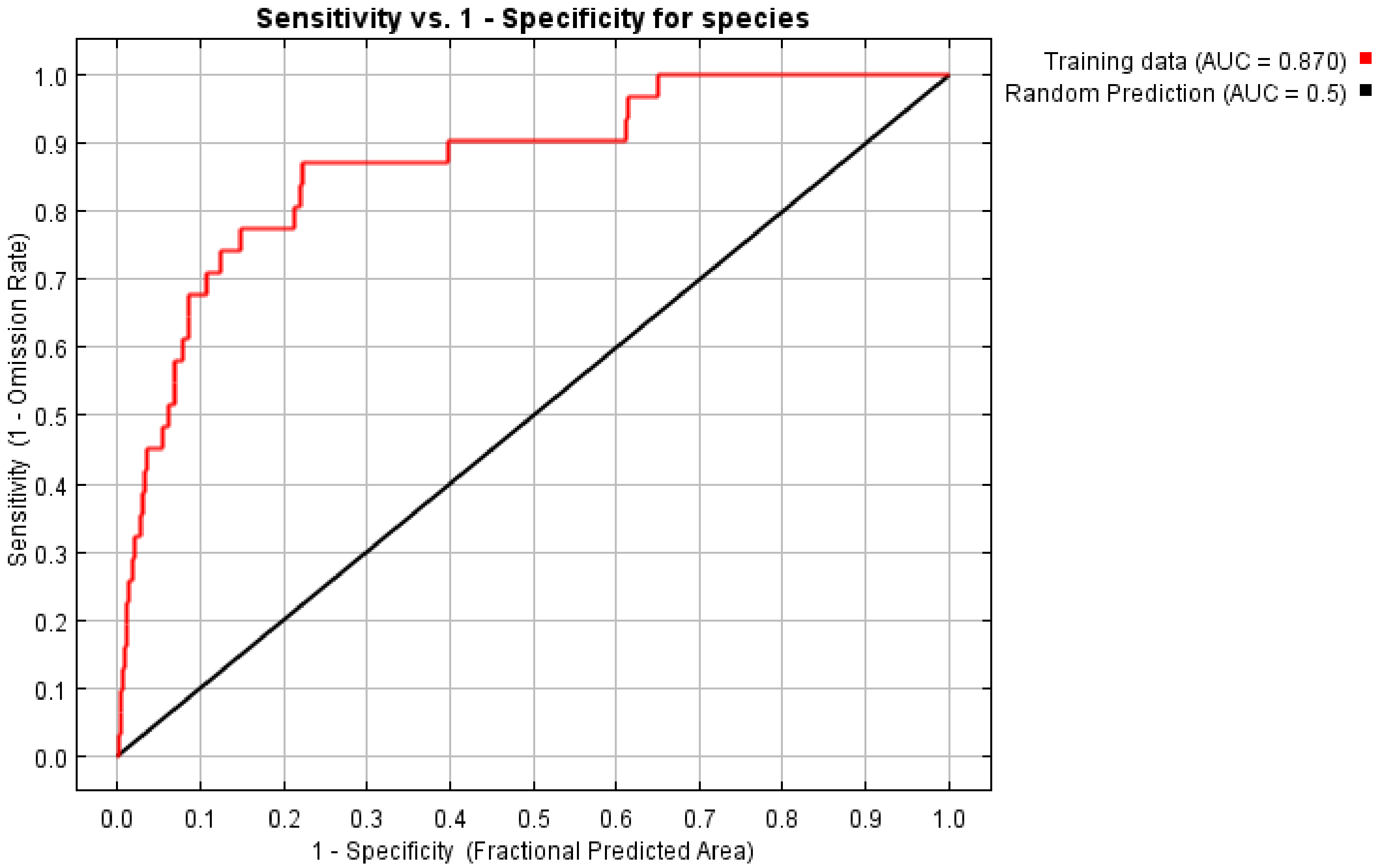
Sensitivity vs. 1 - Specificity for species
Training data (AUC = 0.870)
Random Prediction (AUC = 0.5)
Sensitivity (1 - Omission Rate)
1 - Specificity (Fractional Predicted Area)
0.0
0.1
0.2
0.3
0.4
0.5
0.6
0.7
0.8
0.9
1.0
