## Supplementary File 5 for "Geographic Distributions of Top Beef Salmonella Serovars in the U.S."

### Species Distribution Model Results for Serovar Cerro

Model tuning results for Cerro.

| N Points | N Training Points | N Background Points | Regularization Multiplier | Features | Training AUC |
|---|---|---|---|---|---|
| 59 | 15 | 10015 | 4 | LQPT | 0.86 |

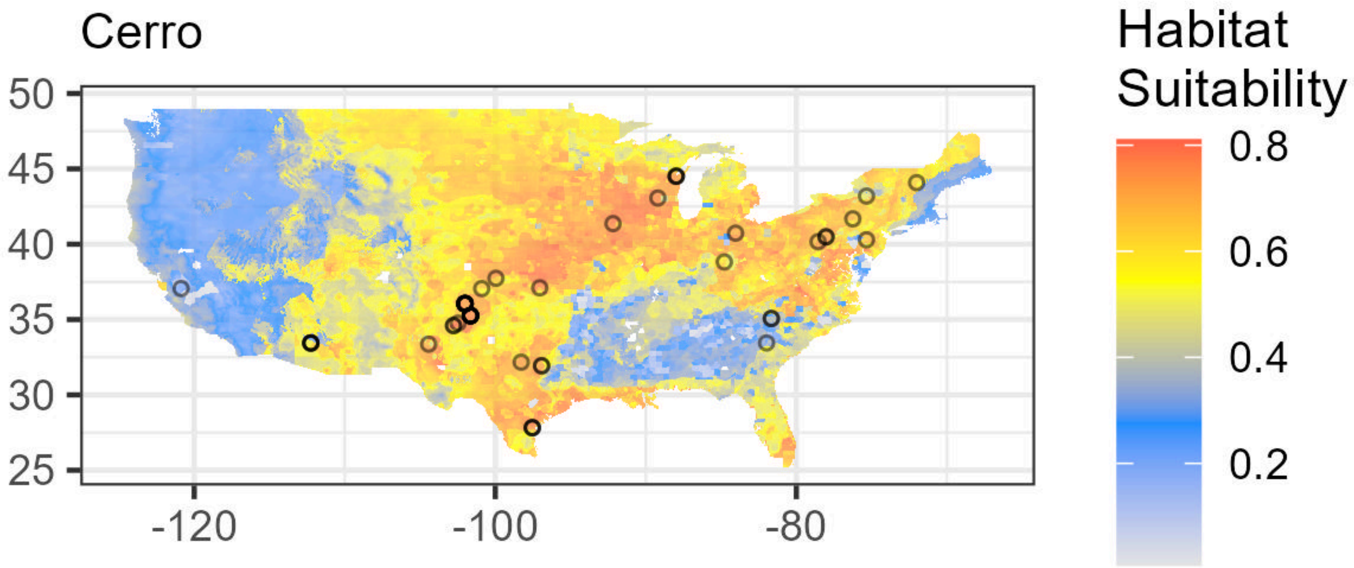

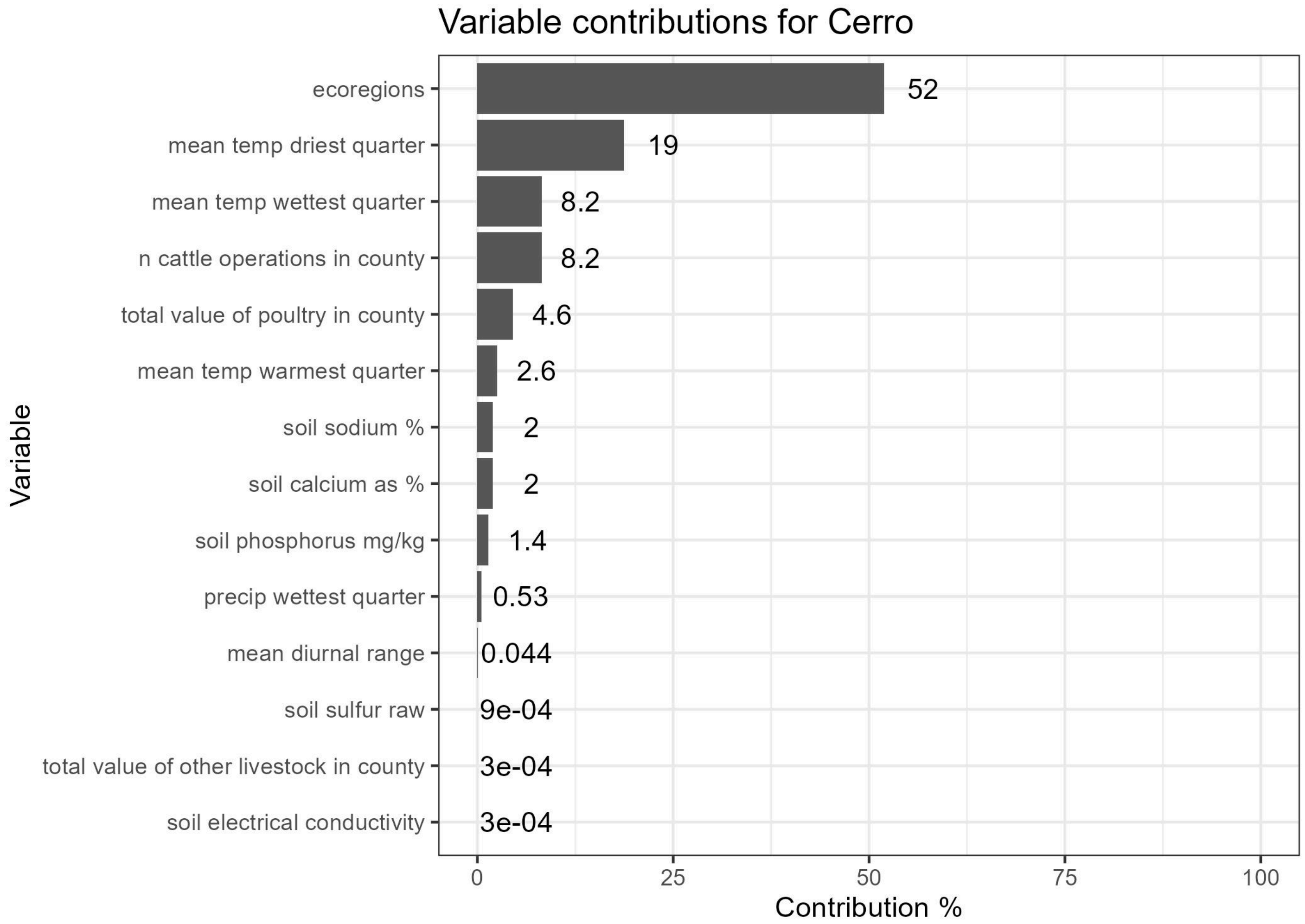


Lambdas for Cerro. Flyways are indicated as follows: (1) Atlantic, (2) Central, (3) Mississippi, (4) Pacific. See Supplementary File 1 for Ecoregion values.

| Feature | Variable | Type | Lambda | Minimum Encountered Value | Maximum Encountered Value |
|---|---|---|---|---|---|
| ecoregions==6.0 | ecoregions | categorical | 0.0000000 | 0.0000000 | 1.000000e+00 |
| ecoregions==32.0 | ecoregions | categorical | 0.7870413 | 0.0000000 | 1.000000e+00 |
| flyways==1.0 | flyways | categorical | 0.0000000 | 0.0000000 | 1.000000e+00 |
| Top5_Ca | soil calcium raw | linear | 0.0000000 | 0.0000000 | 2.550000e+02 |
| Top5_Ca_percWt | soil calcium as % | linear | 0.0000000 | 0.0553820 | 2.106154e+01 |
| Top5_Cu | soil copper raw | linear | 0.0000000 | 0.0000000 | 2.550000e+02 |
| Top5_Cu_mgPerKg | soil copper mg/kg | linear | 0.0000000 | 2.9955404 | 9.188074e+01 |
| Top5_Fe | soil iron raw | linear | 0.0000000 | 0.0000000 | 2.550000e+02 |
| Top5_Fe_percWt | soil iron % | linear | 0.0000000 | 0.1367162 | 7.471953e+00 |
| Top5_K | soil potassium raw | linear | 0.0000000 | 0.0000000 | 2.550000e+02 |
| Top5_K_percWt | soil potassium as % | linear | 0.0000000 | 0.0506226 | 3.412038e+00 |
| Top5_Mn | soil manganese raw | linear | 0.0000000 | 0.0000000 | 2.550000e+02 |

| Feature | Variable | Type | Lambda | Minimum Encountered Value | Maximum Encountered Value |
|---|---|---|---|---|---|
| Top5_Na | soil sodium raw | linear | 0.0000000 | 0.0000000 | 2.550000e+02 |
| Top5_Na_percWt | soil sodium % | linear | 0.0000000 | 0.0213182 | 2.882550e+00 |
| Top5_P | soil phosphorus raw | linear | 0.0000000 | 0.0000000 | 2.550000e+02 |
| Top5_P_mgPerKg | soil phosphorus mg/kg | linear | 0.0000000 | 120.0265503 | 2.351274e+03 |
| Top5_S | soil sulfur raw | linear | 0.0000000 | 0.0000000 | 2.550000e+02 |
| Top5_S_percWt | soil sulfur as % | linear | 0.0000000 | 0.0072871 | 3.637684e-01 |
| Top5_Zn | soil zinc raw | linear | 0.0000000 | 0.0000000 | 2.550000e+02 |
| Top5_Zn_mgPerKg | soil zinc mg/kg | linear | 0.0000000 | 7.0200615 | 3.635258e+02 |
| caco3_kg_sq_m | soil calcium carbonate | linear | 0.0000000 | 0.0000000 | 9.922277e+02 |
| cec_05 | soil cation exchange capacity | linear | 0.0000000 | 0.6000000 | 1.992000e+02 |
| drainage_class_int | soil drainage class | linear | 0.0000000 | 1.0000000 | 8.000000e+00 |
| ec_05 | soil electrical conductivity | linear | 0.0000000 | 0.0000000 | 1.531205e+02 |
| headCattle_nOperations | n cattle operations in county | linear | 0.0000000 | 4.0000000 | 4.000000e+01 |
| headCattle_totalValue | total value of cattle in county | linear | 0.0000000 | 4.0000000 | 4.838209e+06 |
| headOtherLivestock_nOperations | n other livestock operations in county | linear | 0.0000000 | 2.0000000 | 1.900000e+01 |
| headOtherLivestock_totalValue | total value of other livestock in county | linear | 0.0000000 | 2.0000000 | 3.769605e+06 |
| headPoultry_nOperations | n poultry operations in county | linear | 0.0000000 | 1.0000000 | 1.700000e+01 |
| headPoultry_totalValue | total value of poultry in county | linear | 0.0000000 | 1.0000000 | 3.690649e+07 |
| om_kg_sq_m | soil organic matter | linear | 0.0000000 | 0.0069909 | 4.859369e+02 |
| ph_05 | soil pH | linear | 0.0000000 | 3.2685268 | 9.654279e+00 |
| produce_nOperations | n produce operations in county | linear | 0.0000000 | 0.0000000 | 4.100000e+01 |
| texture_05 | soil texture | linear | 0.0000000 | 1.0000000 | 1.200000e+01 |
| wc2.1_2.5m_bio_10 | mean temp warmest quarter | linear | 0.0000000 | 6.4179997 | 3.607733e+01 |

| Feature | Variable | Type | Lambda | Minimum Encountered Value | Maximum Encountered Value |
|---|---|---|---|---|---|
| wc2.1_2.5m_bio_11 | mean temp coldest quarter | linear | 0.0000000 | -14.7220001 | 2.047500e+01 |
| wc2.1_2.5m_bio_12 | annual precipitation | linear | 0.0000000 | 58.0000000 | 3.336000e+03 |
| wc2.1_2.5m_bio_13 | precip wettest month | linear | 0.0000000 | 10.0000000 | 5.370000e+02 |
| wc2.1_2.5m_bio_14 | precip driest month | linear | 0.0000000 | 0.0000000 | 1.430000e+02 |
| wc2.1_2.5m_bio_15 | precip seasonality | linear | 0.0000000 | 5.4618278 | 9.483382e+01 |
| wc2.1_2.5m_bio_16 | precip wettest quarter | linear | 0.0000000 | 28.0000000 | 1.468000e+03 |
| wc2.1_2.5m_bio_17 | precip driest quarter | linear | 0.0000000 | 2.0000000 | 4.520000e+02 |
| wc2.1_2.5m_bio_18 | precip warmest quarter | linear | 0.0000000 | 2.0000000 | 6.310000e+02 |
| wc2.1_2.5m_bio_19 | precip coldest quarter | linear | 0.0000000 | 16.0000000 | 1.402000e+03 |
| wc2.1_2.5m_bio_1 | annual mean temp | linear | 0.0000000 | -2.7510002 | 2.452500e+01 |
| wc2.1_2.5m_bio_2 | mean diurnal range | linear | 0.0000000 | 6.7567463 | 2.100733e+01 |
| wc2.1_2.5m_bio_3 | isothermality | linear | 0.0000000 | 23.1432858 | 6.282427e+01 |
| wc2.1_2.5m_bio_4 | temperature seasonality | linear | 0.0000000 | 252.4221802 | 1.353021e+03 |
| wc2.1_2.5m_bio_5 | max temp warmest month | linear | 0.0000000 | 14.8160000 | 4.558000e+01 |
| wc2.1_2.5m_bio_6 | min temp coldest month | linear | 0.0000000 | -23.5559998 | 1.565833e+01 |
| wc2.1_2.5m_bio_7 | annual temp range | linear | 0.0000000 | 16.3479996 | 5.032800e+01 |
| wc2.1_2.5m_bio_8 | mean temp wettest quarter | linear | 0.0000000 | -10.4660006 | 3.358866e+01 |
| wc2.1_2.5m_bio_9 | mean temp driest quarter | linear | 0.0000000 | -14.7220001 | 2.863067e+01 |
| wc2.1_2.5m_bio_9^2 | mean temp driest quarter | quadratic | -0.4431901 | 0.0000000 | 8.197151e+02 |
| Top5_Ca_percWt*Top5_Na_percWt | soil calcium as %,soil sodium % | product | -1.5593251 | 0.0017536 | 2.175538e+01 |
| Top5_S*wc2.1_2.5m_bio_8 | soil sulfur raw,mean temp wettest quarter | product | 1.0949638 | -2668.8301420 | 8.354650e+03 |

| Feature | Variable | Type | Lambda | Minimum Encountered Value | Maximum Encountered Value |
|---|---|---|---|---|---|
| Top5_Zn_mgPerKg*headPoultry_totalValue | soil zinc mg/kg,total value of poultry in county | product | -1.5789068 | 22.2770386 | 1.795466e+09 |
| headCattle_nOperations*wc2.1_2.5m_bio_8 | n cattle operations in county,mean temp wettest quarter | product | 1.4466574 | -313.9800167 | 1.094302e+03 |
| headPoultry_totalValue*wc2.1_2.5m_bio_16 | total value of poultry in county,precip wettest quarter | product | -1.1978414 | 39.0000000 | 1.177317e+10 |
| headPoultry_totalValue*wc2.1_2.5m_bio_2 | total value of poultry in county,mean diurnal range | product | -2.1280375 | 11.5773335 | 4.242032e+08 |

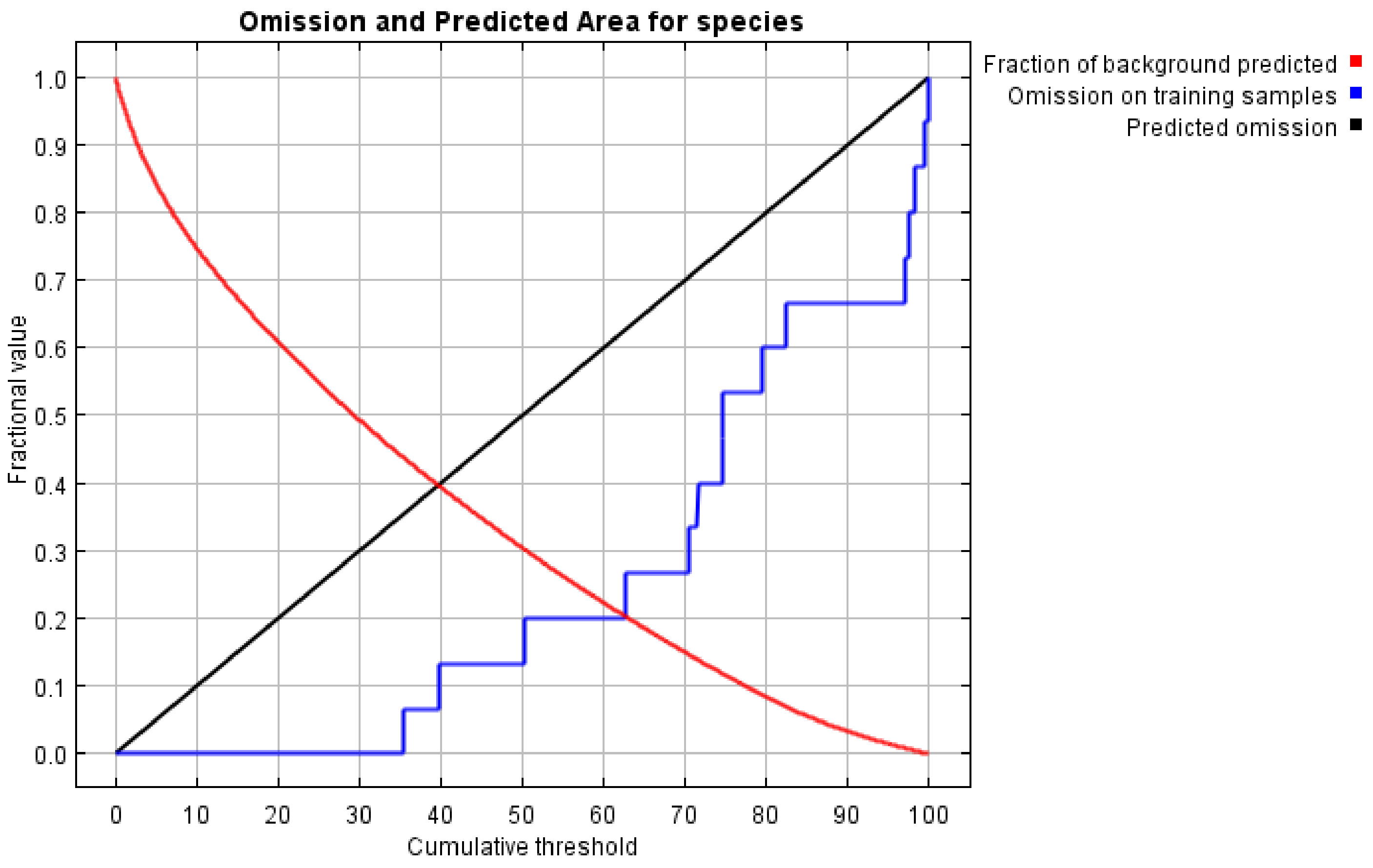

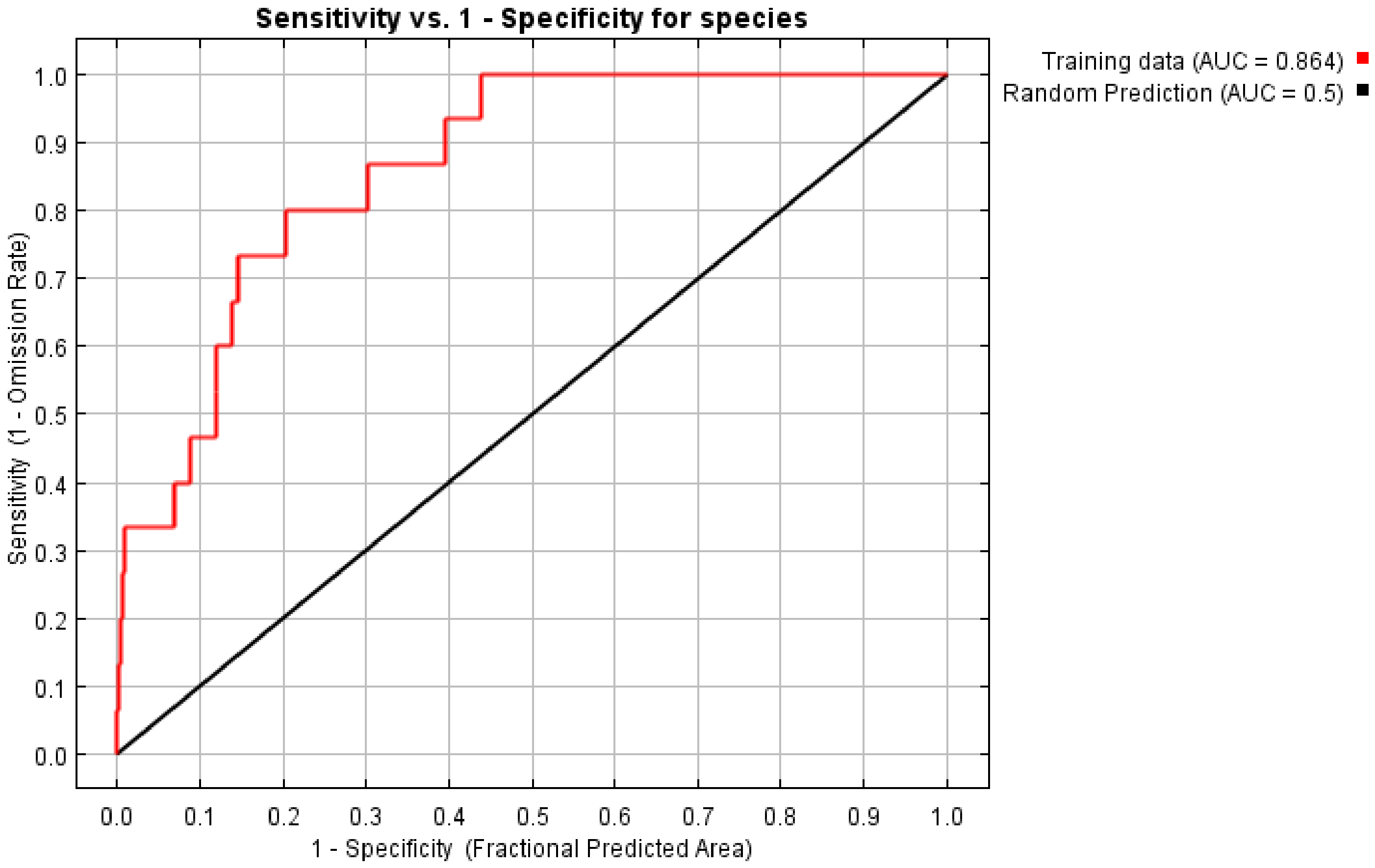
Sensitivity vs. 1 - Specificity for species
Training data (AUC = 0.864)
Random Prediction (AUC = 0.5)
Sensitivity (1 - Omission Rate)
1 - Specificity (Fractional Predicted Area)
0.0
0.1
0.2
0.3
0.4
0.5
0.6
0.7
0.8
0.9
1.0
