## Supplementary File 6 for "Geographic Distributions of Top Beef Salmonella Serovars in the U.S."

### Species Distribution Model Results for Serovar Dublin

Model tuning results for Dublin.

| N Points | N Training Points | N Background Points | Regularization Multiplier | Features | Training AUC |
|---|---|---|---|---|---|
| 81 | 19 | 10019 | 4 | LQPT | 0.89 |

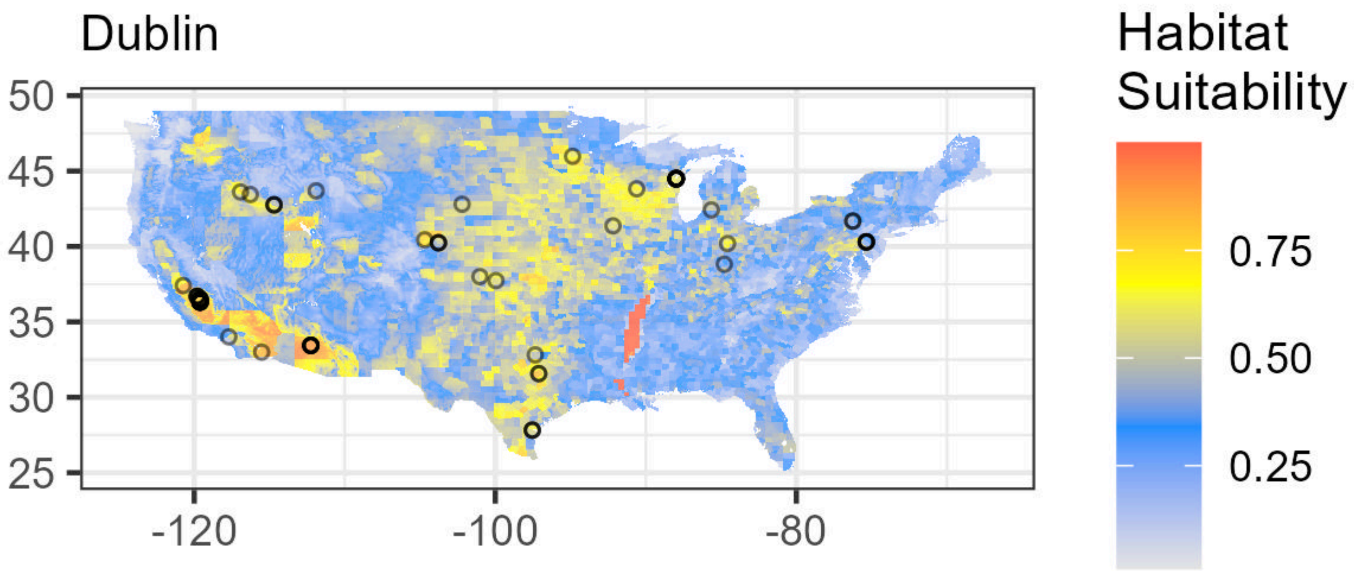

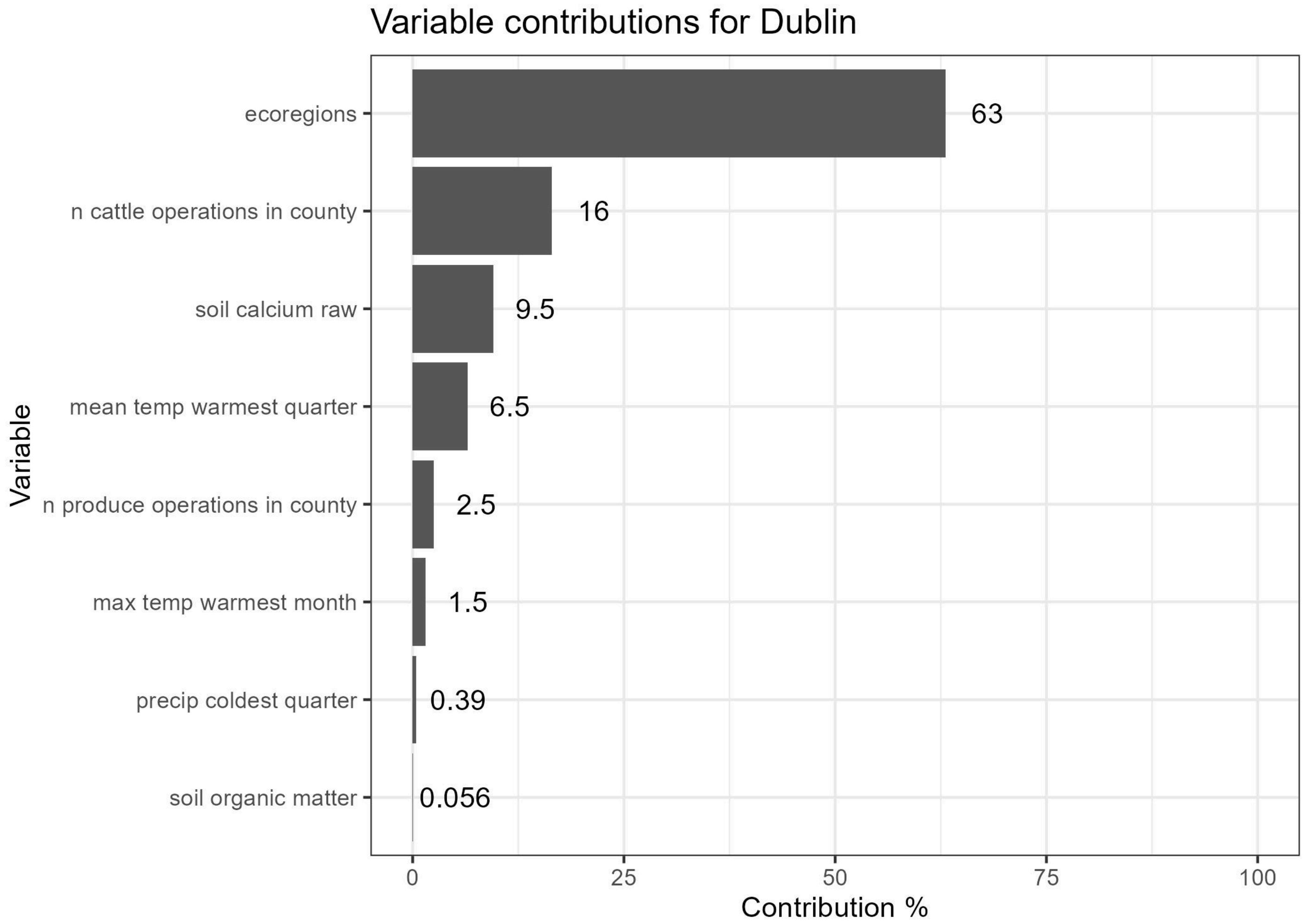


Lambdas for Dublin. Flyways are indicated as follows: (1) Atlantic, (2) Central, (3) Mississippi, (4) Pacific. See Supplementary File 1 for Ecoregion values.

| Feature | Variable | Type | Lambda | Minimum Encountered Value | Maximum Encountered Value |
|---|---|---|---|---|---|
| ecoregions==6.0 | ecoregions | categorical | 0.0000000 | 0.0000000 | 1.000000e+00 |
| ecoregions==10.0 | ecoregions | categorical | 0.5152606 | 0.0000000 | 1.000000e+00 |
| ecoregions==23.0 | ecoregions | categorical | 3.5379705 | 0.0000000 | 1.000000e+00 |
| ecoregions==45.0 | ecoregions | categorical | 0.3915766 | 0.0000000 | 1.000000e+00 |
| flyways==1.0 | flyways | categorical | 0.0000000 | 0.0000000 | 1.000000e+00 |
| Top5_Ca | soil calcium raw | linear | 0.0000000 | 0.0000000 | 2.550000e+02 |
| Top5_Ca_percWt | soil calcium as % | linear | 0.0000000 | 0.0194371 | 1.826883e+01 |
| Top5_Cu | soil copper raw | linear | 0.0000000 | 0.0000000 | 2.550000e+02 |
| Top5_Cu_mgPerKg | soil copper mg/kg | linear | 0.0000000 | 3.6292992 | 9.056213e+01 |
| Top5_Fe | soil iron raw | linear | 0.0000000 | 0.0000000 | 2.550000e+02 |
| Top5_Fe_percWt | soil iron % | linear | 0.0000000 | 0.1798414 | 6.788994e+00 |
| Top5_K | soil potassium raw | linear | 0.0000000 | 0.0000000 | 2.550000e+02 |
| Top5_K_percWt | soil potassium as % | linear | 0.0000000 | 0.0802344 | 3.222574e+00 |

| Feature | Variable | Type | Lambda | Minimum Encountered Value | Maximum Encountered Value |
|---|---|---|---|---|---|
| Top5_Mn | soil manganese raw | linear | 0.0000000 | 0.0000000 | 2.550000e+02 |
| Top5_Na | soil sodium raw | linear | 0.0000000 | 0.0000000 | 2.550000e+02 |
| Top5_Na_percWt | soil sodium % | linear | 0.0000000 | 0.0119298 | 2.820359e+00 |
| Top5_P | soil phosphorus raw | linear | 0.0000000 | 0.0000000 | 2.550000e+02 |
| Top5_P_mgPerKg | soil phosphorus mg/kg | linear | 0.0000000 | 113.9330063 | 2.500958e+03 |
| Top5_S | soil sulfur raw | linear | 0.0000000 | 0.0000000 | 2.550000e+02 |
| Top5_S_percWt | soil sulfur as % | linear | 0.0000000 | 0.0060473 | 4.363081e-01 |
| Top5_Zn | soil zinc raw | linear | 0.0000000 | 0.0000000 | 2.550000e+02 |
| Top5_Zn_mgPerKg | soil zinc mg/kg | linear | 0.0000000 | 7.9354844 | 3.937248e+02 |
| caco3_kg_sq_m | soil calcium carbonate | linear | 0.0000000 | 0.0000000 | 9.757744e+02 |
| cec_05 | soil cation exchange capacity | linear | 0.0000000 | 0.3869249 | 1.992000e+02 |
| drainage_class_int | soil drainage class | linear | 0.0000000 | 1.0000000 | 8.000000e+00 |
| ec_05 | soil electrical conductivity | linear | 0.0000000 | 0.0000000 | 1.400000e+02 |
| headCattle_nOperations | n cattle operations in county | linear | 0.0000000 | 4.0000000 | 4.000000e+01 |
| headCattle_totalValue | total value of cattle in county | linear | 0.0000000 | 4.0000000 | 4.838209e+06 |
| headOtherLivestock_nOperations | n other livestock operations in county | linear | 0.0000000 | 2.0000000 | 1.900000e+01 |
| headOtherLivestock_totalValue | total value of other livestock in county | linear | 0.0000000 | 2.0000000 | 3.769605e+06 |
| headPoultry_nOperations | n poultry operations in county | linear | 0.0000000 | 1.0000000 | 1.700000e+01 |
| headPoultry_totalValue | total value of poultry in county | linear | 0.0000000 | 1.0000000 | 3.690649e+07 |
| om_kg_sq_m | soil organic matter | linear | 0.0000000 | 0.0069909 | 2.976631e+02 |
| ph_05 | soil pH | linear | 0.0000000 | 3.2129552 | 9.694615e+00 |
| produce_nOperations | n produce operations in county | linear | 0.0000000 | 0.0000000 | 4.100000e+01 |
| texture_05 | soil texture | linear | 0.0000000 | 1.0000000 | 1.200000e+01 |

| Feature | Variable | Type | Lambda | Minimum Encountered Value | Maximum Encountered Value |
|---|---|---|---|---|---|
| wc2.1_2.5m_bio_10 | mean temp warmest quarter | linear | 0.0000000 | 6.1120000 | 3.607000e+01 |
| wc2.1_2.5m_bio_11 | mean temp coldest quarter | linear | 0.0000000 | -15.0900002 | 2.031400e+01 |
| wc2.1_2.5m_bio_12 | annual precipitation | linear | 0.0000000 | 56.0000000 | 3.217000e+03 |
| wc2.1_2.5m_bio_13 | precip wettest month | linear | 0.0000000 | 10.0000000 | 5.200000e+02 |
| wc2.1_2.5m_bio_14 | precip driest month | linear | 0.0000000 | 0.0000000 | 1.400000e+02 |
| wc2.1_2.5m_bio_15 | precip seasonality | linear | 0.0000000 | 5.4197531 | 9.387823e+01 |
| wc2.1_2.5m_bio_16 | precip wettest quarter | linear | 0.0000000 | 27.0000000 | 1.443000e+03 |
| wc2.1_2.5m_bio_17 | precip driest quarter | linear | 0.0000000 | 2.0000000 | 4.460000e+02 |
| wc2.1_2.5m_bio_18 | precip warmest quarter | linear | 0.0000000 | 2.0000000 | 6.410000e+02 |
| wc2.1_2.5m_bio_19 | precip coldest quarter | linear | 0.0000000 | 15.0000000 | 1.364000e+03 |
| wc2.1_2.5m_bio_1 | annual mean temp | linear | 0.0000000 | -2.7531667 | 2.445617e+01 |
| wc2.1_2.5m_bio_2 | mean diurnal range | linear | 0.0000000 | 6.4311590 | 2.155233e+01 |
| wc2.1_2.5m_bio_3 | isothermality | linear | 0.0000000 | 22.0201797 | 6.062571e+01 |
| wc2.1_2.5m_bio_4 | temperature seasonality | linear | 0.0000000 | 237.4199066 | 1.358314e+03 |
| wc2.1_2.5m_bio_5 | max temp warmest month | linear | 0.0000000 | 14.4320002 | 4.569600e+01 |
| wc2.1_2.5m_bio_6 | min temp coldest month | linear | 0.0000000 | -23.7399998 | 1.532000e+01 |
| wc2.1_2.5m_bio_7 | annual temp range | linear | 0.0000000 | 16.1652184 | 4.947600e+01 |
| wc2.1_2.5m_bio_8 | mean temp wettest quarter | linear | 0.0000000 | -8.4413338 | 3.334333e+01 |
| wc2.1_2.5m_bio_9 | mean temp driest quarter | linear | 0.0000000 | -14.7206669 | 2.868333e+01 |
| wc2.1_2.5m_bio_19^2 | precip coldest quarter | quadratic | -2.1738276 | 225.0000000 | 1.860496e+06 |
| Top5_Ca*produce_nOperations | soil calcium raw,n produce operations in county | product | 0.8135990 | 0.0000000 | 1.045500e+04 |

| Feature | Variable | Type | Lambda | Minimum Encountered Value | Maximum Encountered Value |
|---|---|---|---|---|---|
| headCattle_nOperations*ph_05 | n cattle operations in county,soil pH | product | 0.3461713 | 16.0647762 | 3.437218e+02 |
| headCattle_nOperations*produce_nOperations | n cattle operations in county,n produce operations in county | product | 0.0967749 | 0.0000000 | 1.400000e+03 |
| headCattle_nOperations*wc2.1_2.5m_bio_10 | n cattle operations in county,mean temp warmest quarter | product | 3.2848076 | 64.0400009 | 1.114696e+03 |

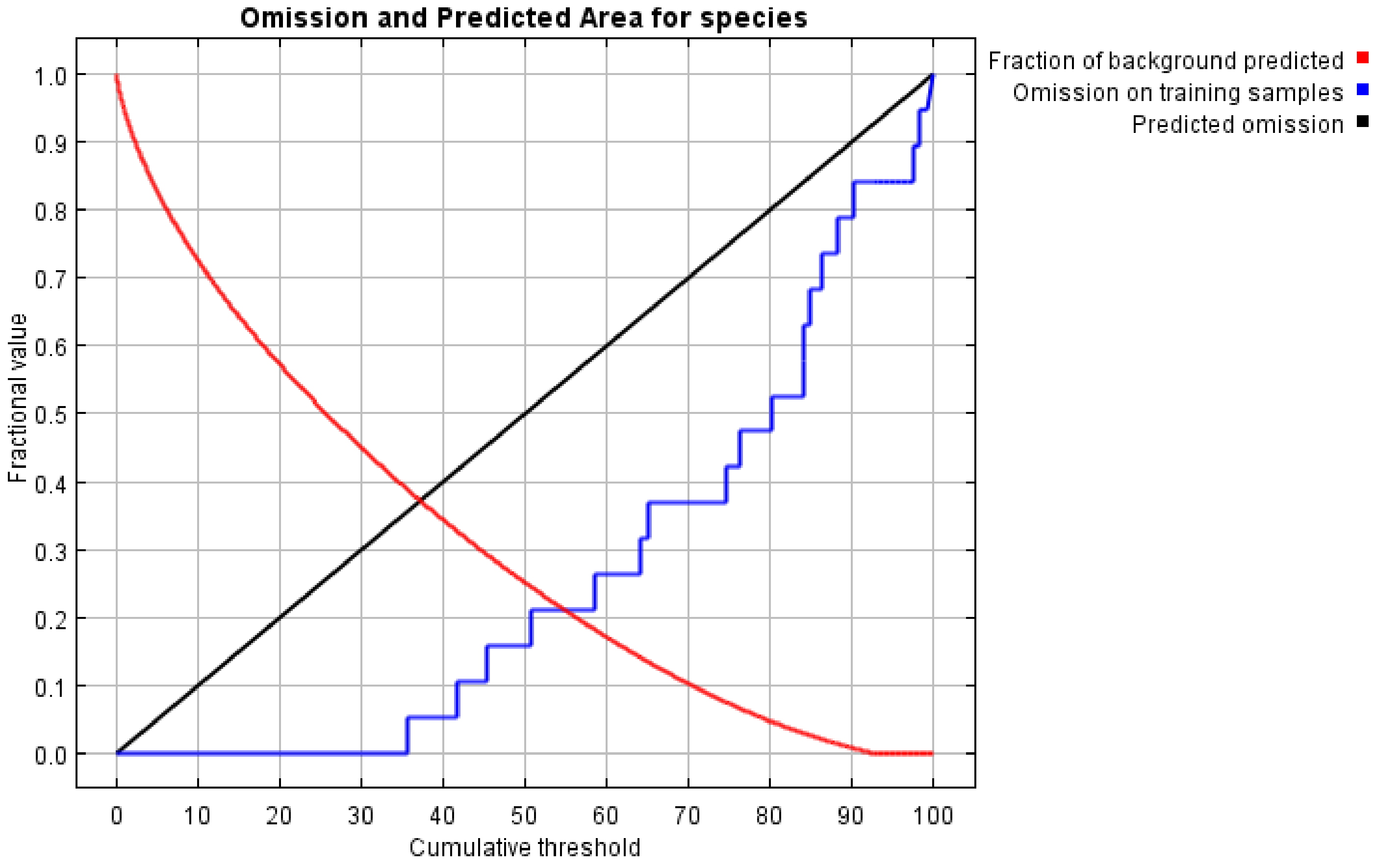

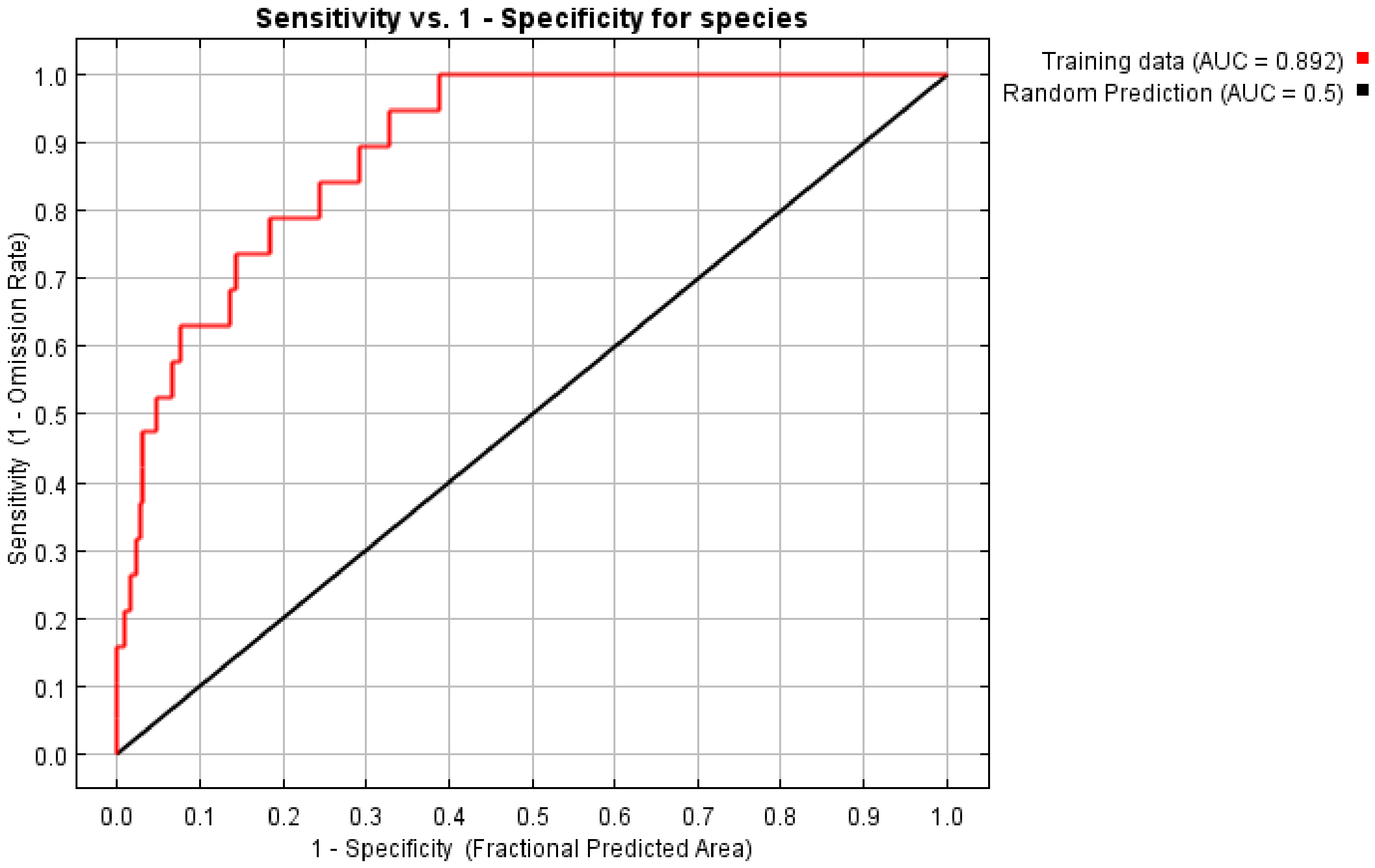

Sensitivity vs. 1 - Specificity for species
Training data (AUC = 0.892)
Random Prediction (AUC = 0.5)
Sensitivity (1 - Omission Rate)
1 - Specificity (Fractional Predicted Area)
0.0
0.1
0.2
0.3
0.4
0.5
0.6
0.7
0.8
0.9
1.0
