## Supplementary File 8 for "Geographic Distributions of Top Beef Salmonella Serovars in the U.S."

### Species Distribution Model Results for Serovar Infantis

Model tuning results for Infantis.

| N Points | N Training Points | N Background Points | Regularization Multiplier | Features | Training AUC |
|---|---|---|---|---|---|
| 34 | 14 | 10014 | 3 | LQPT | 0.83 |

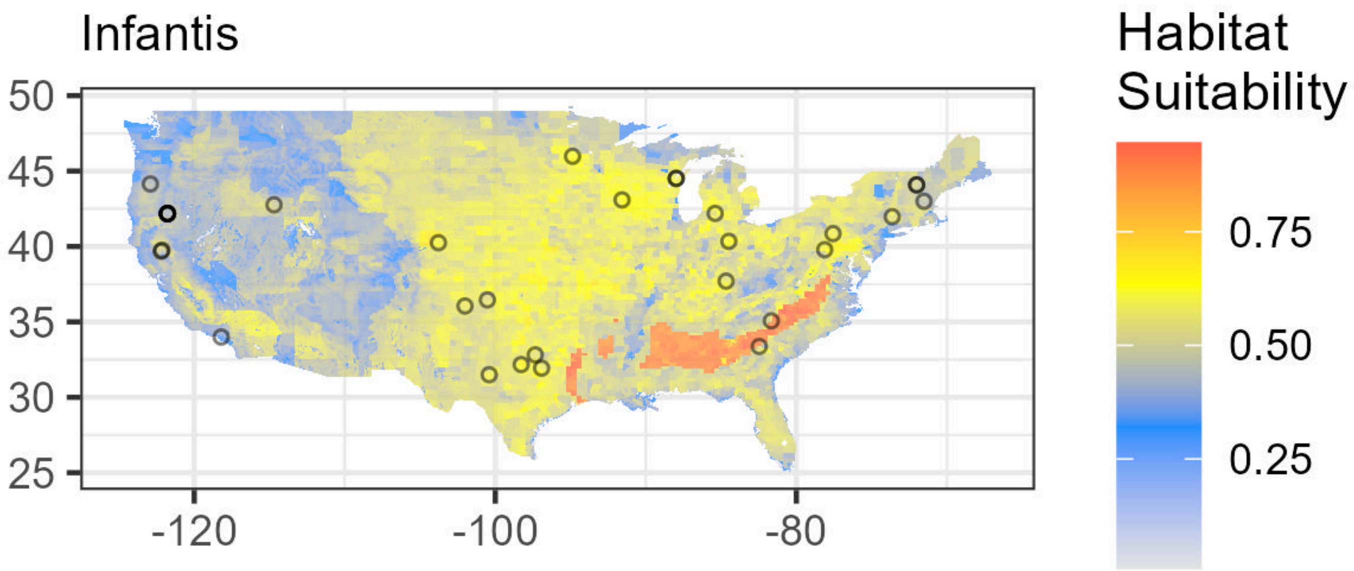

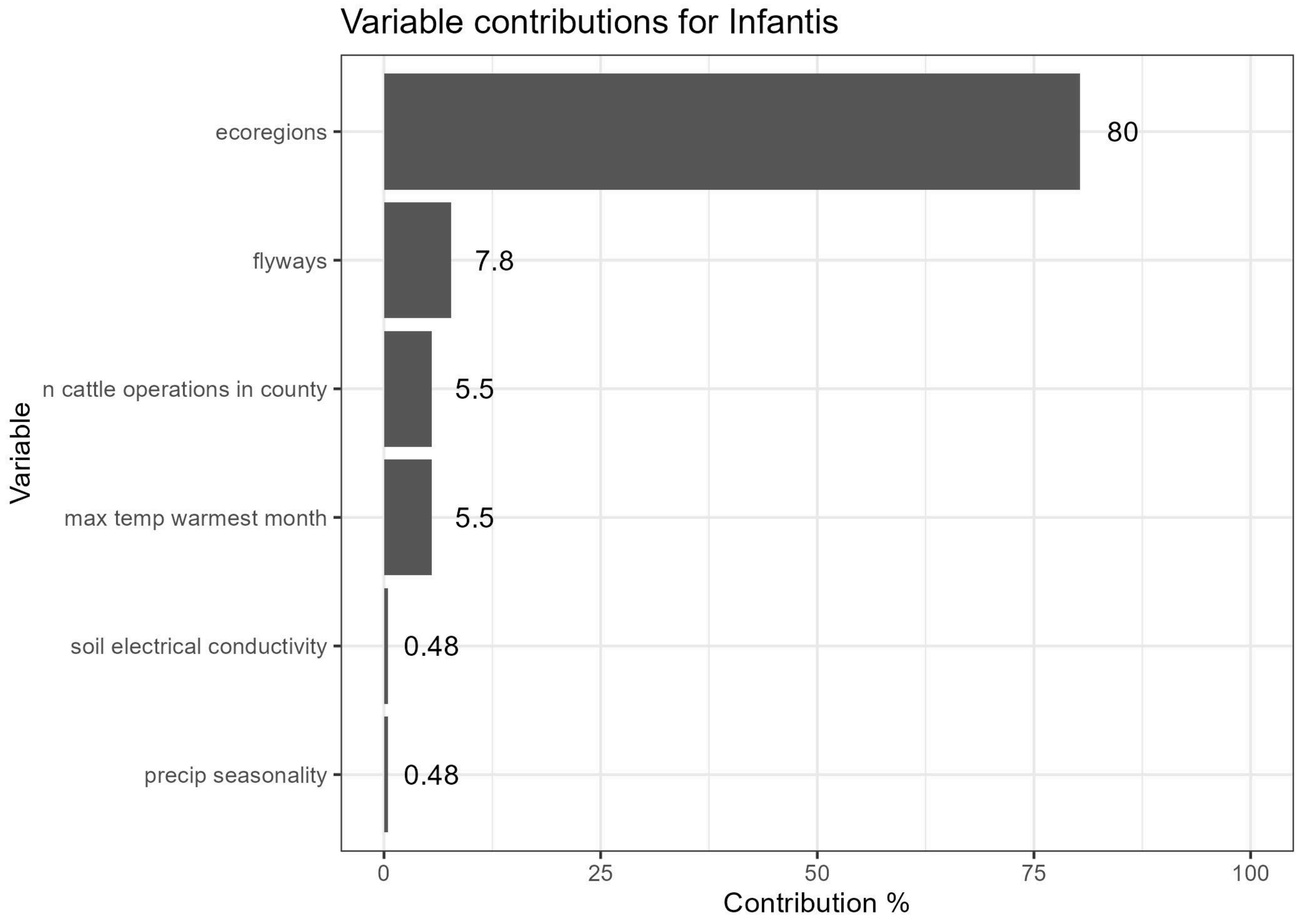


Lambdas for Infantis. Flyways are indicated as follows: (1) Atlantic, (2) Central, (3) Mississippi, (4) Pacific. See Supplementary File 1 for Ecoregion values.

| Feature | Variable | Type | Lambda | Minimum Encountered Value | Maximum Encountered Value |
|---|---|---|---|---|---|
| ecoregions==6.0 | ecoregions | categorical | 0.0000000 | 0.0000000 | 1.000000e+00 |
| ecoregions==10.0 | ecoregions | categorical | 0.7606584 | 0.0000000 | 1.000000e+00 |
| ecoregions==22.0 | ecoregions | categorical | 1.0555865 | 0.0000000 | 1.000000e+00 |
| flyways==1.0 | flyways | categorical | 0.0000000 | 0.0000000 | 1.000000e+00 |
| flyways==4.0 | flyways | categorical | -0.3003124 | 0.0000000 | 1.000000e+00 |
| Top5_Ca | soil calcium raw | linear | 0.0000000 | 0.0000000 | 2.550000e+02 |
| Top5_Ca_percWt | soil calcium as % | linear | 0.0000000 | 0.0695250 | 1.960904e+01 |
| Top5_Cu | soil copper raw | linear | 0.0000000 | 0.0000000 | 2.550000e+02 |
| Top5_Cu_mgPerKg | soil copper mg/kg | linear | 0.0000000 | 2.7603278 | 8.640745e+01 |
| Top5_Fe | soil iron raw | linear | 0.0000000 | 0.0000000 | 2.550000e+02 |
| Top5_Fe_percWt | soil iron % | linear | 0.0000000 | 0.1342737 | 6.557908e+00 |
| Top5_K | soil potassium raw | linear | 0.0000000 | 0.0000000 | 2.550000e+02 |
| Top5_K_percWt | soil potassium as % | linear | 0.0000000 | 0.0550254 | 3.489706e+00 |
| Top5_Mn | soil manganese raw | linear | 0.0000000 | 0.0000000 | 2.550000e+02 |
| Top5_Na | soil sodium raw | linear | 0.0000000 | 0.0000000 | 2.550000e+02 |
| Top5_Na_percWt | soil sodium % | linear | 0.0000000 | 0.0228538 | 2.729023e+00 |
| Top5_P | soil phosphorus raw | linear | 0.0000000 | 0.0000000 | 2.550000e+02 |

| Feature | Variable | Type | Lambda | Minimum Encountered Value | Maximum Encountered Value |
|---|---|---|---|---|---|
| Top5_P_mgPerKg | soil phosphorus mg/kg | linear | 0.0000000 | 96.5034027 | 2.682370e+03 |
| Top5_S | soil sulfur raw | linear | 0.0000000 | 0.0000000 | 2.550000e+02 |
| Top5_S_percWt | soil sulfur as % | linear | 0.0000000 | 0.0072269 | 3.062867e-01 |
| Top5_Zn | soil zinc raw | linear | 0.0000000 | 0.0000000 | 2.550000e+02 |
| Top5_Zn_mgPerKg | soil zinc mg/kg | linear | 0.0000000 | 6.1639123 | 4.239237e+02 |
| caco3_kg_sq_m | soil calcium carbonate | linear | 0.0000000 | 0.0000000 | 1.102458e+03 |
| cec_05 | soil cation exchange capacity | linear | 0.0000000 | 0.5051942 | 1.550606e+02 |
| drainage_class_int | soil drainage class | linear | 0.0000000 | 1.0000000 | 8.000000e+00 |
| ec_05 | soil electrical conductivity | linear | 0.0000000 | 0.0000000 | 1.400000e+02 |
| headCattle_nOperations | n cattle operations in county | linear | 0.0000000 | 4.0000000 | 4.000000e+01 |
| headCattle_totalValue | total value of cattle in county | linear | 0.0000000 | 4.0000000 | 4.838209e+06 |
| headOtherLivestock_nOperations | n other livestock operations in county | linear | 0.0000000 | 2.0000000 | 1.900000e+01 |
| headOtherLivestock_totalValue | total value of other livestock in county | linear | 0.0000000 | 2.0000000 | 3.769605e+06 |
| headPoultry_nOperations | n poultry operations in county | linear | 0.0000000 | 1.0000000 | 1.700000e+01 |
| headPoultry_totalValue | total value of poultry in county | linear | 0.0000000 | 1.0000000 | 3.690649e+07 |
| om_kg_sq_m | soil organic matter | linear | 0.0000000 | 0.0069909 | 3.200079e+02 |
| ph_05 | soil pH | linear | 0.0000000 | 3.2129552 | 9.500000e+00 |
| produce_nOperations | n produce operations in county | linear | 0.0000000 | 0.0000000 | 4.100000e+01 |
| texture_05 | soil texture | linear | 0.0000000 | 1.0000000 | 1.200000e+01 |
| wc2.1_2.5m_bio_10 | mean temp warmest quarter | linear | 0.0000000 | 6.8513331 | 3.623200e+01 |
| wc2.1_2.5m_bio_11 | mean temp coldest quarter | linear | 0.0000000 | -14.7773333 | 2.032067e+01 |
| wc2.1_2.5m_bio_12 | annual precipitation | linear | 0.0000000 | 56.0000000 | 3.343000e+03 |
| wc2.1_2.5m_bio_13 | precip wettest month | linear | 0.0000000 | 11.0000000 | 5.410000e+02 |
| wc2.1_2.5m_bio_14 | precip driest month | linear | 0.0000000 | 0.0000000 | 1.480000e+02 |
| wc2.1_2.5m_bio_15 | precip seasonality | linear | 0.0000000 | 5.5102468 | 9.287923e+01 |
| wc2.1_2.5m_bio_16 | precip wettest quarter | linear | 0.0000000 | 27.0000000 | 1.479000e+03 |
| wc2.1_2.5m_bio_17 | precip driest quarter | linear | 0.0000000 | 2.0000000 | 4.670000e+02 |
| wc2.1_2.5m_bio_18 | precip warmest quarter | linear | 0.0000000 | 2.0000000 | 6.390000e+02 |

| Feature | Variable | Type | Lambda | Minimum Encountered Value | Maximum Encountered Value |
|---|---|---|---|---|---|
| wc2.1_2.5m_bio_19 | precip coldest quarter | linear | 0.0000000 | 15.0000000 | 1.411000e+03 |
| wc2.1_2.5m_bio_1 | annual mean temp | linear | 0.0000000 | -2.3499999 | 2.450133e+01 |
| wc2.1_2.5m_bio_2 | mean diurnal range | linear | 0.0000000 | 6.0111108 | 2.171233e+01 |
| wc2.1_2.5m_bio_3 | isothermality | linear | 0.0000000 | 23.8849716 | 6.461557e+01 |
| wc2.1_2.5m_bio_4 | temperature seasonality | linear | 0.0000000 | 260.6383972 | 1.354152e+03 |
| wc2.1_2.5m_bio_5 | max temp warmest month | linear | 0.0000000 | 14.4480000 | 4.595600e+01 |
| wc2.1_2.5m_bio_6 | min temp coldest month | linear | 0.0000000 | -23.3759995 | 1.509200e+01 |
| wc2.1_2.5m_bio_7 | annual temp range | linear | 0.0000000 | 16.5928574 | 5.016400e+01 |
| wc2.1_2.5m_bio_8 | mean temp wettest quarter | linear | 0.0000000 | -8.7173328 | 3.331267e+01 |
| wc2.1_2.5m_bio_9 | mean temp driest quarter | linear | 0.0000000 | -14.7773333 | 2.880467e+01 |
| ec_05*wc2.1_2.5m_bio_15 | soil electrical conductivity,precip seasonality | product | -3.8960683 | 0.0000000 | 3.331397e+03 |
| headCattle_nOperations*wc2.1_2.5m_bio_5 | n cattle operations in county,max temp warmest month | product | 1.3322209 | 91.2959976 | 1.445400e+03 |

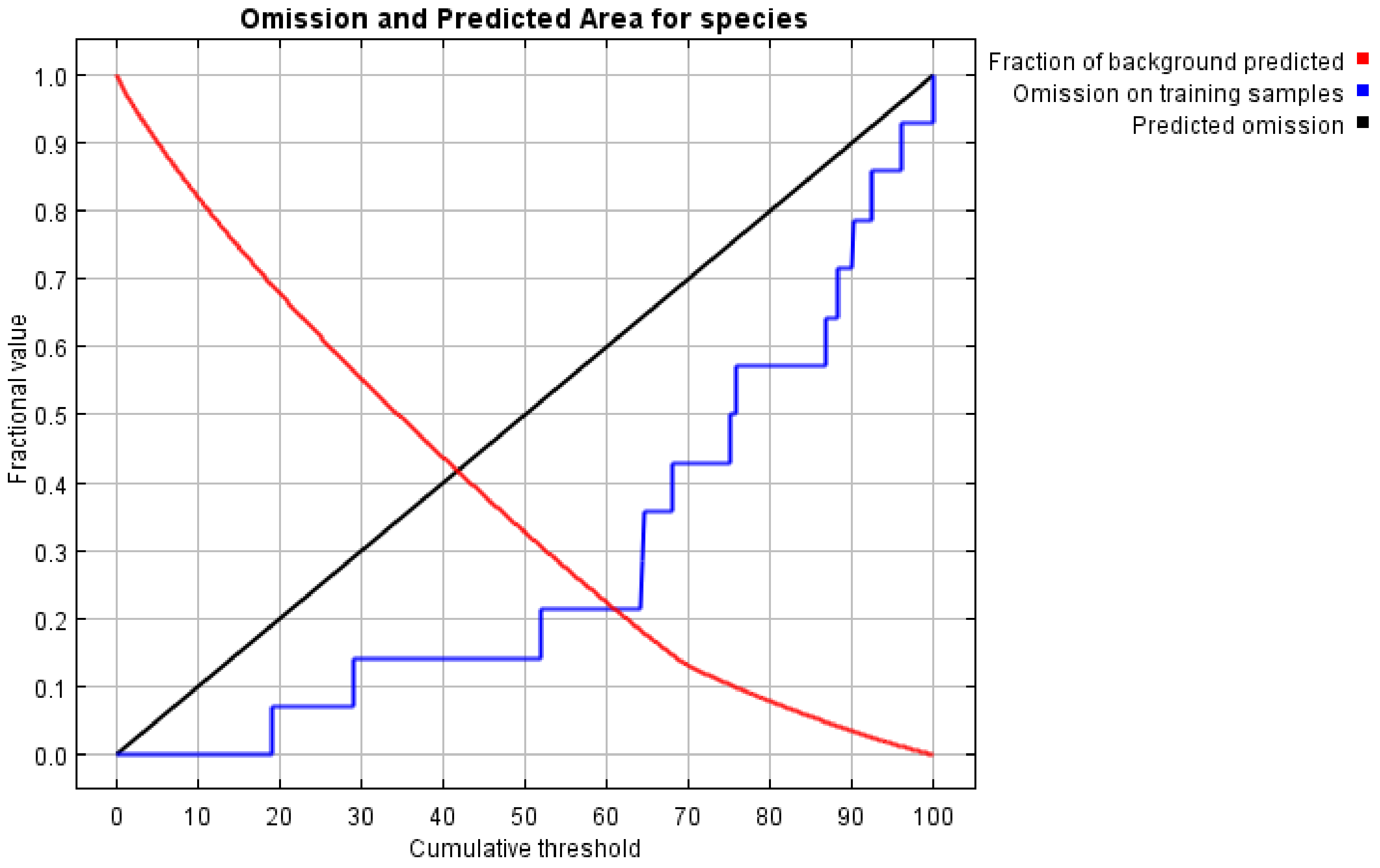

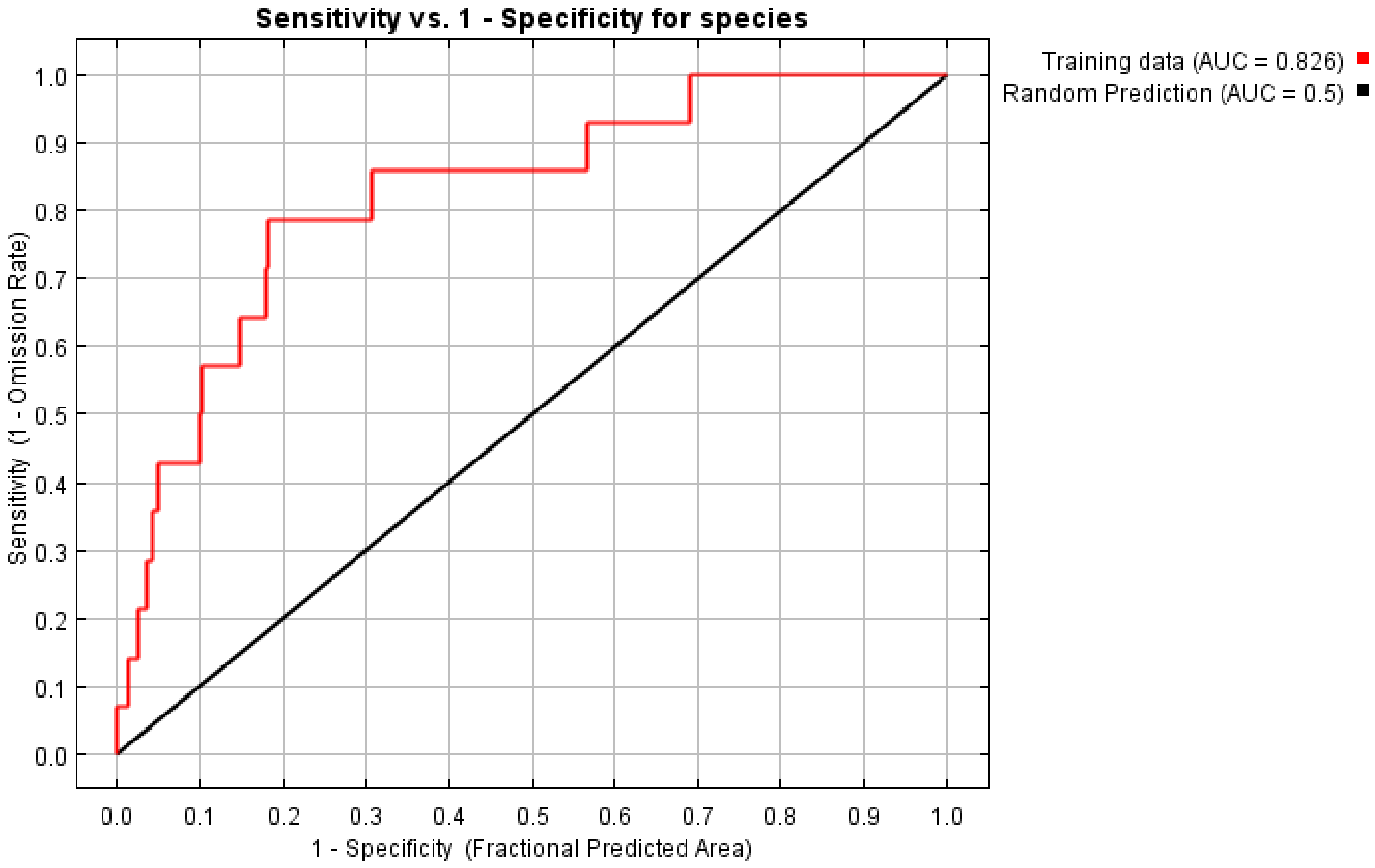

Sensitivity vs. 1 - Specificity for species
Training data (AUC = 0.826)
Random Prediction (AUC = 0.5)
Sensitivity (1 - Omission Rate)
1 - Specificity (Fractional Predicted Area)
0.0
0.1
0.2
0.3
0.4
0.5
0.6
0.7
0.8
0.9
1.0
