## Supplementary File 11 for "Geographic Distributions of Top Beef Salmonella Serovars in the U.S."

### Species Distribution Model Results for Serovar Meleagridis

Model tuning results for Meleagridis.

| N Points | N Training Points | N Background Points | Regularization Multiplier | Features | Training AUC |
|---|---|---|---|---|---|
| 34 | 16 | 10016 | 4 | LQPT | 0.69 |

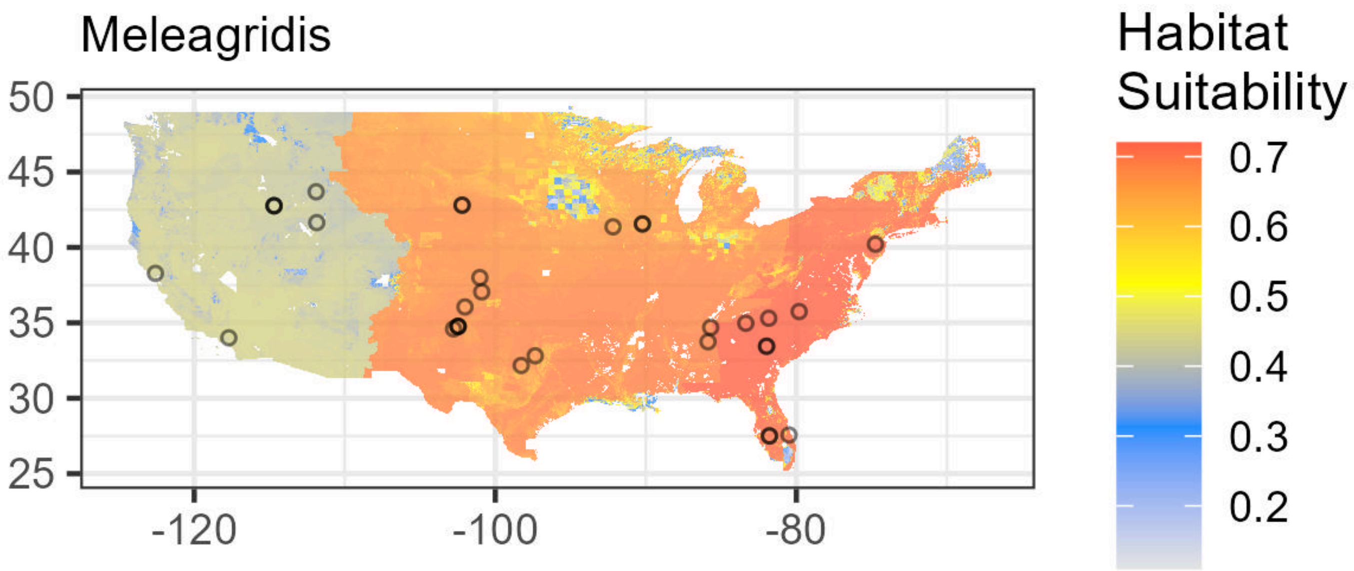

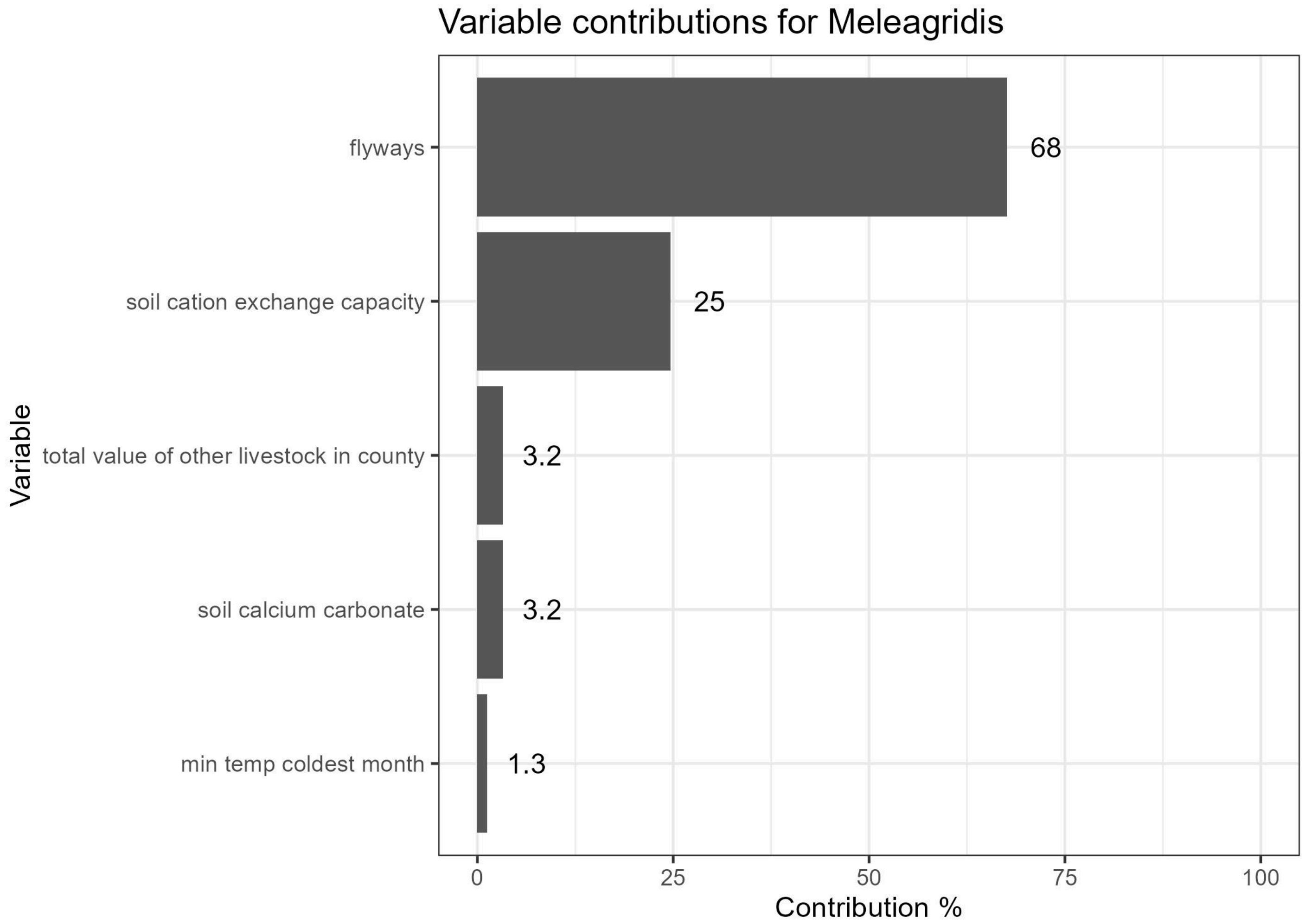


Lambdas for Meleagridis. Flyways are indicated as follows: (1) Atlantic, (2) Central, (3) Mississippi, (4) Pacific. See Supplementary File 1 for Ecoregion values.

| Feature | Variable | Type | Lambda | Minimum Encountered Value | Maximum Encountered Value |
|---|---|---|---|---|---|
| ecoregions==6.0 | ecoregions | categorical | 0.0000000 | 0.0000000 | 1.000000e+00 |
| flyways==1.0 | flyways | categorical | 0.0873649 | 0.0000000 | 1.000000e+00 |
| flyways==4.0 | flyways | categorical | -0.6862936 | 0.0000000 | 1.000000e+00 |
| Top5_Ca | soil calcium raw | linear | 0.0000000 | 0.0000000 | 2.550000e+02 |
| Top5_Ca_percWt | soil calcium as % | linear | 0.0000000 | 0.0600963 | 1.879637e+01 |
| Top5_Cu | soil copper raw | linear | 0.0000000 | 0.0000000 | 2.550000e+02 |
| Top5_Cu_mgPerKg | soil copper mg/kg | linear | 0.0000000 | 3.2157192 | 9.937318e+01 |
| Top5_Fe | soil iron raw | linear | 0.0000000 | 0.0000000 | 2.550000e+02 |
| Top5_Fe_percWt | soil iron % | linear | 0.0000000 | 0.1198162 | 6.861789e+00 |
| Top5_K | soil potassium raw | linear | 0.0000000 | 0.0000000 | 2.550000e+02 |
| Top5_K_percWt | soil potassium as % | linear | 0.0000000 | 0.0429253 | 3.389892e+00 |

| Feature | Variable | Type | Lambda | Minimum Encountered Value | Maximum Encountered Value |
|---|---|---|---|---|---|
| Top5_Mn | soil manganese raw | linear | 0.0000000 | 0.0000000 | 2.550000e+02 |
| Top5_Na | soil sodium raw | linear | 0.0000000 | 0.0000000 | 2.550000e+02 |
| Top5_Na_percWt | soil sodium % | linear | 0.0000000 | 0.0186184 | 2.556385e+00 |
| Top5_P | soil phosphorus raw | linear | 0.0000000 | 0.0000000 | 2.550000e+02 |
| Top5_P_mgPerKg | soil phosphorus mg/kg | linear | 0.0000000 | 104.0249023 | 2.351274e+03 |
| Top5_S | soil sulfur raw | linear | 0.0000000 | 0.0000000 | 2.550000e+02 |
| Top5_S_percWt | soil sulfur as % | linear | 0.0000000 | 0.0065469 | 3.401941e-01 |
| Top5_Zn | soil zinc raw | linear | 0.0000000 | 0.0000000 | 2.550000e+02 |
| Top5_Zn_mgPerKg | soil zinc mg/kg | linear | 0.0000000 | 7.4528823 | 4.241208e+02 |
| caco3_kg_sq_m | soil calcium carbonate | linear | 0.0000000 | 0.0000000 | 1.102458e+03 |
| cec_05 | soil cation exchange capacity | linear | 0.0000000 | 0.9165860 | 1.647350e+02 |
| drainage_class_int | soil drainage class | linear | 0.0000000 | 1.0000000 | 8.000000e+00 |
| ec_05 | soil electrical conductivity | linear | 0.0000000 | 0.0000000 | 1.400000e+02 |
| headCattle_nOperations | n cattle operations in county | linear | 0.0000000 | 4.0000000 | 4.000000e+01 |
| headCattle_totalValue | total value of cattle in county | linear | 0.0000000 | 4.0000000 | 4.838209e+06 |
| headOtherLivestock_nOperations | n other livestock operations in county | linear | 0.0000000 | 2.0000000 | 1.900000e+01 |
| headOtherLivestock_totalValue | total value of other livestock in county | linear | 0.0000000 | 2.0000000 | 3.769605e+06 |
| headPoultry_nOperations | n poultry operations in county | linear | 0.0000000 | 1.0000000 | 1.700000e+01 |
| headPoultry_totalValue | total value of poultry in county | linear | 0.0000000 | 1.0000000 | 3.690649e+07 |
| om_kg_sq_m | soil organic matter | linear | 0.0000000 | 0.0000000 | 4.576203e+02 |
| ph_05 | soil pH | linear | 0.0000000 | 3.3669238 | 9.654279e+00 |
| produce_nOperations | n produce operations in county | linear | 0.0000000 | 0.0000000 | 4.000000e+01 |

| Feature | Variable | Type | Lambda | Minimum Encountered Value | Maximum Encountered Value |
|---|---|---|---|---|---|
| texture_05 | soil texture | linear | 0.0000000 | 1.0000000 | 1.200000e+01 |
| wc2.1_2.5m_bio_10 | mean temp warmest quarter | linear | 0.0000000 | 6.4173331 | 3.598000e+01 |
| wc2.1_2.5m_bio_11 | mean temp coldest quarter | linear | 0.0000000 | -15.1380005 | 2.045267e+01 |
| wc2.1_2.5m_bio_12 | annual precipitation | linear | 0.0000000 | 57.0000000 | 3.189000e+03 |
| wc2.1_2.5m_bio_13 | precip wettest month | linear | 0.0000000 | 10.0000000 | 5.160000e+02 |
| wc2.1_2.5m_bio_14 | precip driest month | linear | 0.0000000 | 0.0000000 | 1.480000e+02 |
| wc2.1_2.5m_bio_15 | precip seasonality | linear | 0.0000000 | 5.5347414 | 9.477441e+01 |
| wc2.1_2.5m_bio_16 | precip wettest quarter | linear | 0.0000000 | 28.0000000 | 1.422000e+03 |
| wc2.1_2.5m_bio_17 | precip driest quarter | linear | 0.0000000 | 2.0000000 | 4.620000e+02 |
| wc2.1_2.5m_bio_18 | precip warmest quarter | linear | 0.0000000 | 2.0000000 | 6.380000e+02 |
| wc2.1_2.5m_bio_19 | precip coldest quarter | linear | 0.0000000 | 17.0000000 | 1.357000e+03 |
| wc2.1_2.5m_bio_1 | annual mean temp | linear | 0.0000000 | -2.7888334 | 2.458200e+01 |
| wc2.1_2.5m_bio_2 | mean diurnal range | linear | 0.0000000 | 6.4159093 | 2.107167e+01 |
| wc2.1_2.5m_bio_3 | isothermality | linear | 0.0000000 | 22.9389553 | 6.441906e+01 |
| wc2.1_2.5m_bio_4 | temperature seasonality | linear | 0.0000000 | 201.2461853 | 1.360078e+03 |
| wc2.1_2.5m_bio_5 | max temp warmest month | linear | 0.0000000 | 14.8520002 | 4.531200e+01 |
| wc2.1_2.5m_bio_6 | min temp coldest month | linear | 0.0000000 | -23.6440010 | 1.627273e+01 |
| wc2.1_2.5m_bio_7 | annual temp range | linear | 0.0000000 | 15.0428581 | 4.997600e+01 |
| wc2.1_2.5m_bio_8 | mean temp wettest quarter | linear | 0.0000000 | -10.9726667 | 3.358866e+01 |
| wc2.1_2.5m_bio_9 | mean temp driest quarter | linear | 0.0000000 | -14.8206673 | 2.855867e+01 |
| cec_05^2 | soil cation exchange capacity | quadratic | -2.1294181 | 0.8401299 | 2.713762e+04 |
| wc2.1_2.5m_bio_6^2 | min temp coldest month | quadratic | -0.1471728 | 0.0000000 | 5.590388e+02 |

| Feature | Variable | Type | Lambda | Minimum Encountered Value | Maximum Encountered Value |
|---|---|---|---|---|---|
| caco3_kg_sq_m*headOtherLivestock_totalValue | soil calcium carbonate,total value of other livestock in county | product | -2.0798823 | 0.0000000 | 6.508517e+08 |

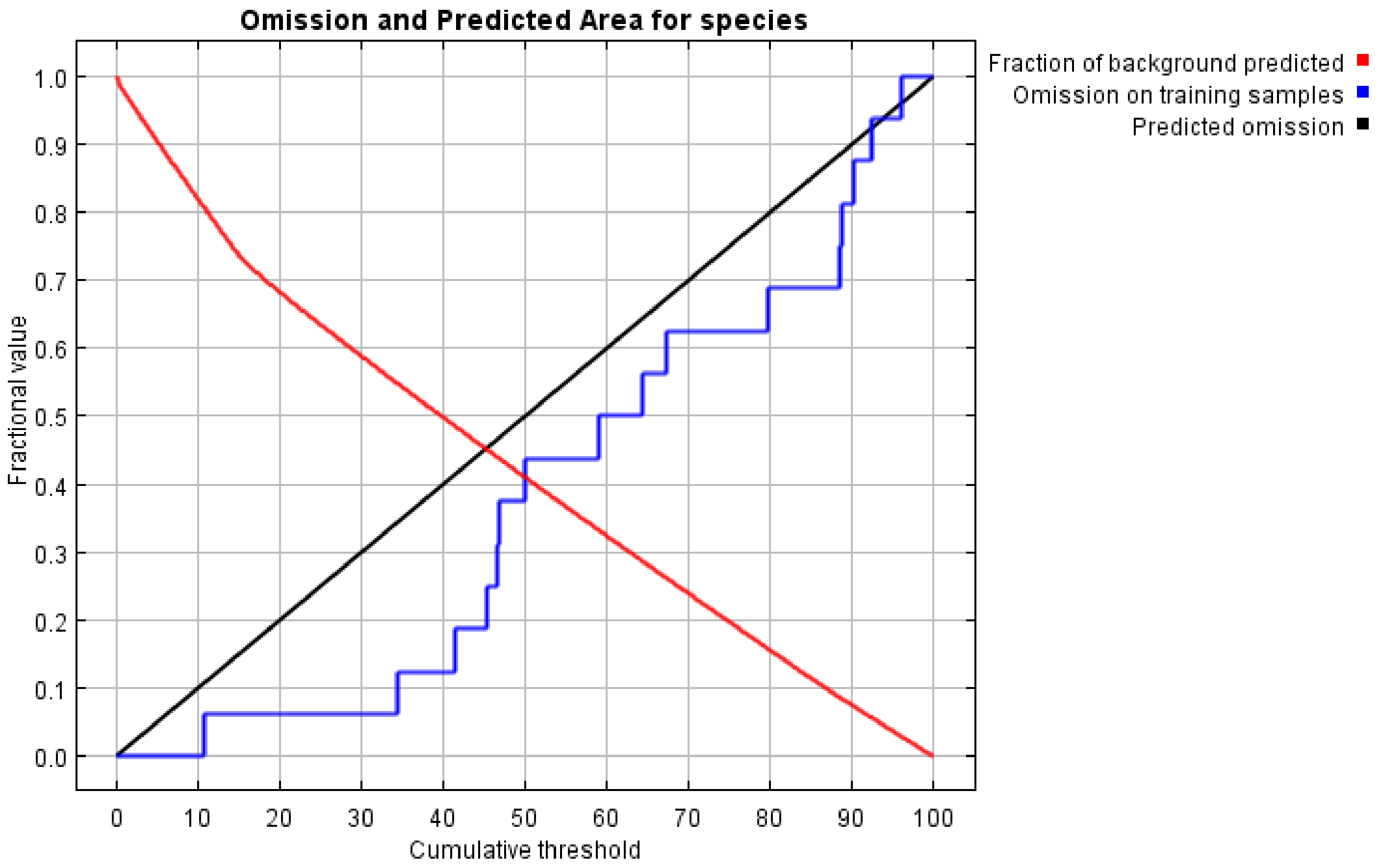

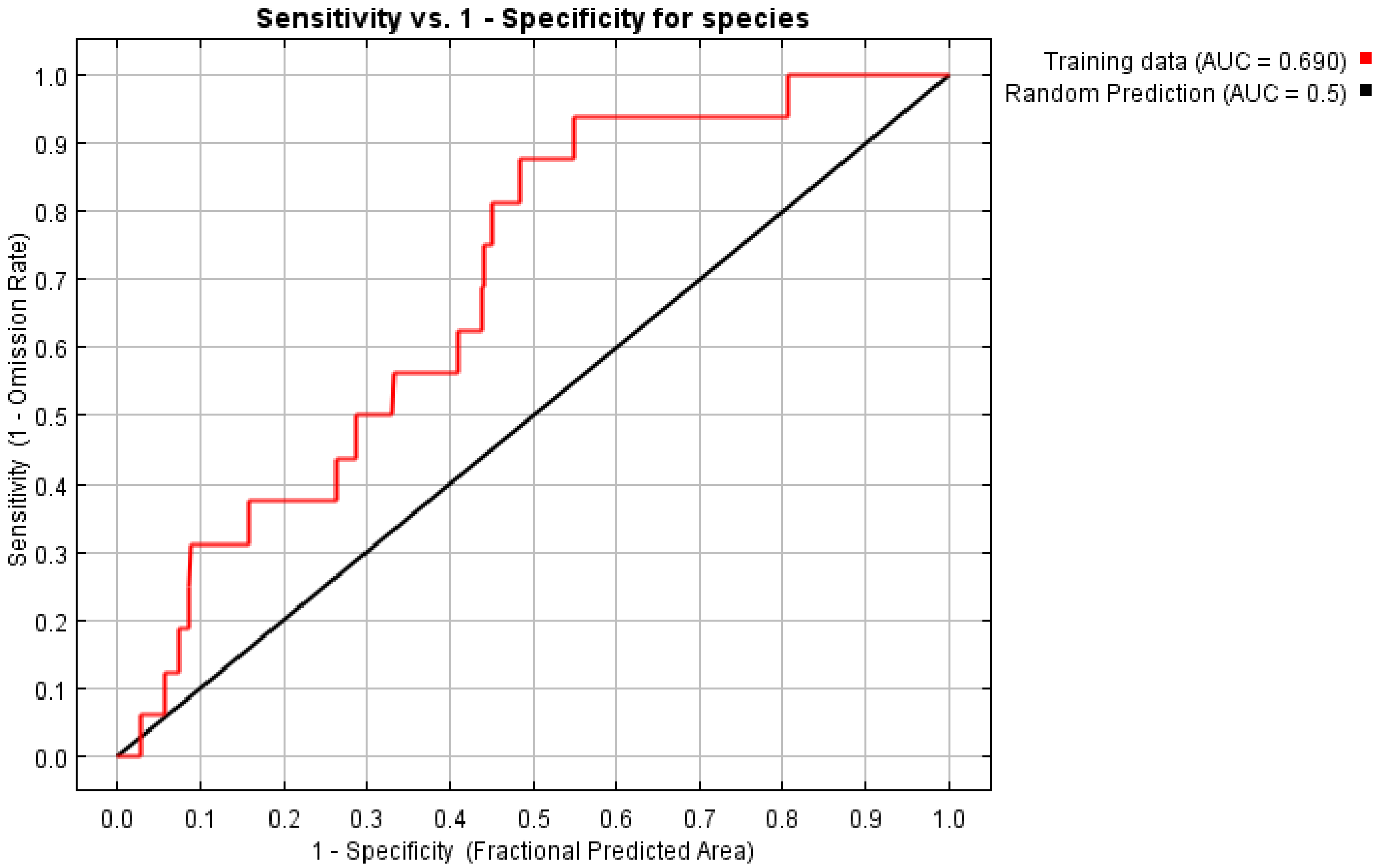

Sensitivity vs. 1 - Specificity for species
Training data (AUC = 0.690)
Random Prediction (AUC = 0.5)
Sensitivity (1 - Omission Rate)
1 - Specificity (Fractional Predicted Area)
0.0
0.1
0.2
0.3
0.4
0.5
0.6
0.7
0.8
0.9
1.0
