## Supplementary File 12 for "Geographic Distributions of Top Beef Salmonella Serovars in the U.S."

### Species Distribution Model Results for Serovar Montevideo

Model tuning results for Montevideo.

| N Points | N Training Points | N Background Points | Regularization Multiplier | Features | Training AUC |
|---|---|---|---|---|---|
| 228 | 44 | 10040 | 4 | LQPT | 0.87 |

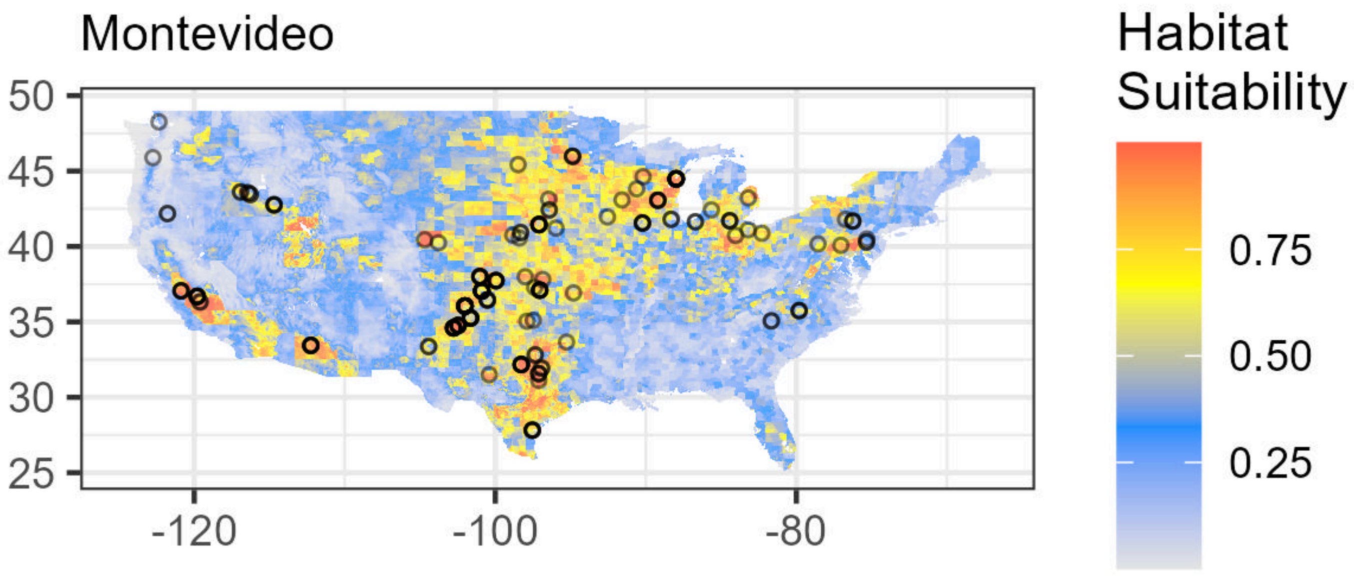

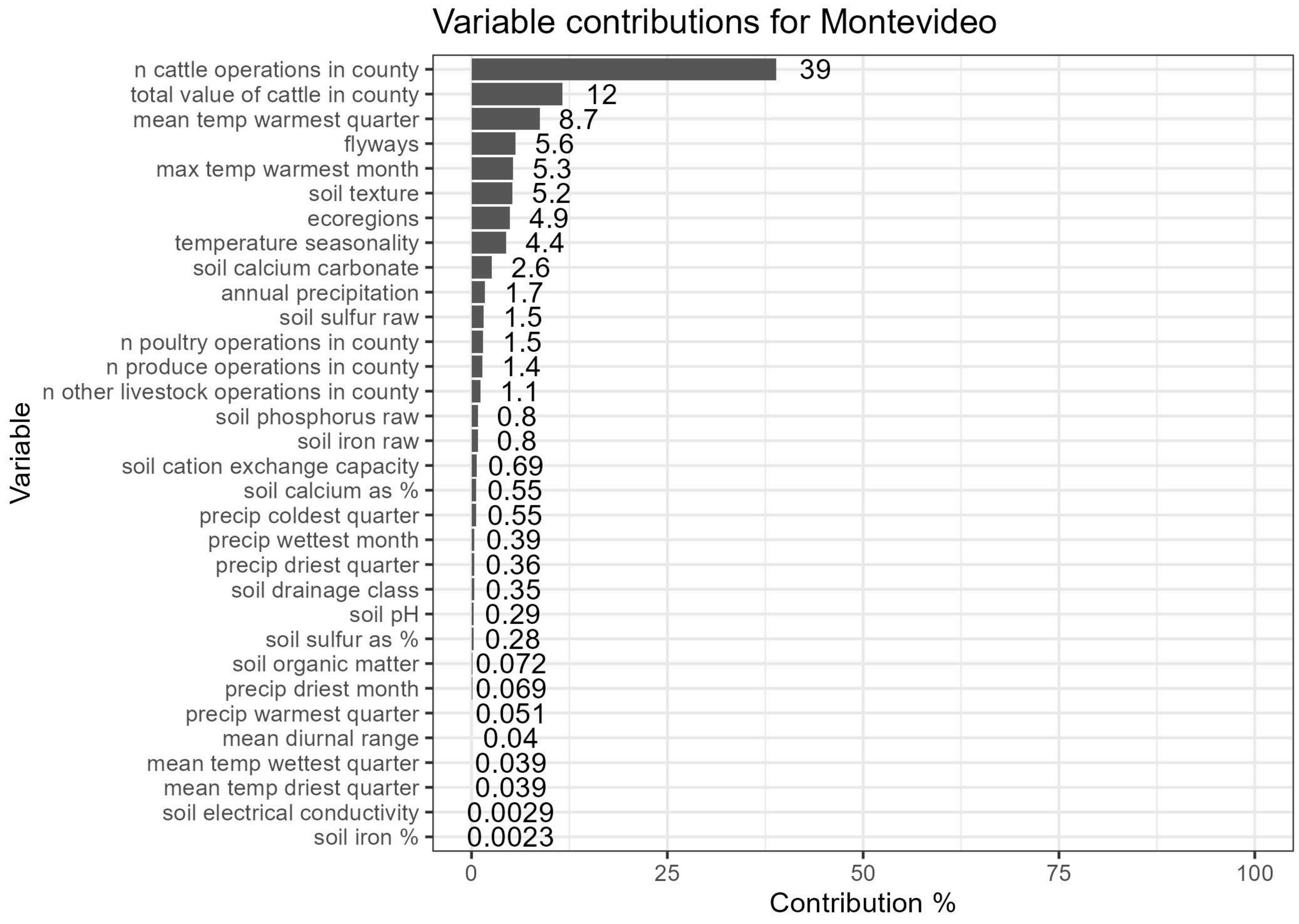


Lambdas for Montevideo. Flyways are indicated as follows: (1) Atlantic, (2) Central, (3) Mississippi, (4) Pacific. See Supplementary File 1 for Ecoregion values.

| Feature | Variable | Type | Lambda | Minimum Encountered Value | Maximum Encountered Value |
|---|---|---|---|---|---|
| ecoregions==6.0 | ecoregions | categorical | 0.0000000 | 0.0000000 | 1.000000e+00 |
| ecoregions==23.0 | ecoregions | categorical | 0.6293740 | 0.0000000 | 1.000000e+00 |
| ecoregions==40.0 | ecoregions | categorical | -0.0209730 | 0.0000000 | 1.000000e+00 |
| flyways==1.0 | flyways | categorical | 0.0000000 | 0.0000000 | 1.000000e+00 |
| flyways==2.0 | flyways | categorical | 0.0690669 | 0.0000000 | 1.000000e+00 |
| Top5_Ca | soil calcium raw | linear | 0.0000000 | 0.0000000 | 2.550000e+02 |
| Top5_Ca_percWt | soil calcium as % | linear | 0.0000000 | 0.0656684 | 2.028087e+01 |
| Top5_Cu | soil copper raw | linear | 0.0000000 | 0.0000000 | 2.550000e+02 |
| Top5_Cu_mgPerKg | soil copper mg/kg | linear | 0.0000000 | 2.4137752 | 8.277317e+01 |
| Top5_Fe | soil iron raw | linear | 0.0000000 | 0.0000000 | 2.550000e+02 |
| Top5_Fe_percWt | soil iron % | linear | 0.0000000 | 0.1342737 | 7.543058e+00 |
| Top5_K | soil potassium raw | linear | 0.0000000 | 0.0000000 | 2.550000e+02 |
| Top5_K_percWt | soil potassium as % | linear | 0.0000000 | 0.0550254 | 3.180072e+00 |
| Top5_Mn | soil manganese raw | linear | 0.0000000 | 0.0000000 | 2.550000e+02 |
| Top5_Na | soil sodium raw | linear | 0.0000000 | 0.0000000 | 2.550000e+02 |

| Feature | Variable | Type | Lambda | Minimum Encountered Value | Maximum Encountered Value |
|---|---|---|---|---|---|
| Top5_Na_percWt | soil sodium % | linear | 0.0000000 | 0.0207029 | 2.577029e+00 |
| Top5_P | soil phosphorus raw | linear | 0.0000000 | 0.0000000 | 2.550000e+02 |
| Top5_P_mgPerKg | soil phosphorus mg/kg | linear | 0.0000000 | 96.5034027 | 2.500958e+03 |
| Top5_S | soil sulfur raw | linear | 0.0000000 | 0.0000000 | 2.550000e+02 |
| Top5_S_percWt | soil sulfur as % | linear | 0.0000000 | 0.0066458 | 2.969956e-01 |
| Top5_Zn | soil zinc raw | linear | 0.0000000 | 0.0000000 | 2.550000e+02 |
| Top5_Zn_mgPerKg | soil zinc mg/kg | linear | 0.0000000 | 5.7095504 | 3.521571e+02 |
| caco3_kg_sq_m | soil calcium carbonate | linear | 0.0000000 | 0.0000000 | 1.053772e+03 |
| cec_05 | soil cation exchange capacity | linear | 0.0000000 | 0.0683986 | 1.950097e+02 |
| drainage_class_int | soil drainage class | linear | 0.0000000 | 1.0000000 | 8.000000e+00 |
| ec_05 | soil electrical conductivity | linear | 0.0000000 | 0.0000000 | 1.400000e+02 |
| headCattle_nOperations | n cattle operations in county | linear | 0.0000000 | 4.0000000 | 4.000000e+01 |
| headCattle_totalValue | total value of cattle in county | linear | 0.0000000 | 4.0000000 | 4.838209e+06 |
| headOtherLivestock_nOperations | n other livestock operations in county | linear | 0.0000000 | 2.0000000 | 1.900000e+01 |
| headOtherLivestock_totalValue | total value of other livestock in county | linear | 0.0000000 | 2.0000000 | 3.769605e+06 |
| headPoultry_nOperations | n poultry operations in county | linear | 0.0000000 | 1.0000000 | 1.700000e+01 |
| headPoultry_totalValue | total value of poultry in county | linear | 0.0000000 | 1.0000000 | 3.690649e+07 |
| om_kg_sq_m | soil organic matter | linear | 0.0000000 | 0.1234893 | 5.875868e+02 |
| ph_05 | soil pH | linear | 0.0000000 | 3.2119956 | 1.004157e+01 |
| produce_nOperations | n produce operations in county | linear | 0.0000000 | 0.0000000 | 4.000000e+01 |
| texture_05 | soil texture | linear | 0.0000000 | 1.0000000 | 1.200000e+01 |
| wc2.1_2.5m_bio_10 | mean temp warmest quarter | linear | 0.0000000 | 2.8539999 | 3.610333e+01 |
| wc2.1_2.5m_bio_11 | mean temp coldest quarter | linear | 0.0000000 | -14.7726669 | 2.048333e+01 |
| wc2.1_2.5m_bio_12 | annual precipitation | linear | 0.0000000 | 57.0000000 | 3.242000e+03 |
| wc2.1_2.5m_bio_13 | precip wettest month | linear | 0.0000000 | 9.0000000 | 5.310000e+02 |
| wc2.1_2.5m_bio_14 | precip driest month | linear | 0.0000000 | 0.0000000 | 1.420000e+02 |

| Feature | Variable | Type | Lambda | Minimum Encountered Value | Maximum Encountered Value |
|---|---|---|---|---|---|
| wc2.1_2.5m_bio_15 | precip seasonality | linear | 0.0000000 | 5.4699855 | 9.451818e+01 |
| wc2.1_2.5m_bio_16 | precip wettest quarter | linear | 0.0000000 | 26.0000000 | 1.457000e+03 |
| wc2.1_2.5m_bio_17 | precip driest quarter | linear | 0.0000000 | 2.0000000 | 4.450000e+02 |
| wc2.1_2.5m_bio_18 | precip warmest quarter | linear | 0.0000000 | 2.0000000 | 6.430000e+02 |
| wc2.1_2.5m_bio_19 | precip coldest quarter | linear | 0.0000000 | 17.0000000 | 1.379000e+03 |
| wc2.1_2.5m_bio_1 | annual mean temp | linear | 0.0000000 | -3.4956667 | 2.458783e+01 |
| wc2.1_2.5m_bio_2 | mean diurnal range | linear | 0.0000000 | 6.4159093 | 2.144800e+01 |
| wc2.1_2.5m_bio_3 | isothermality | linear | 0.0000000 | 23.9120445 | 5.967122e+01 |
| wc2.1_2.5m_bio_4 | temperature seasonality | linear | 0.0000000 | 269.3755493 | 1.363594e+03 |
| wc2.1_2.5m_bio_5 | max temp warmest month | linear | 0.0000000 | 9.0719995 | 4.549600e+01 |
| wc2.1_2.5m_bio_6 | min temp coldest month | linear | 0.0000000 | -23.5520000 | 1.627273e+01 |
| wc2.1_2.5m_bio_7 | annual temp range | linear | 0.0000000 | 15.0636368 | 4.970400e+01 |
| wc2.1_2.5m_bio_8 | mean temp wettest quarter | linear | 0.0000000 | -9.0813332 | 3.327533e+01 |
| wc2.1_2.5m_bio_9 | mean temp driest quarter | linear | 0.0000000 | -14.7726669 | 2.868867e+01 |
| Top5_Ca_percWt*Top5_Fe | soil calcium as %,soil iron raw | product | -2.9275165 | 0.0000000 | 3.053153e+03 |
| Top5_Fe*wc2.1_2.5m_bio_19 | soil iron raw,precip coldest quarter | product | -0.1202093 | 0.0000000 | 3.516450e+05 |
| Top5_P*caco3_kg_sq_m | soil phosphorus raw,soil calcium carbonate | product | 1.0636915 | 0.0000000 | 2.563539e+05 |
| Top5_P*headCattle_totalValue | soil phosphorus raw,total value of cattle in county | product | 0.7899290 | 0.0000000 | 1.233743e+09 |
| Top5_S*headPoultry_nOperations | soil sulfur raw,n poultry operations in county | product | 1.0124145 | 0.0000000 | 4.335000e+03 |
| Top5_S*produce_nOperations | soil sulfur raw,n produce operations in county | product | 0.1009309 | 0.0000000 | 1.020000e+04 |
| Top5_S_percWt*wc2.1_2.5m_bio_2 | soil sulfur as %,mean diurnal range | product | -0.8847097 | 0.0877977 | 5.143865e+00 |
| caco3_kg_sq_m*wc2.1_2.5m_bio_12 | soil calcium carbonate,annual precipitation | product | 0.9222924 | 0.0000000 | 7.081453e+05 |

| Feature | Variable | Type | Lambda | Minimum Encountered Value | Maximum Encountered Value |
|---|---|---|---|---|---|
| headCattle_nOperations*produce_nOperations | n cattle operations in county,n produce operations in county | product | 0.7176172 | 0.0000000 | 1.400000e+03 |
| headCattle_nOperations*wc2.1_2.5m_bio_10 | n cattle operations in county,mean temp warmest quarter | product | 4.3215188 | 62.2880020 | 1.146156e+03 |
| headCattle_totalValue*wc2.1_2.5m_bio_12 | total value of cattle in county,annual precipitation | product | 2.2667639 | 2025.0000000 | 3.154512e+09 |
| headCattle_totalValue*wc2.1_2.5m_bio_17 | total value of cattle in county,precip driest quarter | product | 0.3617733 | 195.0000000 | 2.227546e+08 |
| produce_nOperations*texture_05 | n produce operations in county,soil texture | product | 0.3836541 | 0.0000000 | 3.172434e+02 |
| produce_nOperations*wc2.1_2.5m_bio_10 | n produce operations in county,mean temp warmest quarter | product | 0.1970861 | 0.0000000 | 9.835640e+02 |
| wc2.1_2.5m_bio_12*wc2.1_2.5m_bio_19 | annual precipitation,precip coldest quarter | product | -6.2659546 | 1311.0000000 | 4.470718e+06 |

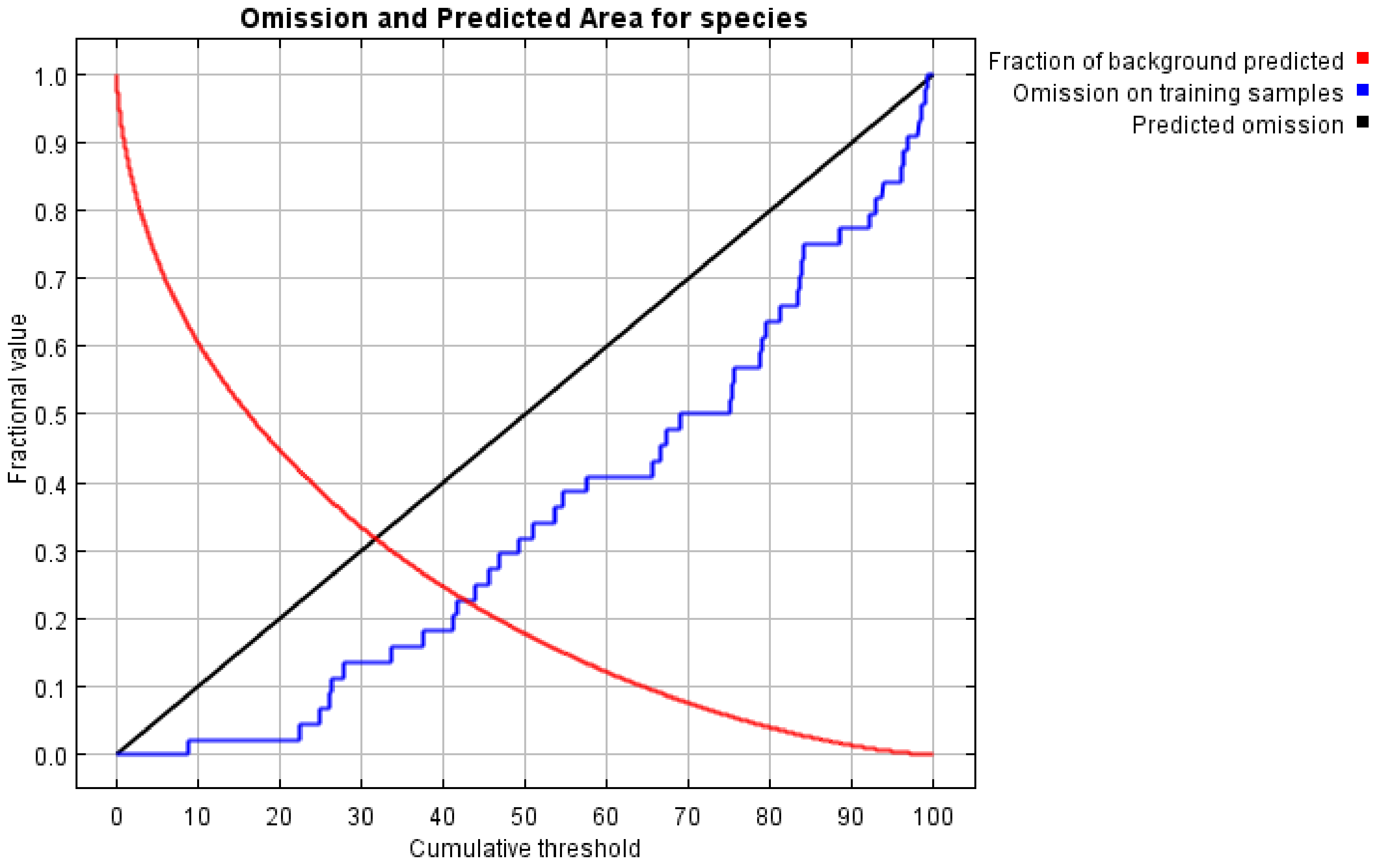
Omission and Predicted Area for species
Fraction of background predicted
Omission on training samples
Predicted omission
Fractional value
Cumulative threshold
0.0
0.1
0.2
0.3
0.4
0.5
0.6
0.7
0.8
0.9
1.0
0
10
20
30
40
50
60
70
80
90
100

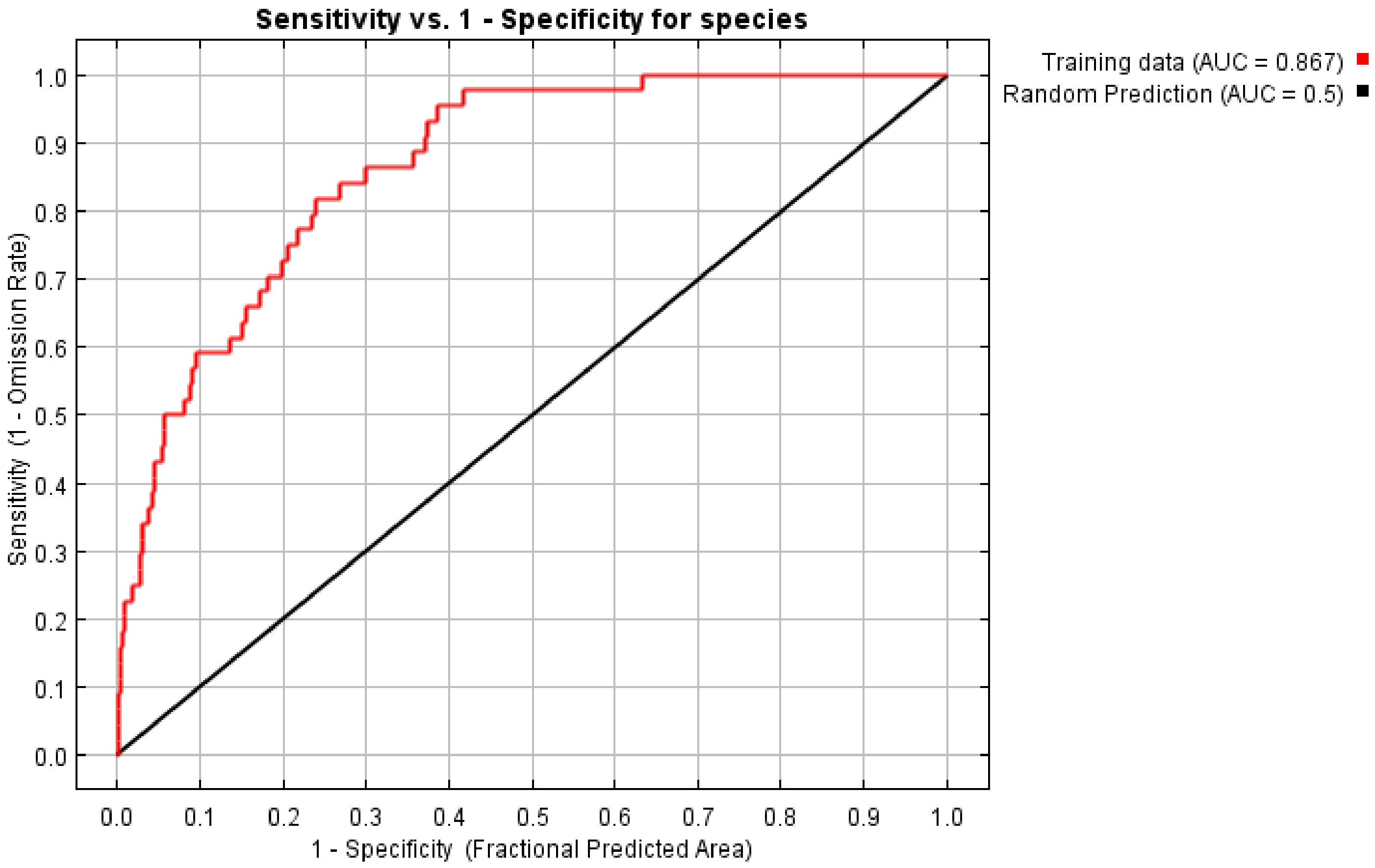
Sensitivity vs. 1 - Specificity for species
Training data (AUC = 0.867)
Random Prediction (AUC = 0.5)
Sensitivity (1 - Omission Rate)
1 - Specificity (Fractional Predicted Area)
0.0
0.1
0.2
0.3
0.4
0.5
0.6
0.7
0.8
0.9
1.0
