## Supplementary File 13 for "Geographic Distributions of Top Beef Salmonella Serovars in the U.S."

### Species Distribution Model Results for Serovar Muenchen

Model tuning results for Muenchen.

| N Points | N Training Points | N Background Points | Regularization Multiplier | Features | Training AUC |
|---|---|---|---|---|---|
| 83 | 26 | 10021 | 4 | LQPT | 0.85 |

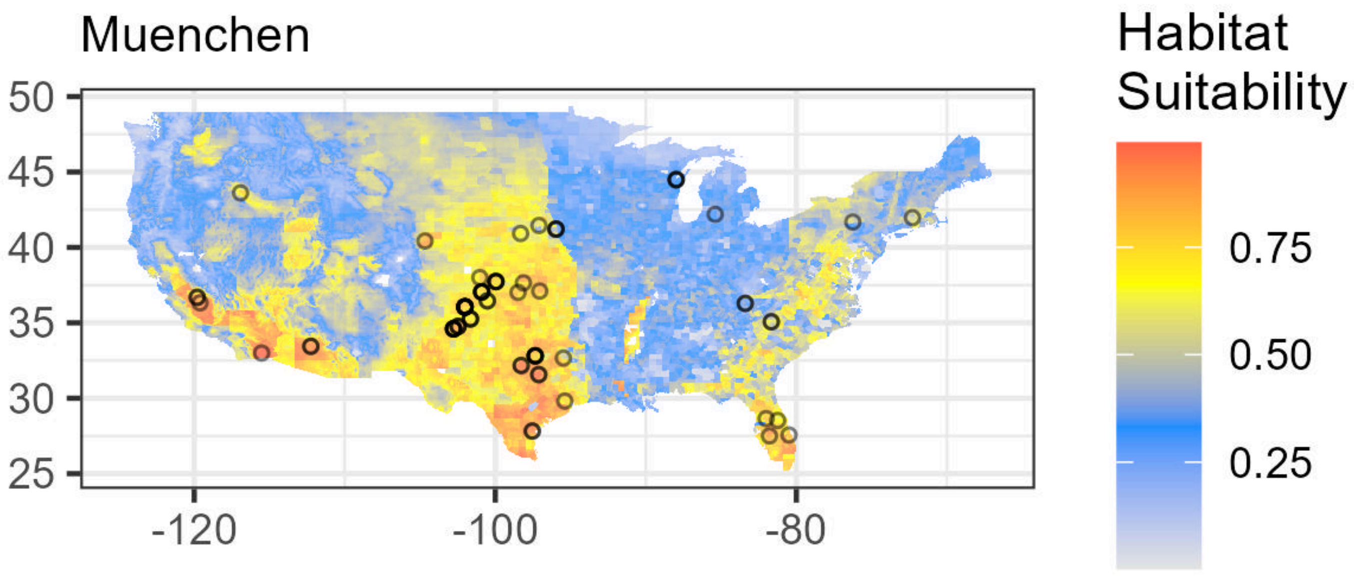

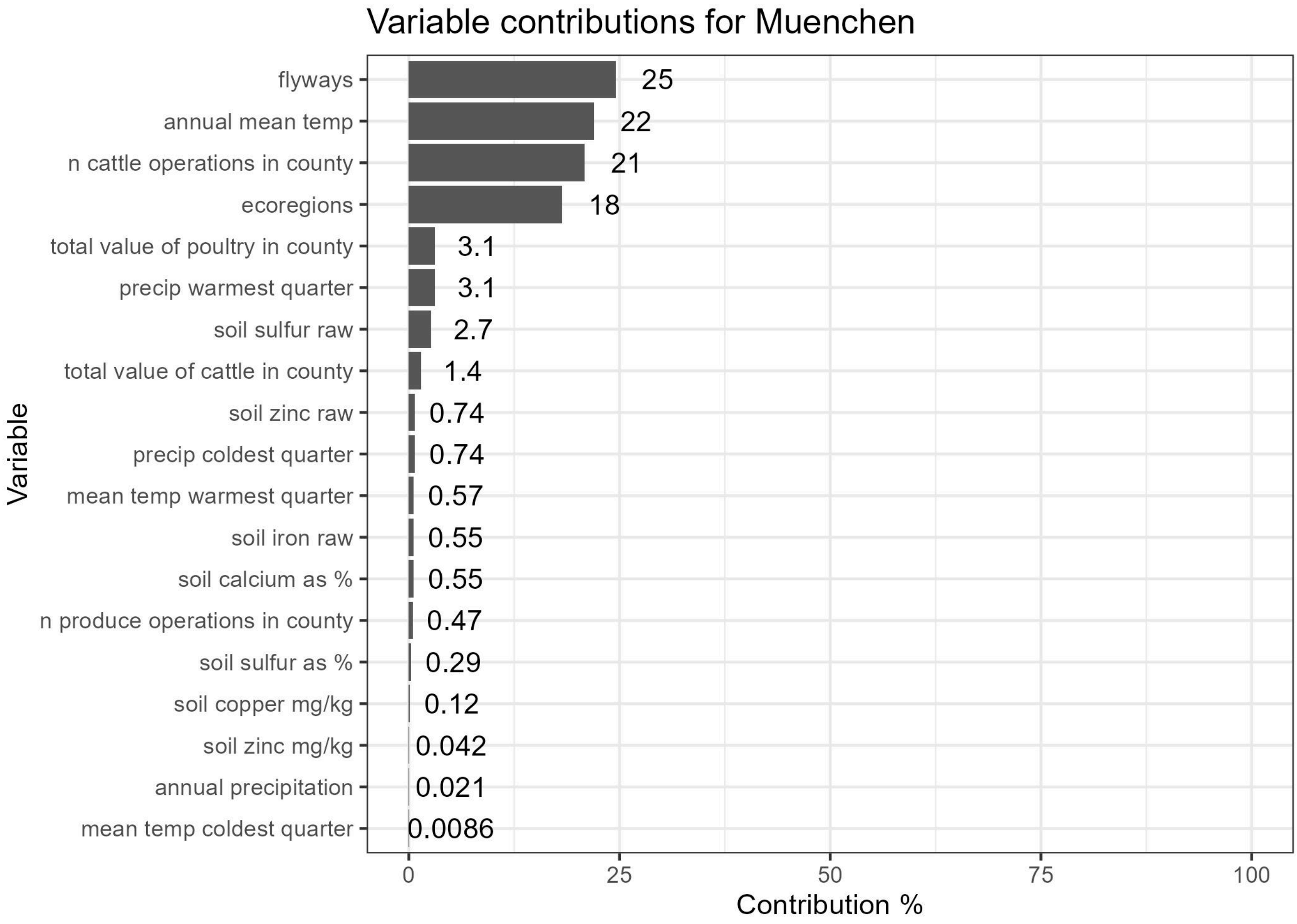


Lambdas for Muenchen. Flyways are indicated as follows: (1) Atlantic, (2) Central, (3) Mississippi, (4) Pacific. See Supplementary File 1 for Ecoregion values.

| Feature | Variable | Type | Lambda | Minimum Encountered Value | Maximum Encountered Value |
|---|---|---|---|---|---|
| ecoregions==6.0 | ecoregions | categorical | 0.0000000 | 0.0000000 | 1.000000e+00 |
| ecoregions==9.0 | ecoregions | categorical | 0.1933363 | 0.0000000 | 1.000000e+00 |
| ecoregions==13.0 | ecoregions | categorical | 0.0584890 | 0.0000000 | 1.000000e+00 |
| ecoregions==23.0 | ecoregions | categorical | 1.6853762 | 0.0000000 | 1.000000e+00 |
| flyways==1.0 | flyways | categorical | 0.0000000 | 0.0000000 | 1.000000e+00 |
| flyways==3.0 | flyways | categorical | -0.8156262 | 0.0000000 | 1.000000e+00 |
| flyways==4.0 | flyways | categorical | -0.0391873 | 0.0000000 | 1.000000e+00 |
| Top5_Ca | soil calcium raw | linear | 0.0000000 | 0.0000000 | 2.550000e+02 |
| Top5_Ca_percWt | soil calcium as % | linear | 0.0000000 | 0.0388513 | 2.004092e+01 |
| Top5_Cu | soil copper raw | linear | 0.0000000 | 0.0000000 | 2.550000e+02 |
| Top5_Cu_mgPerKg | soil copper mg/kg | linear | 0.0000000 | 2.8219459 | 1.143508e+02 |
| Top5_Fe | soil iron raw | linear | 0.0000000 | 0.0000000 | 2.550000e+02 |
| Top5_Fe_percWt | soil iron % | linear | 0.0000000 | 0.1556349 | 6.911553e+00 |
| Top5_K | soil potassium raw | linear | 0.0000000 | 0.0000000 | 2.550000e+02 |

| Feature | Variable | Type | Lambda | Minimum Encountered Value | Maximum Encountered Value |
|---|---|---|---|---|---|
| Top5_K_percWt | soil potassium as % | linear | 0.0000000 | 0.0660172 | 3.244768e+00 |
| Top5_Mn | soil manganese raw | linear | 0.0000000 | 0.0000000 | 2.550000e+02 |
| Top5_Na | soil sodium raw | linear | 0.0000000 | 0.0000000 | 2.550000e+02 |
| Top5_Na_percWt | soil sodium % | linear | 0.0000000 | 0.0260447 | 3.166267e+00 |
| Top5_P | soil phosphorus raw | linear | 0.0000000 | 0.0000000 | 2.550000e+02 |
| Top5_P_mgPerKg | soil phosphorus mg/kg | linear | 0.0000000 | 123.4395752 | 2.347342e+03 |
| Top5_S | soil sulfur raw | linear | 0.0000000 | 0.0000000 | 2.550000e+02 |
| Top5_S_percWt | soil sulfur as % | linear | 0.0000000 | 0.0070236 | 3.381642e-01 |
| Top5_Zn | soil zinc raw | linear | 0.0000000 | 0.0000000 | 2.550000e+02 |
| Top5_Zn_mgPerKg | soil zinc mg/kg | linear | 0.0000000 | 7.5341401 | 4.134917e+02 |
| caco3_kg_sq_m | soil calcium carbonate | linear | 0.0000000 | 0.0000000 | 1.150172e+03 |
| cec_05 | soil cation exchange capacity | linear | 0.0000000 | 0.4358235 | 1.698311e+02 |
| drainage_class_int | soil drainage class | linear | 0.0000000 | 1.0000000 | 8.000000e+00 |
| ec_05 | soil electrical conductivity | linear | 0.0000000 | 0.0000000 | 1.400000e+02 |
| headCattle_nOperations | n cattle operations in county | linear | 0.0000000 | 4.0000000 | 4.000000e+01 |
| headCattle_totalValue | total value of cattle in county | linear | 0.0000000 | 4.0000000 | 4.838209e+06 |
| headOtherLivestock_nOperations | n other livestock operations in county | linear | 0.0000000 | 2.0000000 | 1.900000e+01 |
| headOtherLivestock_totalValue | total value of other livestock in county | linear | 0.0000000 | 2.0000000 | 3.769605e+06 |
| headPoultry_nOperations | n poultry operations in county | linear | 0.0000000 | 1.0000000 | 1.700000e+01 |
| headPoultry_totalValue | total value of poultry in county | linear | 0.0000000 | 1.0000000 | 3.690649e+07 |
| om_kg_sq_m | soil organic matter | linear | 0.0000000 | 0.0000000 | 3.436402e+02 |
| ph_05 | soil pH | linear | 0.0000000 | 3.2314589 | 9.588049e+00 |

| Feature | Variable | Type | Lambda | Minimum Encountered Value | Maximum Encountered Value |
|---|---|---|---|---|---|
| produce_nOperations | n produce operations in county | linear | 0.0000000 | 0.0000000 | 4.100000e+01 |
| texture_05 | soil texture | linear | 0.0000000 | 1.0000000 | 1.200000e+01 |
| wc2.1_2.5m_bio_10 | mean temp warmest quarter | linear | 0.0000000 | 5.6320000 | 3.575067e+01 |
| wc2.1_2.5m_bio_11 | mean temp coldest quarter | linear | 0.0000000 | -14.8253336 | 2.034733e+01 |
| wc2.1_2.5m_bio_12 | annual precipitation | linear | 0.0000000 | 57.0000000 | 3.255000e+03 |
| wc2.1_2.5m_bio_13 | precip wettest month | linear | 0.0000000 | 11.0000000 | 5.360000e+02 |
| wc2.1_2.5m_bio_14 | precip driest month | linear | 0.0000000 | 0.0000000 | 1.460000e+02 |
| wc2.1_2.5m_bio_15 | precip seasonality | linear | 0.0000000 | 5.4388194 | 1.070725e+02 |
| wc2.1_2.5m_bio_16 | precip wettest quarter | linear | 0.0000000 | 27.0000000 | 1.457000e+03 |
| wc2.1_2.5m_bio_17 | precip driest quarter | linear | 0.0000000 | 2.0000000 | 4.590000e+02 |
| wc2.1_2.5m_bio_18 | precip warmest quarter | linear | 0.0000000 | 2.0000000 | 6.400000e+02 |
| wc2.1_2.5m_bio_19 | precip coldest quarter | linear | 0.0000000 | 16.0000000 | 1.386000e+03 |
| wc2.1_2.5m_bio_1 | annual mean temp | linear | 0.0000000 | -3.1535001 | 2.447950e+01 |
| wc2.1_2.5m_bio_2 | mean diurnal range | linear | 0.0000000 | 6.2884617 | 2.171233e+01 |
| wc2.1_2.5m_bio_3 | isothermality | linear | 0.0000000 | 21.9432907 | 6.109463e+01 |
| wc2.1_2.5m_bio_4 | temperature seasonality | linear | 0.0000000 | 229.3383636 | 1.363949e+03 |
| wc2.1_2.5m_bio_5 | max temp warmest month | linear | 0.0000000 | 13.7679996 | 4.514800e+01 |
| wc2.1_2.5m_bio_6 | min temp coldest month | linear | 0.0000000 | -23.3279991 | 1.512800e+01 |
| wc2.1_2.5m_bio_7 | annual temp range | linear | 0.0000000 | 14.3642864 | 4.984400e+01 |
| wc2.1_2.5m_bio_8 | mean temp wettest quarter | linear | 0.0000000 | -8.9613333 | 3.345667e+01 |
| wc2.1_2.5m_bio_9 | mean temp driest quarter | linear | 0.0000000 | -14.8253336 | 2.854333e+01 |
| Top5_Cu_mgPerKg^2 | soil copper mg/kg | quadratic | -2.9117480 | 7.9633787 | 1.307611e+04 |
| Top5_Zn_mgPerKg^2 | soil zinc mg/kg | quadratic | -1.8734484 | 56.7632672 | 1.709754e+05 |

| Feature | Variable | Type | Lambda | Minimum Encountered Value | Maximum Encountered Value |
|---|---|---|---|---|---|
| Top5_Ca_percWt*Top5_Fe | soil calcium as %,soil iron raw | product | -0.3261250 | 0.0000000 | 3.035049e+03 |
| Top5_S*headCattle_totalValue | soil sulfur raw,total value of cattle in county | product | 0.1320118 | 0.0000000 | 1.233743e+09 |
| Top5_S*wc2.1_2.5m_bio_1 | soil sulfur raw,annual mean temp | product | 0.5888965 | -804.1425204 | 6.242272e+03 |
| Top5_Zn*wc2.1_2.5m_bio_19 | soil zinc raw,precip coldest quarter | product | -1.1384437 | 0.0000000 | 3.271650e+05 |
| headCattle_nOperations*wc2.1_2.5m_bio_10 | n cattle operations in county,mean temp warmest quarter | product | 1.6028270 | 63.7040024 | 1.131614e+03 |
| headCattle_nOperations*wc2.1_2.5m_bio_1 | n cattle operations in county,annual mean temp | product | 0.7918955 | -107.2190027 | 7.640710e+02 |
| headCattle_totalValue*wc2.1_2.5m_bio_12 | total value of cattle in county,annual precipitation | product | 0.7550555 | 1940.0000000 | 3.396423e+09 |
| headPoultry_totalValue*wc2.1_2.5m_bio_18 | total value of poultry in county,precip warmest quarter | product | -5.4840316 | 24.0000000 | 1.147792e+10 |
| produce_nOperations*wc2.1_2.5m_bio_1 | n produce operations in county,annual mean temp | product | 0.8394491 | -47.3025012 | 6.930033e+02 |

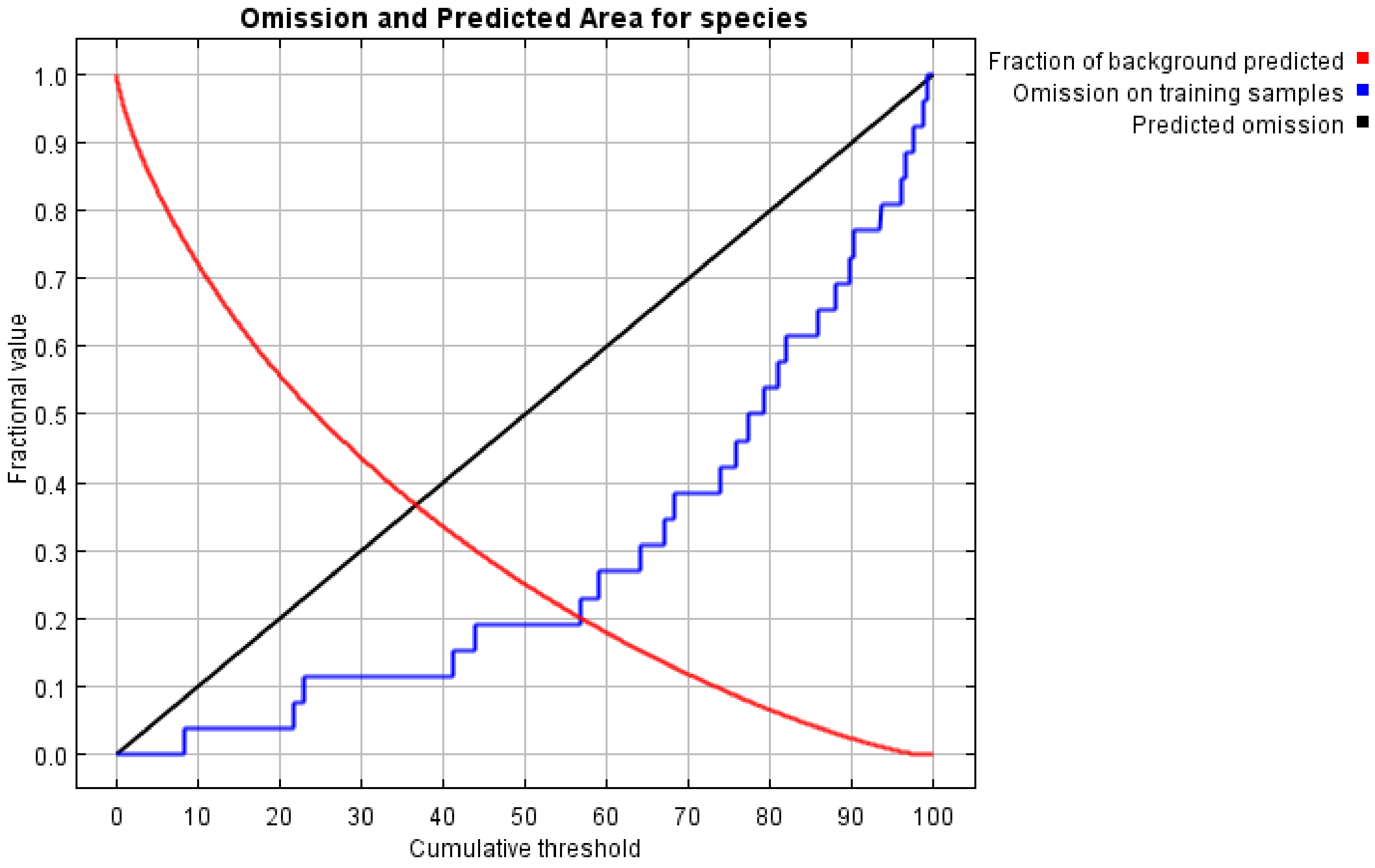
Omission and Predicted Area for species
Fraction of background predicted
Omission on training samples
Predicted omission
Fractional value
Cumulative threshold
0.0
0.1
0.2
0.3
0.4
0.5
0.6
0.7
0.8
0.9
1.0
0
10
20
30
40
50
60
70
80
90
100

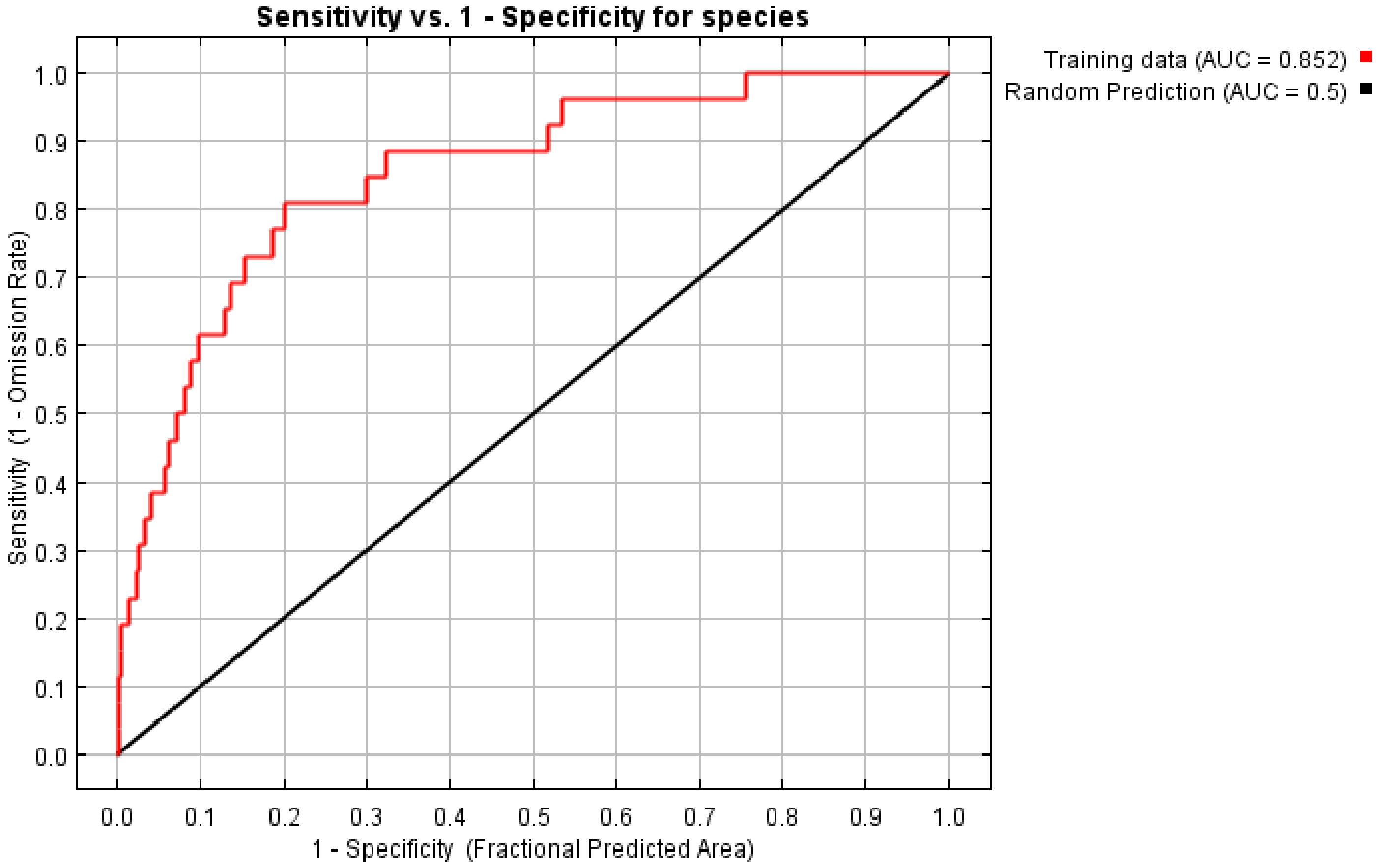
Sensitivity vs. 1 - Specificity for species
Training data (AUC = 0.852)
Random Prediction (AUC = 0.5)
Sensitivity (1 - Omission Rate)
1 - Specificity (Fractional Predicted Area)
0.0
0.1
0.2
0.3
0.4
0.5
0.6
0.7
0.8
0.9
1.0
