## Supplementary File 15 for "Geographic Distributions of Top Beef Salmonella Serovars in the U.S."

### Species Distribution Model Results for Serovar Newport

Model tuning results for Newport.

| N Points | N Training Points | N Background Points | Regularization Multiplier | Features | Training AUC |
|---:|---:|---:|---:|---|---:|
| 47 | 18 | 10016 | 3.5 | LQPT | 0.86 |

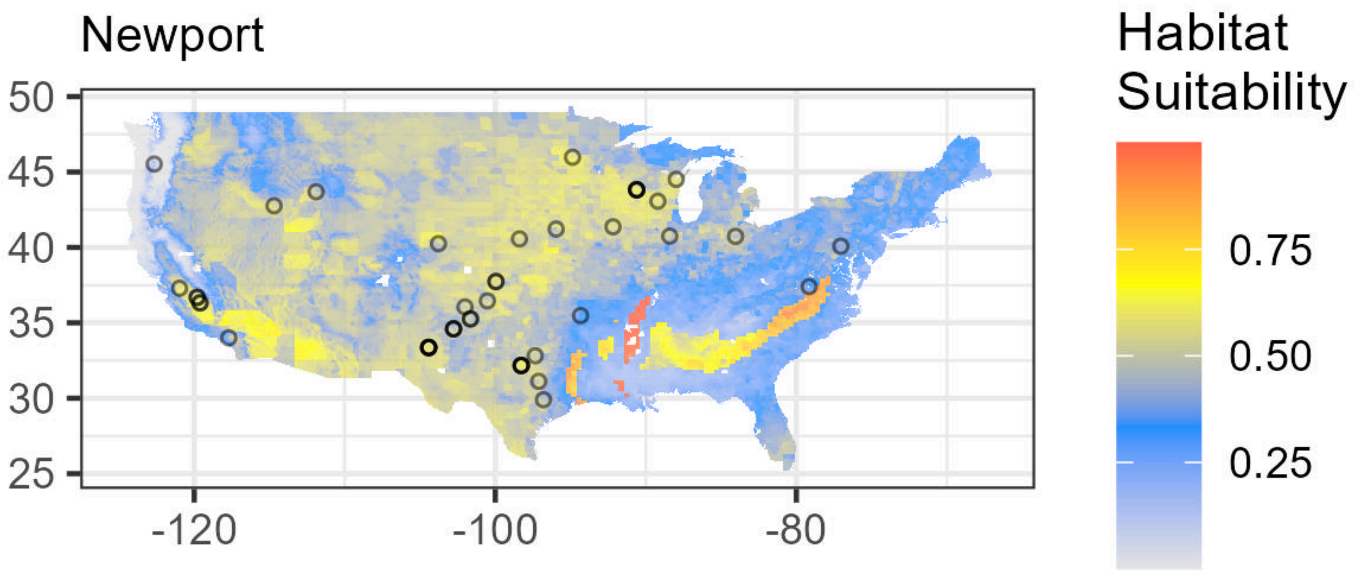

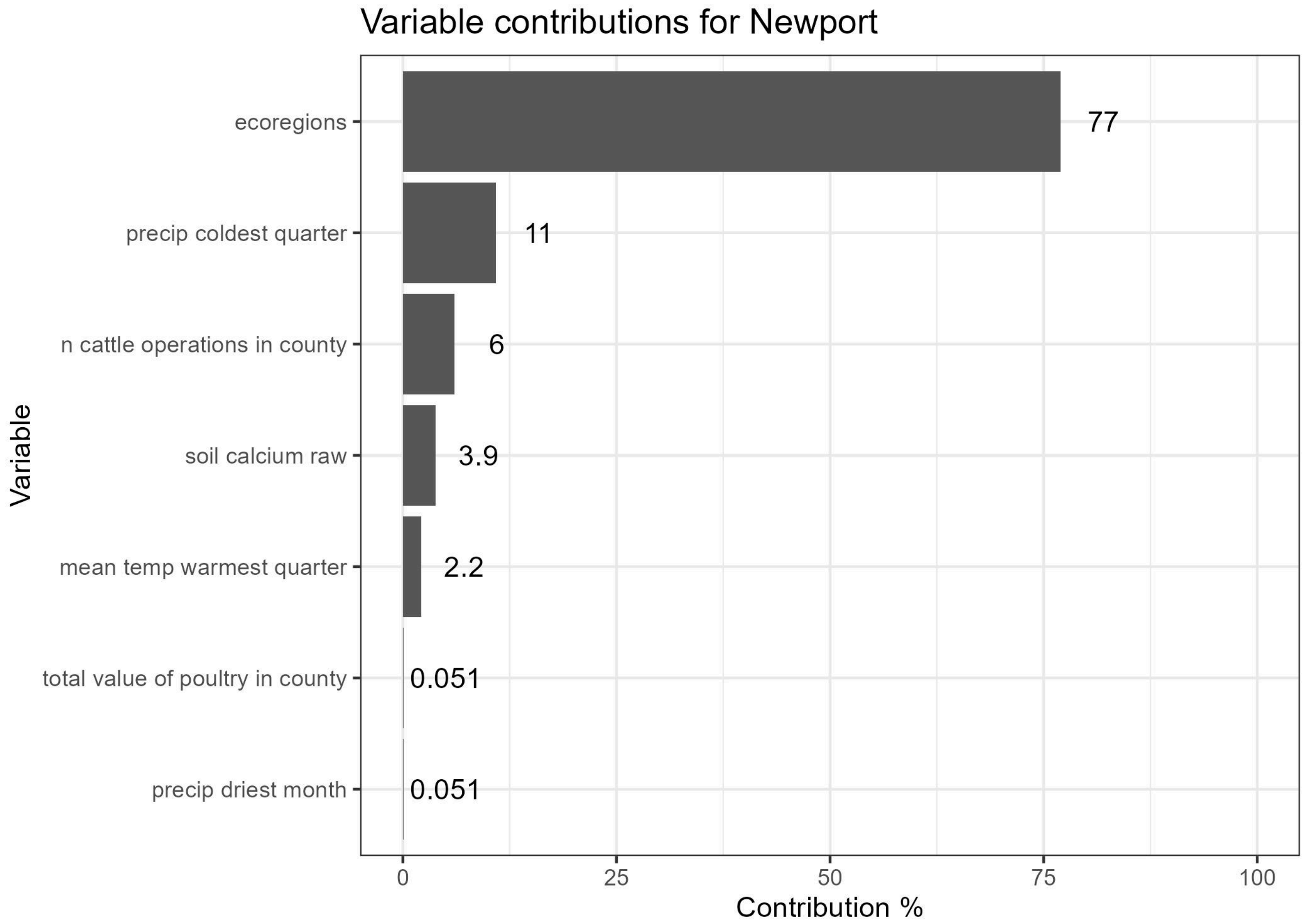


Lambdas for Newport. Flyways are indicated as follows: (1) Atlantic, (2) Central, (3) Mississippi, (4) Pacific. See Supplementary File 1 for Ecoregion values.

| Feature | Variable | Type | Lambda | Minimum Encountered Value | Maximum Encountered Value |
|---|---|---|---|---|---|
| ecoregions==6.0 | ecoregions | categorical | 0.0000000 | 0.0000000 | 1.000000e+00 |
| ecoregions==10.0 | ecoregions | categorical | 0.5338972 | 0.0000000 | 1.000000e+00 |
| ecoregions==22.0 | ecoregions | categorical | 1.6174987 | 0.0000000 | 1.000000e+00 |
| ecoregions==23.0 | ecoregions | categorical | 3.0531068 | 0.0000000 | 1.000000e+00 |
| ecoregions==32.0 | ecoregions | categorical | 0.3087582 | 0.0000000 | 1.000000e+00 |
| ecoregions==40.0 | ecoregions | categorical | -0.0803843 | 0.0000000 | 1.000000e+00 |
| flyways==1.0 | flyways | categorical | 0.0000000 | 0.0000000 | 1.000000e+00 |
| Top5_Ca | soil calcium raw | linear | 0.0000000 | 0.0000000 | 2.550000e+02 |
| Top5_Ca_percWt | soil calcium as % | linear | 0.0000000 | 0.0685176 | 2.048439e+01 |
| Top5_Cu | soil copper raw | linear | 0.0000000 | 0.0000000 | 2.550000e+02 |
| Top5_Cu_mgPerKg | soil copper mg/kg | linear | 0.0000000 | 2.7762976 | 9.474970e+01 |
| Top5_Fe | soil iron raw | linear | 0.0000000 | 0.0000000 | 2.550000e+02 |
| Top5_Fe_percWt | soil iron % | linear | 0.0000000 | 0.1629301 | 6.979847e+00 |
| Top5_K | soil potassium raw | linear | 0.0000000 | 0.0000000 | 2.550000e+02 |

| Feature | Variable | Type | Lambda | Minimum Encountered Value | Maximum Encountered Value |
|---|---|---|---|---|---|
| Top5_K_percWt | soil potassium as % | linear | 0.0000000 | 0.0677590 | 3.326587e+00 |
| Top5_Mn | soil manganese raw | linear | 0.0000000 | 0.0000000 | 2.550000e+02 |
| Top5_Na | soil sodium raw | linear | 0.0000000 | 0.0000000 | 2.550000e+02 |
| Top5_Na_percWt | soil sodium % | linear | 0.0000000 | 0.0225875 | 3.196614e+00 |
| Top5_P | soil phosphorus raw | linear | 0.0000000 | 0.0000000 | 2.550000e+02 |
| Top5_P_mgPerKg | soil phosphorus mg/kg | linear | 0.0000000 | 123.9033432 | 2.232643e+03 |
| Top5_S | soil sulfur raw | linear | 0.0000000 | 0.0000000 | 2.550000e+02 |
| Top5_S_percWt | soil sulfur as % | linear | 0.0000000 | 0.0069159 | 4.180052e-01 |
| Top5_Zn | soil zinc raw | linear | 0.0000000 | 0.0000000 | 2.550000e+02 |
| Top5_Zn_mgPerKg | soil zinc mg/kg | linear | 0.0000000 | 6.3626819 | 3.677134e+02 |
| caco3_kg_sq_m | soil calcium carbonate | linear | 0.0000000 | 0.0000000 | 1.082013e+03 |
| cec_05 | soil cation exchange capacity | linear | 0.0000000 | 0.4987435 | 1.546882e+02 |
| drainage_class_int | soil drainage class | linear | 0.0000000 | 1.0000000 | 8.000000e+00 |
| ec_05 | soil electrical conductivity | linear | 0.0000000 | 0.0000000 | 1.400000e+02 |
| headCattle_nOperations | n cattle operations in county | linear | 0.0000000 | 4.0000000 | 4.000000e+01 |
| headCattle_totalValue | total value of cattle in county | linear | 0.0000000 | 4.0000000 | 4.838209e+06 |
| headOtherLivestock_nOperations | n other livestock operations in county | linear | 0.0000000 | 2.0000000 | 1.900000e+01 |
| headOtherLivestock_totalValue | total value of other livestock in county | linear | 0.0000000 | 2.0000000 | 3.769605e+06 |
| headPoultry_nOperations | n poultry operations in county | linear | 0.0000000 | 1.0000000 | 1.700000e+01 |
| headPoultry_totalValue | total value of poultry in county | linear | 0.0000000 | 1.0000000 | 3.690649e+07 |
| om_kg_sq_m | soil organic matter | linear | 0.0000000 | 0.0112372 | 3.414911e+02 |
| ph_05 | soil pH | linear | 0.0000000 | 3.5535202 | 9.500000e+00 |

| Feature | Variable | Type | Lambda | Minimum Encountered Value | Maximum Encountered Value |
|---|---|---|---|---|---|
| produce_nOperations | n produce operations in county | linear | 0.0000000 | 0.0000000 | 4.000000e+01 |
| texture_05 | soil texture | linear | 0.0000000 | 1.0000000 | 1.200000e+01 |
| wc2.1_2.5m_bio_10 | mean temp warmest quarter | linear | 0.0000000 | 6.4179997 | 3.463667e+01 |
| wc2.1_2.5m_bio_11 | mean temp coldest quarter | linear | 0.0000000 | -15.1086664 | 2.038478e+01 |
| wc2.1_2.5m_bio_12 | annual precipitation | linear | 0.0000000 | 58.0000000 | 3.344000e+03 |
| wc2.1_2.5m_bio_13 | precip wettest month | linear | 0.0000000 | 10.0000000 | 5.490000e+02 |
| wc2.1_2.5m_bio_14 | precip driest month | linear | 0.0000000 | 0.0000000 | 1.530000e+02 |
| wc2.1_2.5m_bio_15 | precip seasonality | linear | 0.0000000 | 5.5186062 | 9.482897e+01 |
| wc2.1_2.5m_bio_16 | precip wettest quarter | linear | 0.0000000 | 27.0000000 | 1.494000e+03 |
| wc2.1_2.5m_bio_17 | precip driest quarter | linear | 0.0000000 | 2.0000000 | 4.760000e+02 |
| wc2.1_2.5m_bio_18 | precip warmest quarter | linear | 0.0000000 | 2.0000000 | 6.390000e+02 |
| wc2.1_2.5m_bio_19 | precip coldest quarter | linear | 0.0000000 | 16.0000000 | 1.423000e+03 |
| wc2.1_2.5m_bio_1 | annual mean temp | linear | 0.0000000 | -2.7510002 | 2.448062e+01 |
| wc2.1_2.5m_bio_2 | mean diurnal range | linear | 0.0000000 | 6.6294870 | 2.147567e+01 |
| wc2.1_2.5m_bio_3 | isothermality | linear | 0.0000000 | 23.7819824 | 6.146171e+01 |
| wc2.1_2.5m_bio_4 | temperature seasonality | linear | 0.0000000 | 254.2902832 | 1.356662e+03 |
| wc2.1_2.5m_bio_5 | max temp warmest month | linear | 0.0000000 | 14.8160000 | 4.488800e+01 |
| wc2.1_2.5m_bio_6 | min temp coldest month | linear | 0.0000000 | -23.9480000 | 1.523913e+01 |
| wc2.1_2.5m_bio_7 | annual temp range | linear | 0.0000000 | 16.6437511 | 4.934800e+01 |
| wc2.1_2.5m_bio_8 | mean temp wettest quarter | linear | 0.0000000 | -8.9813337 | 3.347866e+01 |
| wc2.1_2.5m_bio_9 | mean temp driest quarter | linear | 0.0000000 | -14.7886667 | 2.847267e+01 |
| wc2.1_2.5m_bio_19^2 | precip coldest quarter | quadratic | -12.7419168 | 256.0000000 | 2.024929e+06 |

| Feature | Variable | Type | Lambda | Minimum Encountered Value | Maximum Encountered Value |
|---|---|---|---|---|---|
| Top5_Ca*headCattle_nOperations | soil calcium raw,n cattle operations in county | product | 0.3908168 | 0.0000000 | 1.020000e+04 |
| headCattle_nOperations*wc2.1_2.5m_bio_10 | n cattle operations in county,mean temp warmest quarter | product | 1.0526051 | 62.6160011 | 1.140876e+03 |
| headPoultry_totalValue*wc2.1_2.5m_bio_14 | total value of poultry in county,precip driest month | product | -0.6973891 | 0.0000000 | 2.989426e+09 |

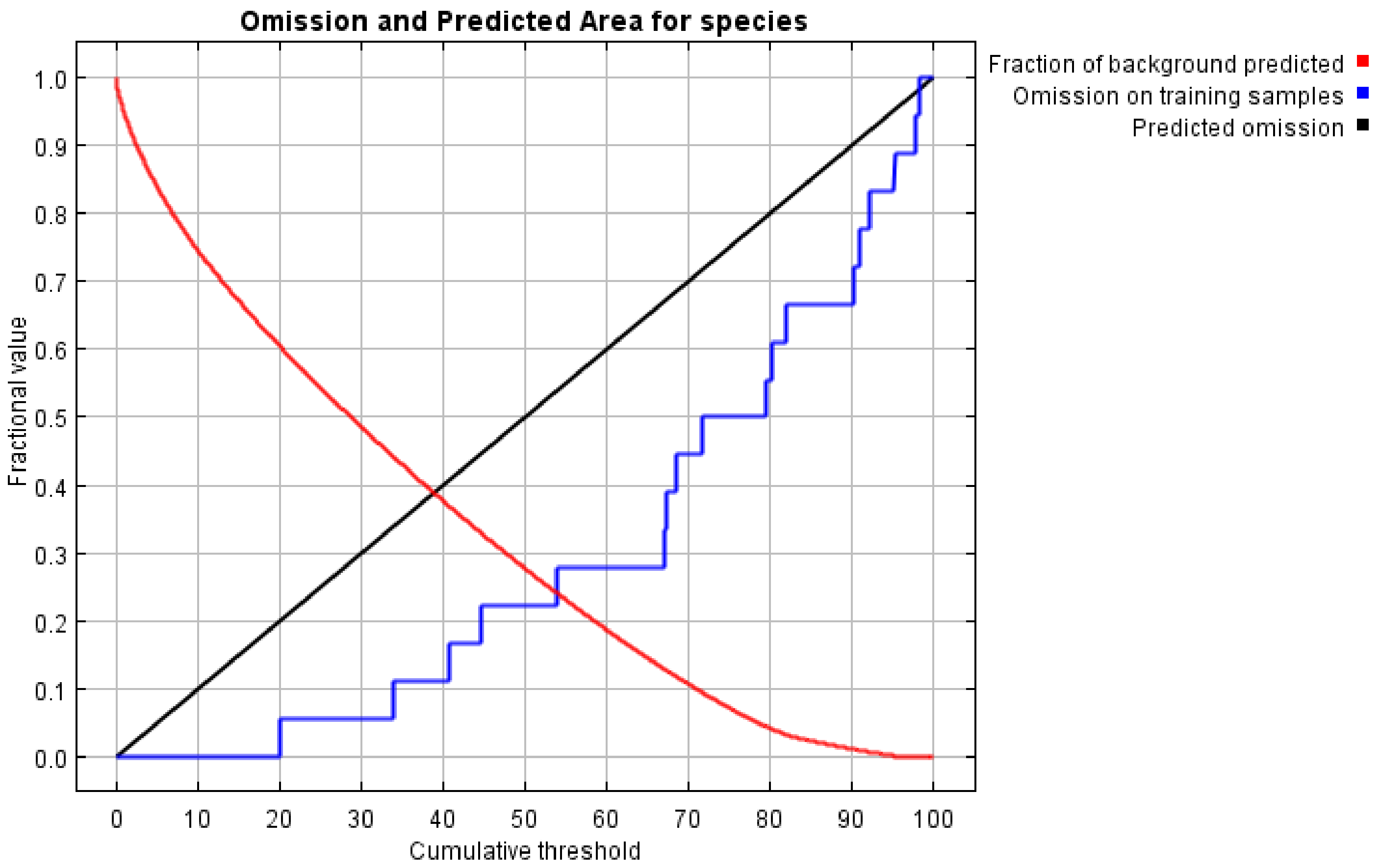

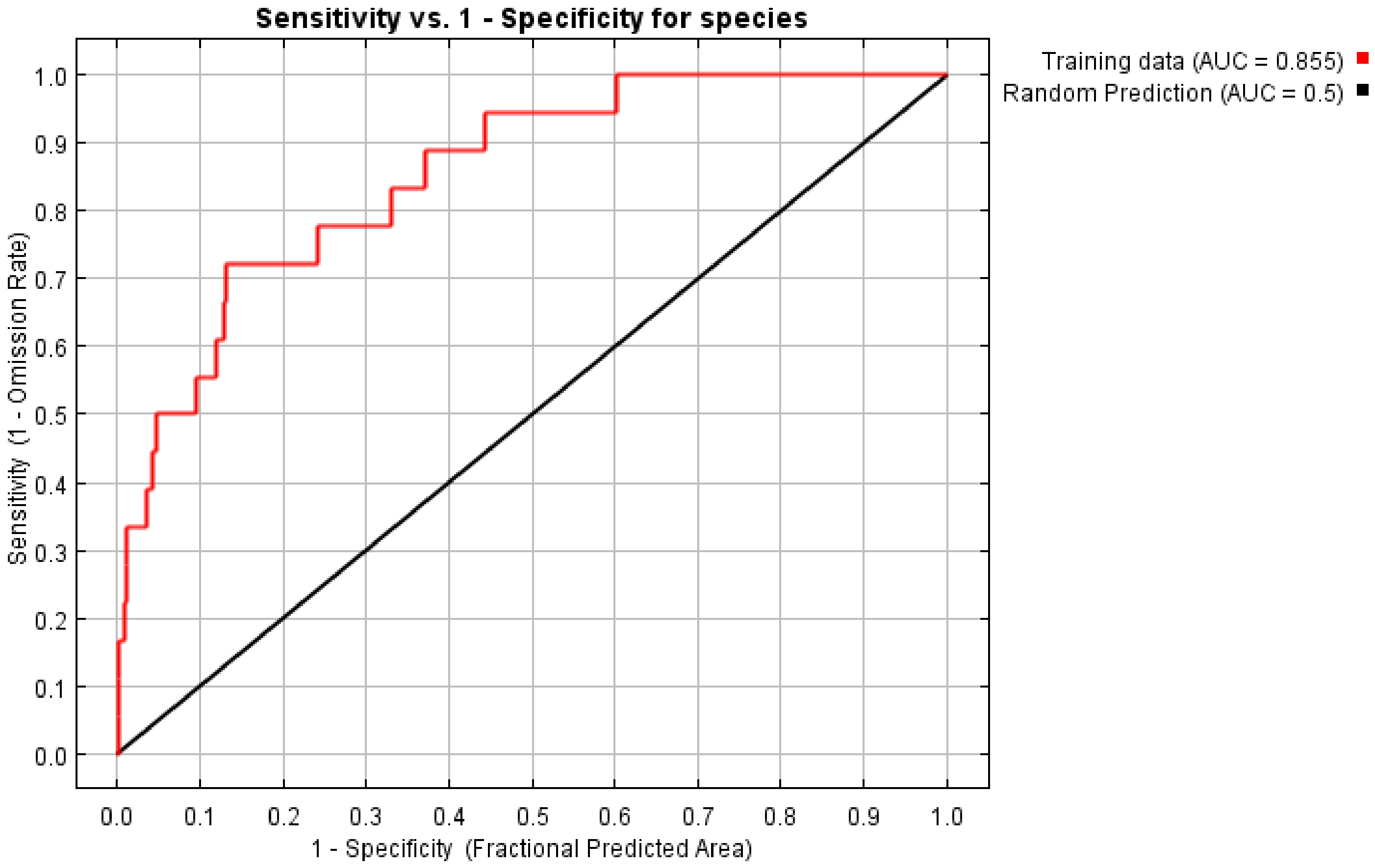
Sensitivity vs. 1 - Specificity for species
Training data (AUC = 0.855)
Random Prediction (AUC = 0.5)
Sensitivity (1 - Omission Rate)
1 - Specificity (Fractional Predicted Area)
0.0
0.1
0.2
0.3
0.4
0.5
0.6
0.7
0.8
0.9
1.0
