## Supplementary File 16 for "Geographic Distributions of Top Beef Salmonella Serovars in the U.S."

### Species Distribution Model Results for Serovar Typhimurium

Model tuning results for Typhimurium.

| N Points | N Training Points | N Background Points | Regularization Multiplier | Features | Training AUC |
|---|---|---|---|---|---|
| 45 | 23 | 10022 | 4 | LQPT | 0.84 |

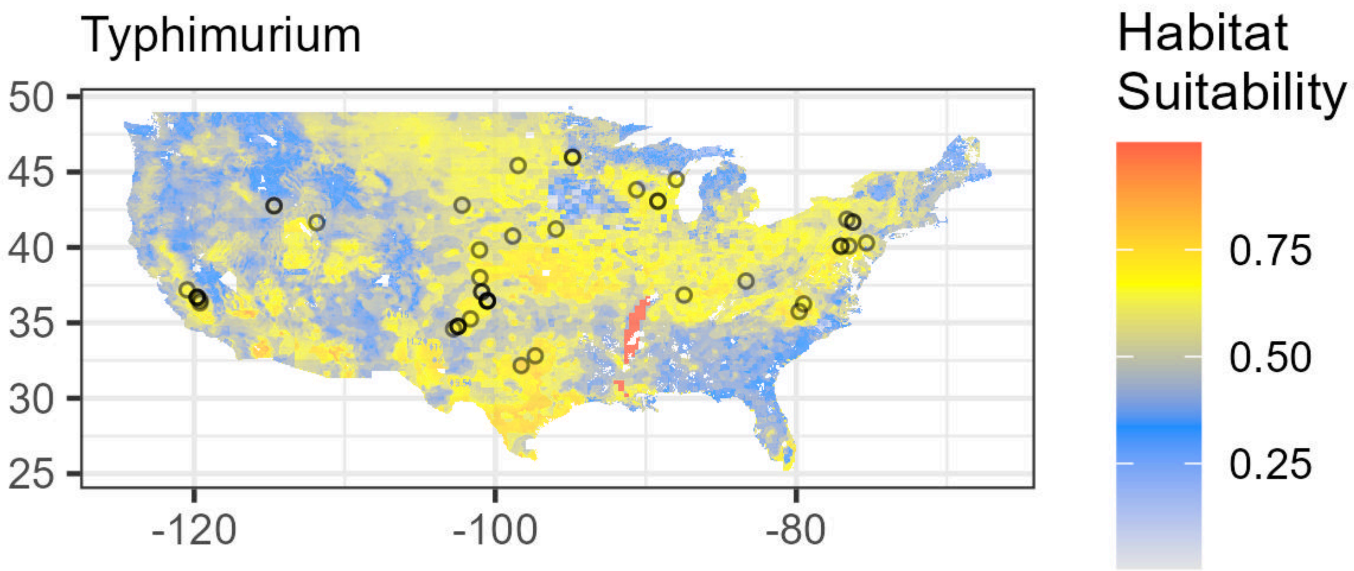

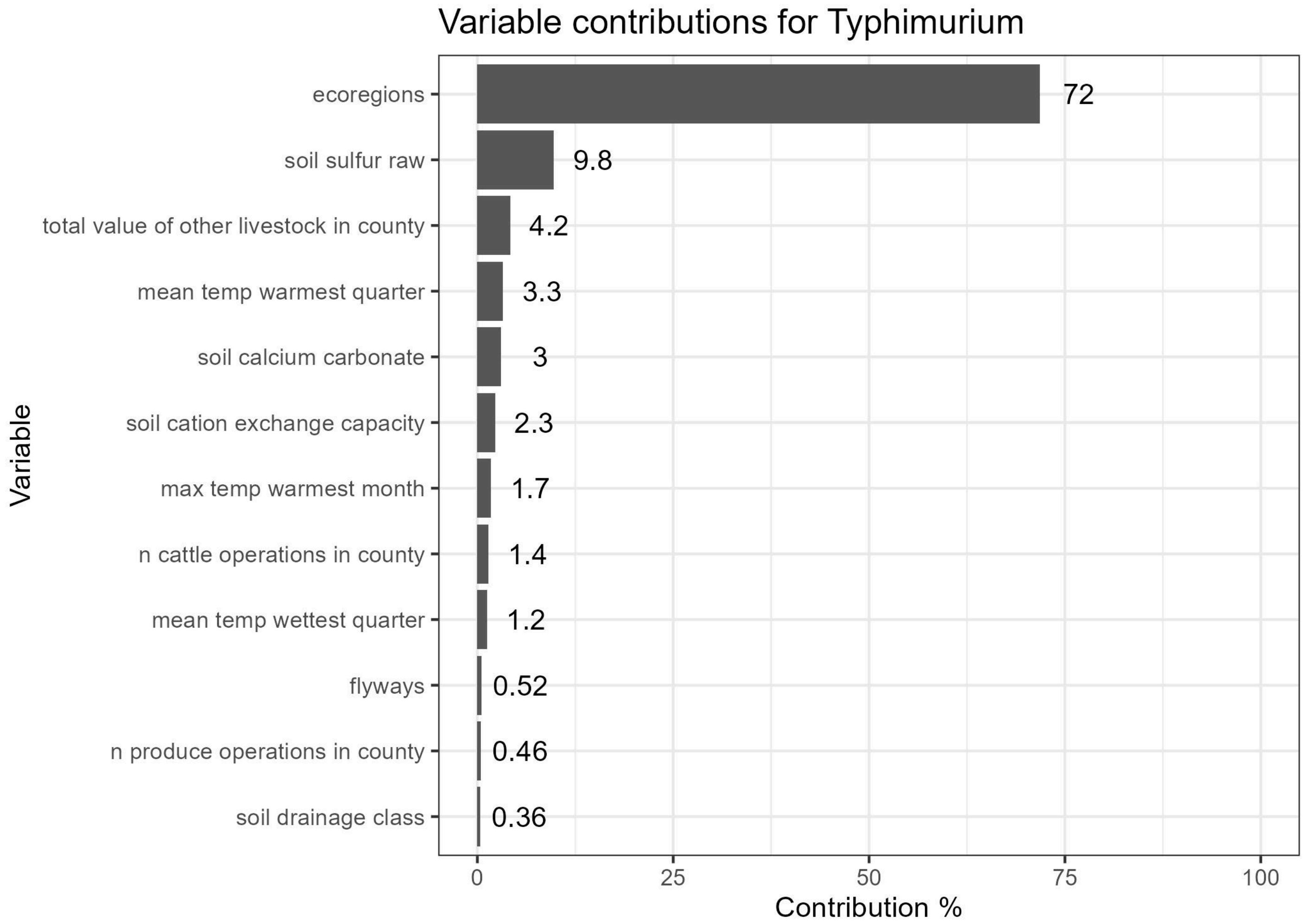


Lambdas for Typhimurium. Flyways are indicated as follows: (1) Atlantic, (2) Central, (3) Mississippi, (4) Pacific. See Supplementary File 1 for Ecoregion values.

| Feature | Variable | Type | Lambda | Minimum Encountered Value | Maximum Encountered Value |
|---|---|---|---|---|---|
| ecoregions==6.0 | ecoregions | categorical | 0.0000000 | 0.0000000 | 1.000000e+00 |
| ecoregions==21.0 | ecoregions | categorical | -0.0872538 | 0.0000000 | 1.000000e+00 |
| ecoregions==23.0 | ecoregions | categorical | 2.8397471 | 0.0000000 | 1.000000e+00 |
| flyways==1.0 | flyways | categorical | 0.0000000 | 0.0000000 | 1.000000e+00 |
| Top5_Ca | soil calcium raw | linear | 0.0000000 | 0.0000000 | 2.550000e+02 |
| Top5_Ca_percWt | soil calcium as % | linear | 0.0000000 | 0.0754027 | 2.008226e+01 |
| Top5_Cu | soil copper raw | linear | 0.0000000 | 0.0000000 | 2.550000e+02 |
| Top5_Cu_mgPerKg | soil copper mg/kg | linear | 0.0000000 | 2.7817333 | 9.932500e+01 |
| Top5_Fe | soil iron raw | linear | 0.0000000 | 0.0000000 | 2.550000e+02 |
| Top5_Fe_percWt | soil iron % | linear | 0.0000000 | 0.1390035 | 7.231884e+00 |
| Top5_K | soil potassium raw | linear | 0.0000000 | 0.0000000 | 2.550000e+02 |
| Top5_K_percWt | soil potassium as % | linear | 0.0000000 | 0.0600070 | 3.489706e+00 |

| Feature | Variable | Type | Lambda | Minimum Encountered Value | Maximum Encountered Value |
|---|---|---|---|---|---|
| Top5_Mn | soil manganese raw | linear | 0.0000000 | 0.0000000 | 2.550000e+02 |
| Top5_Na | soil sodium raw | linear | 0.0000000 | 0.0000000 | 2.550000e+02 |
| Top5_Na_percWt | soil sodium % | linear | 0.0000000 | 0.0243300 | 3.002722e+00 |
| Top5_P | soil phosphorus raw | linear | 0.0000000 | 0.0000000 | 2.550000e+02 |
| Top5_P_mgPerKg | soil phosphorus mg/kg | linear | 0.0000000 | 110.0913849 | 2.417007e+03 |
| Top5_S | soil sulfur raw | linear | 0.0000000 | 0.0000000 | 2.550000e+02 |
| Top5_S_percWt | soil sulfur as % | linear | 0.0000000 | 0.0071759 | 3.793745e-01 |
| Top5_Zn | soil zinc raw | linear | 0.0000000 | 0.0000000 | 2.550000e+02 |
| Top5_Zn_mgPerKg | soil zinc mg/kg | linear | 0.0000000 | 5.8781433 | 3.877300e+02 |
| caco3_kg_sq_m | soil calcium carbonate | linear | 0.0000000 | 0.0000000 | 1.111125e+03 |
| cec_05 | soil cation exchange capacity | linear | 0.0000000 | 0.1115968 | 1.825905e+02 |
| drainage_class_int | soil drainage class | linear | 0.0000000 | 1.0000000 | 8.000000e+00 |
| ec_05 | soil electrical conductivity | linear | 0.0000000 | 0.0000000 | 1.400000e+02 |
| headCattle_nOperations | n cattle operations in county | linear | 0.0000000 | 4.0000000 | 4.000000e+01 |
| headCattle_totalValue | total value of cattle in county | linear | 0.0000000 | 4.0000000 | 4.838209e+06 |
| headOtherLivestock_nOperations | n other livestock operations in county | linear | 0.0000000 | 2.0000000 | 1.900000e+01 |
| headOtherLivestock_totalValue | total value of other livestock in county | linear | 0.0000000 | 2.0000000 | 3.769605e+06 |
| headPoultry_nOperations | n poultry operations in county | linear | 0.0000000 | 1.0000000 | 1.700000e+01 |
| headPoultry_totalValue | total value of poultry in county | linear | 0.0000000 | 1.0000000 | 3.690649e+07 |
| om_kg_sq_m | soil organic matter | linear | 0.0000000 | 0.0000000 | 2.980959e+02 |
| ph_05 | soil pH | linear | 0.0000000 | 3.2245059 | 9.500000e+00 |
| produce_nOperations | n produce operations in county | linear | 0.0000000 | 0.0000000 | 4.100000e+01 |

| Feature | Variable | Type | Lambda | Minimum Encountered Value | Maximum Encountered Value |
|---|---|---|---|---|---|
| texture_05 | soil texture | linear | 0.0000000 | 1.0000000 | 1.200000e+01 |
| wc2.1_2.5m_bio_10 | mean temp warmest quarter | linear | 0.0000000 | 5.1506667 | 3.547667e+01 |
| wc2.1_2.5m_bio_11 | mean temp coldest quarter | linear | 0.0000000 | -14.8226671 | 2.051600e+01 |
| wc2.1_2.5m_bio_12 | annual precipitation | linear | 0.0000000 | 60.0000000 | 3.363000e+03 |
| wc2.1_2.5m_bio_13 | precip wettest month | linear | 0.0000000 | 10.0000000 | 5.440000e+02 |
| wc2.1_2.5m_bio_14 | precip driest month | linear | 0.0000000 | 0.0000000 | 1.440000e+02 |
| wc2.1_2.5m_bio_15 | precip seasonality | linear | 0.0000000 | 5.3961720 | 9.455029e+01 |
| wc2.1_2.5m_bio_16 | precip wettest quarter | linear | 0.0000000 | 27.0000000 | 1.494000e+03 |
| wc2.1_2.5m_bio_17 | precip driest quarter | linear | 0.0000000 | 2.0000000 | 4.560000e+02 |
| wc2.1_2.5m_bio_18 | precip warmest quarter | linear | 0.0000000 | 2.0000000 | 6.470000e+02 |
| wc2.1_2.5m_bio_19 | precip coldest quarter | linear | 0.0000000 | 16.0000000 | 1.425000e+03 |
| wc2.1_2.5m_bio_1 | annual mean temp | linear | 0.0000000 | -3.6818333 | 2.463317e+01 |
| wc2.1_2.5m_bio_2 | mean diurnal range | linear | 0.0000000 | 6.6750002 | 2.182900e+01 |
| wc2.1_2.5m_bio_3 | isothermality | linear | 0.0000000 | 22.6988010 | 6.472662e+01 |
| wc2.1_2.5m_bio_4 | temperature seasonality | linear | 0.0000000 | 217.3670959 | 1.357699e+03 |
| wc2.1_2.5m_bio_5 | max temp warmest month | linear | 0.0000000 | 12.2080002 | 4.544400e+01 |
| wc2.1_2.5m_bio_6 | min temp coldest month | linear | 0.0000000 | -23.3400002 | 1.549565e+01 |
| wc2.1_2.5m_bio_7 | annual temp range | linear | 0.0000000 | 15.0636368 | 4.959200e+01 |
| wc2.1_2.5m_bio_8 | mean temp wettest quarter | linear | 0.0000000 | -8.5446663 | 3.347600e+01 |
| wc2.1_2.5m_bio_9 | mean temp driest quarter | linear | 0.0000000 | -14.8226671 | 2.837667e+01 |
| cec_05^2 | soil cation exchange capacity | quadratic | -1.6805272 | 0.0124538 | 3.333930e+04 |
| Top5_S*produce_nOperations | soil sulfur raw,n produce operations in county | product | 0.1623617 | 0.0000000 | 1.045500e+04 |

| Feature | Variable | Type | Lambda | Minimum Encountered Value | Maximum Encountered Value |
|---|---|---|---|---|---|
| Top5_S*wc2.1_2.5m_bio_10 | soil sulfur raw,mean temp warmest quarter | product | 0.5762508 | 0.0000000 | 9.046550e+03 |
| Top5_S*wc2.1_2.5m_bio_8 | soil sulfur raw,mean temp wettest quarter | product | 0.5336478 | -2178.8899040 | 8.116480e+03 |
| cec_05*headOtherLivestock_totalValue | soil cation exchange capacity,total value of other livestock in county | product | -4.8801525 | 1.9992802 | 8.089056e+07 |
| headCattle_nOperations*wc2.1_2.5m_bio_10 | n cattle operations in county,mean temp warmest quarter | product | 1.1243384 | 63.3973312 | 1.113948e+03 |

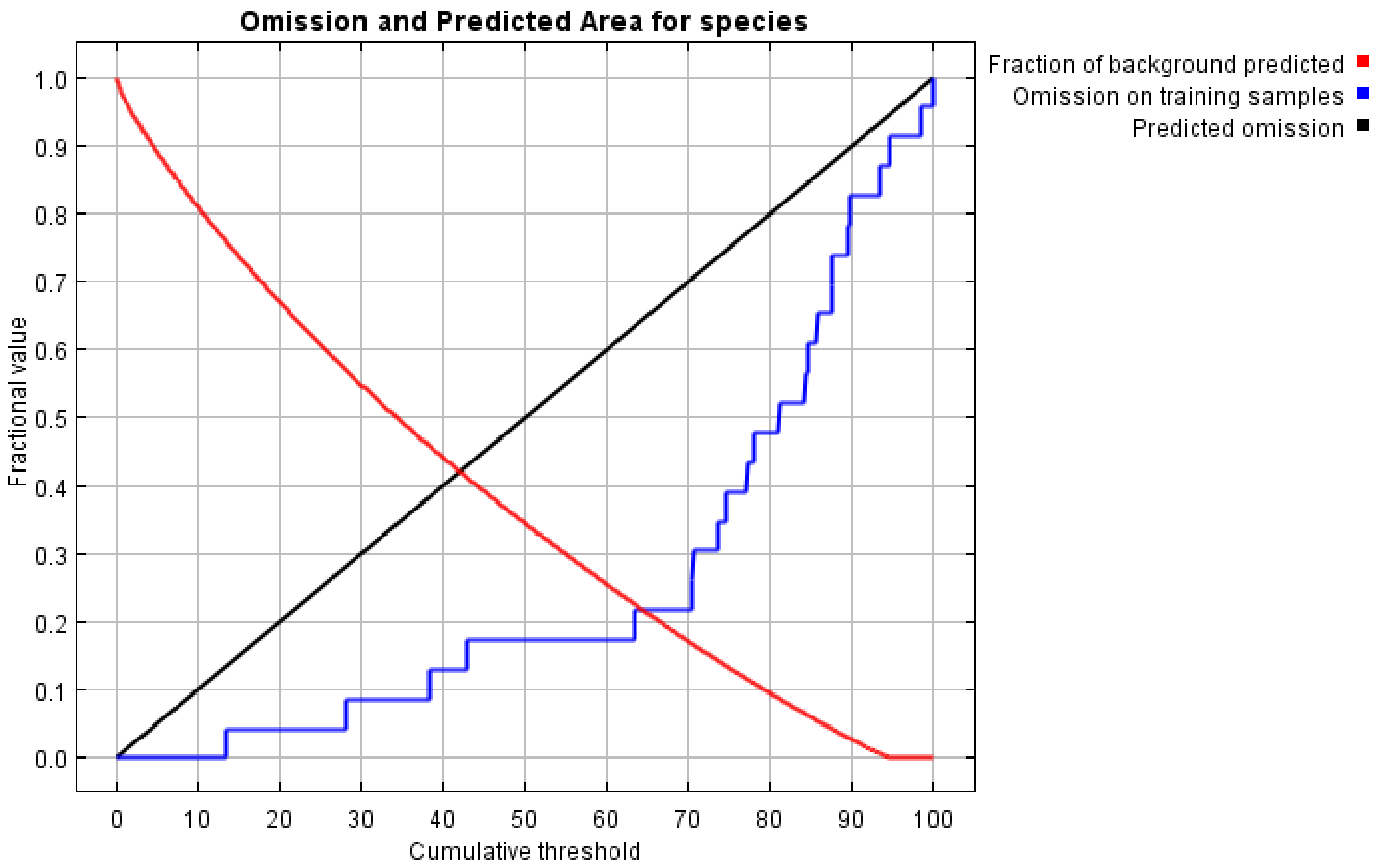

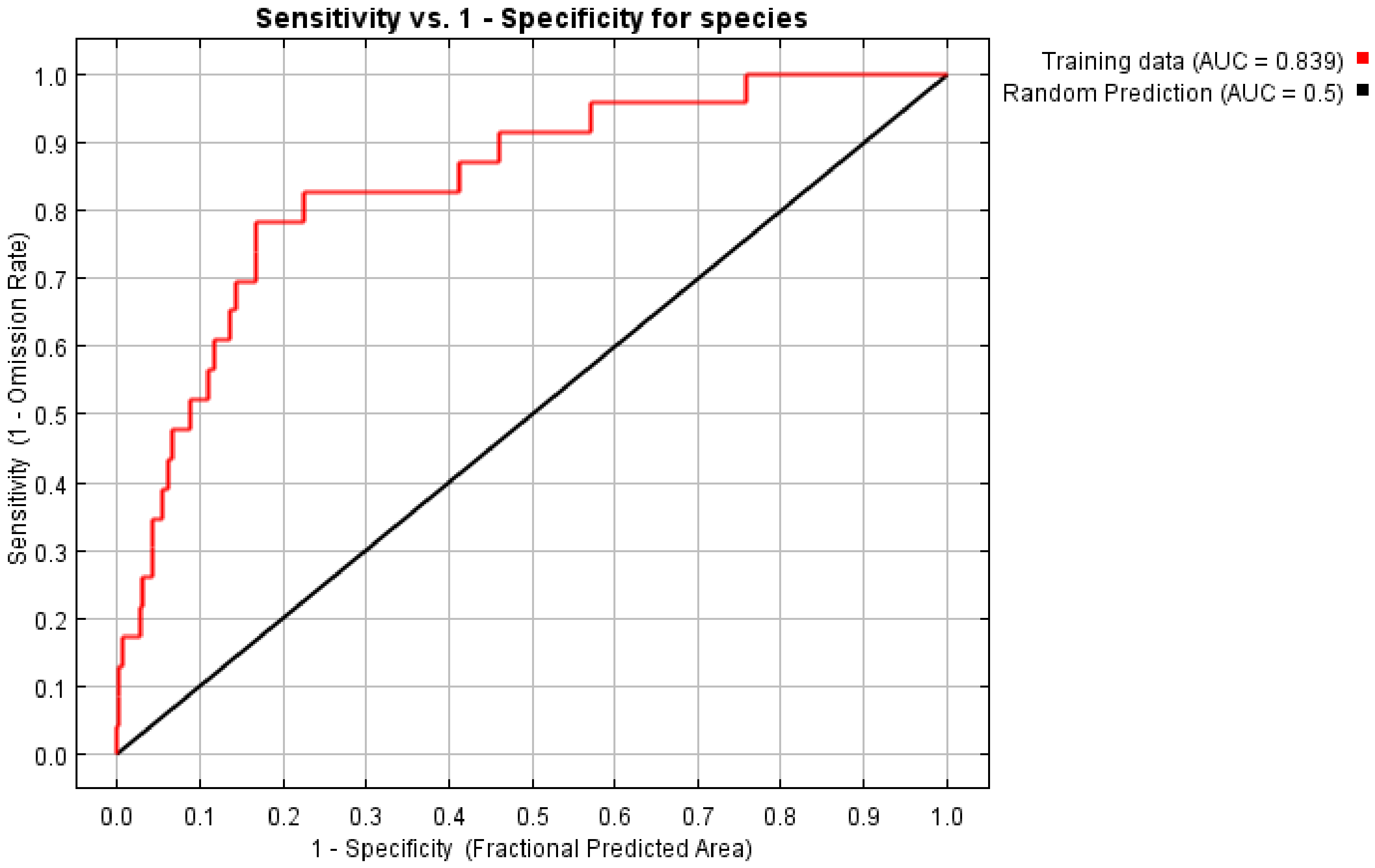

Sensitivity vs. 1 - Specificity for species
Training data (AUC = 0.839)
Random Prediction (AUC = 0.5)
Sensitivity (1 - Omission Rate)
1 - Specificity (Fractional Predicted Area)
0.0
0.1
0.2
0.3
0.4
0.5
0.6
0.7
0.8
0.9
1.0
